# Prediction of oncogene mutation status in non-small cell lung cancer: A systematic review and meta-analysis with a special focus on artificial-intelligence-based methods

**DOI:** 10.1101/2024.05.31.24308261

**Authors:** Almudena Fuster-Matanzo, Alfonso Picó Peris, Fuensanta Bellvís Bataller, Ana Jimenez-Pastor, Glen J. Weiss, Luis Martí-Bonmatí, Antonio Lázaro Sánchez, Giuseppe L. Banna, Alfredo Addeo, Ángel Alberich-Bayarri

**Author notes:** ^†^These authors contributed equally to this work. **Correspondence:** Almudena Fuster-Matanzo Quantitative Imaging Biomarkers in Medicine (Quibim), EDIFICIO EUROPA, Av. d’Aragó, 30, Planta 13 46021 Valencia, Spain Mail.

## Abstract

**Background:** In non-small cell lung cancer (NSCLC), alternative strategies to determine patient oncogene mutation status are essential to overcome some of the drawbacks associated with current methods. We aimed to review the use of radiomics alone or in combination with clinical data and to evaluate the performance of artificial intelligence (AI)-based models on the prediction of oncogene mutation status.

**Methods:** A PRISMA-compliant literature review was conducted. The Medline (via Pubmed), Embase, and Cochrane Library databases were searched for studies published through June 30, 2023 predicting oncogene mutation status in patients with NSCLC using radiomics. Independent meta-analyses evaluating the performance of AI-based models developed with radiomics features or with a combination of radiomics features plus clinical data for the prediction of different oncogenic driver mutations were performed. A meta-regression to analyze the influence of methodological/clinical factors was also conducted.

**Results:** Out of the 615 studies identified, 89 evaluating models for the prediction of epidermal growth factor-1 (EGFR), anaplastic lymphoma kinase (ALK), and Kirsten rat sarcoma virus (KRAS) mutations were included in the systematic review. A total of 38 met the inclusion criteria for the meta-analyses. The AI algorithms’ sensitivity/false positive rate (FPR) in predicting EGFR, ALK, and KRAS mutations using radiomics-based models was 0.753 (95% CI 0.721–0.783)/0.346 (95% CI 0.305–0.390), 0.754 (95% CI 0.639–0.841)/ 0.225 (95% CI 0.163–0.302), and 0.744 (95% CI 0.605–0.846)/0.376 (95% CI 0.274–0.491), respectively. A meta-analysis of combined models was only possible for EGFR mutation, revealing a sensitivity/FPR of 0.800 (95% CI 0.767–0.830)/0.335 (95% CI 0.279–0.396). No statistically significant results were obtained in the meta-regression.

**Conclusions:** Radiomics-based models may represent valuable non-invasive tools for the determination of oncogene mutation status in NSCLC. Further investigation is required to analyze whether clinical data might boost their performance.

## INTRODUCTION

Lung cancer represents the most often diagnosed cancer in both women and men worldwide, ranking first and third, respectively, and remaining the leading cause of cancer death^1^. Non-small cell lung cancer (NSCLC), the most frequent histological subtype, accounts for 80%–85% of cases, being adenocarcinoma the most common subtype (40%–50% of cases). Adenocarcinoma can be further subdivided into distinct molecular subtypes^2^. Indeed, molecular subtyping has become highly relevant in the disease context, as genotype-driven therapy (“targeted therapy”) is nowadays the standard of care for a significant subgroup of patients with advanced and metastatic NSCLC^3^. However, traditional methods for determining the molecular genotype, as well as the possible emergence of drug resistance mutations during patient’s follow-up, entail invasive biopsies and genetic sequence testing, procedures with multiple number of associated drawbacks including high costs, sampling bias, lack of enough sample, turnaround time, and medical complications^4-6^. Importantly, the overall accessibility of molecular diagnostics and liquid biopsy may be limited for many patients^7^, highlighting the need to investigate complementary methods to characterize the oncogene mutation status of lesions.

Radiological imaging represents a potent non-invasive tool for lung cancer, from the screening, diagnosis and staging of the disease to the management, therapeutic planification and follow-up of both early- and advanced-stage cases^8^. Specifically, computed tomography (CT) remains the standard of care for lung cancer visualization, providing excellent morphological and textural information. In recent years, radiomics, the process of extracting and analyzing quantitative features from medical images to investigate potential connections with biology and clinical outcomes, has gained increasing attention for its applicability in several oncological diseases including lung cancer^8^. The application of artificial intelligence (AI) to imaging analyses has enabled important clinical needs to be met. This includes the prognostication of outcomes or the prediction of response to treatment, disease progression, or the mutational and molecular profiling of tumors^9^. In particular, the use of radiomics coupled with AI methods has demonstrated to be a promising non-invasive alternative tool for the prediction of oncogene mutation status in NSCLC^8^.

In this systematic review and meta-analysis we aimed to: 1) review the available scientific evidence on the use of imaging-based models and radiomics for the prediction of the main targetable oncogenic driver alterations in NSCLC, including epidermal growth factor receptor (EGFR), anaplastic lymphoma kinase (ALK), and Kirsten rat sarcoma virus (KRAS); 2) analyze the overall performance of specifically, AI-based methods, for the prediction of oncogene mutation status; 3) evaluate whether the inclusion of clinical variables in the models improve their performance; 4) evaluate the impact of the available evidence from a clinical perspective.

## MATERIAL AND METHODS

This systematic review was conducted in accordance with the Preferred Reporting Items for Systematic Reviews and Meta-Analysis (PRISMA) guidelines^10^. The review was registered on PROSPERO before initiation (registration no. CRD42022349809).

### Search strategy

A systematic search for eligible publications published through 30 June 2023 was performed in Medline (via Pubmed), Cochrane Library and EMBASE databases using the keywords “Radiomics”, “NSCLC” and “Mutational status”. Further details on the search terms used in each database are provided in **Supplementary Table S1**. There were no limitations on the publishing year, participant age, or nationality. The search was exclusively limited to English-language publications.

### Study selection

Literature search and study selection were independently performed by two reviewers (A.F.M. and A.L.S.). To find relevant publications, they reviewed the titles and abstracts. Studies that satisfied the inclusion criteria were then manually assessed for eligibility by full-text screening. Covidence systematic review software (Veritas Health Innovation, Melbourne, Australia. Available at www.covidence.org) was used as a screening and data extraction tool.

#### Inclusion criteria

Papers were included in the qualitative synthesis (systematic review) if meeting the following inclusion criteria based on Patient, Index test, Comparator, Reference test, Diagnosis of reference (PIRD) questions: 1) being focused on the ability of radiomics to predict oncogene mutation status in NSCLC; 2) radiomics features were extracted from CT or from F-18 fluoro-deoxy-glucose (FDG)/CT scans; 3) a full text was available; 4) were written in English.

#### Exclusion criteria

Papers describing studies conducted using MRI scans (not the standard of care for NSCLC patients) or performed in phantom or animal models, or published as case reports, editorials, reviews, poster presentations, letters, editorials, or meeting abstracts were excluded. Papers not on the field of interest were also excluded.

For the quantitative synthesis (meta-analysis), the following additional exclusion criteria were applied: 1) oncogene mutation status was not the primary objective of the paper; 2) were focused on specific mutation subtypes; 3) did not apply AI-based methodologies; 4) developed simultaneous detection models or discriminant models; 5) sensitivity or specificity metrics were not available and could not be calculated; 6) were not comparable with the other articles included (model was developed based on intra- and extra-tumor derived radiomics features); 7) only included models developed with a combination of quantitative features extracted from PET/CT or from PET images (strictly adhering to a clinical perspective, PET scanning equipment is not always available and CT remains the standard of care for NSCLC patients); 8) did not reach a sufficient quality score according to the quality assessment (described below).

#### Quality assessment

The methodological quality of each study for its possible inclusion in the quantitative assessment was evaluated by using the Checklist for Artificial Intelligence in Medical Imaging (CLAIM)^11^. Classification, image reconstruction, text analysis, and workflow optimization are some of the applications of AI in medical imaging that are addressed by CLAIM, which is modeled after the Standards for Reporting of Diagnostic Accuracy Studies (STARD) guideline^12-15^. CLAIM checklist consists of 42 items divided into the conventional sections included in peer-reviewed scientific articles: title or abstract (1 item), abstract (1 item), introduction (2 items), methods (28 items subdivided into study design [2 items], data [7 items], ground truth [5 items], data partitions [3 items], model [3 items], training [3 items] and evaluation [5 items]), results (5 items subdivided into data [2 items] and model performance [3 items]), discussion (2 items) and other information (3 items). The CLAIM guideline offers a roadmap for writers and reviewers with the intention of fostering clear, open, and verifiable scientific discourse on the use of AI in medical imaging^11^.

For our quality assessment, a score was calculated for each paper ([total score, 42 - number of “not applicable” fields in each case]). A cut-off value of at least half of the total score after removing the “not applicable” items was established for the inclusion in the quantitative analysis. Therefore, this cut-off value varied for each study depending on the number of items that were applicable from among the 42 total items included in the CLAIM checklist (e.g., a cut-off value of 19 was established for those studies in which only 38 items of the checklist were applicable). **See Supplementary Table S2**. The assessment of the rigor, quality, and generalizability of the work of all enrolled studies was performed by three reviewers (A.J.P., F.B.B. and A.P.P.).

### Data extraction

Data extracted included the following: (1) study details: first author, publication year, research questions, study design; (2) patient details: the source of data acquisition (single-center/multicenter), sample size, smoking history, age, sex, TNM staging, treatment status (naïve or any treatment received prior image acquisition), histological subtype; (3) imaging details: imaging modality, plain or contrast CT; (4) oncogene mutation status-related information: type of mutation, specific subtype of mutation (if available), sequencing method; sequencing kit (5) radiomics details: segmentation software, type of segmentation (manual, automatic, or semi-automatic), radiomics feature extraction software, number of imaging features extracted, number and name of radiomics features included in final models, features selection methods, type of models constructed (machine learning [ML], deep learning [DL], classical statistical model), final classifier used in machine learning models, clinical variables included in the models (if applicable), and models performance. Two independent reviewers (A.F.M. and A.L.S.) completed the initial screening and extracted data from all included studies.

### Data analysis

For studies including models based on features extracted from different imaging modalities, only those based on CT scans were included in the quantitative analysis. A bivariate analysis of sensitivity and specificity as proposed by Reitsma et al.^16^ was chosen to perform the meta-analyses. This method has the distinct advantage of preserving the two-dimensional nature of the underlying data. It can also produce summary estimates of sensitivity and specificity (false positive rate [FPR, 1-specificity]), recognizing any possible correlation between these two measures. The method uses a random effect approach in which the values of the sensitivity and FPR estimates are obtained with restricted maximum likelihood. As a complement to the bivariate approach, the summary receiver operating characteristic (sROC) was calculated by converting each pair of sensitivity and specificity into a single measure of accuracy, the diagnostic odds ratio (DOR).

The analyses were carried out by reproducing the confusion matrices of each model presented in the studies, the number of cases and the prevalence of oncogene mutant positive cases. All calculations were performed on the basis of validation cohorts for studies applying a training/validation split method, or on the basis of the total sample when cross-validation was the validation strategy. To ensure homogeneity, calculations were conducted based on internal validation cohort data when external validation was also performed (minority of the cases).

Finally, a meta-regression analysis was performed to measure the possible influence of the following predictors: (1) average age of the cases, (2) manual segmentation vs semi-automatic segmentation vs both procedures (no studies including automatic segmentation approaches met the inclusion criteria for the quantitative analysis), (3) whether the model included only radiomics features or was combined with clinical variables, and (4) whether the model was classified as ML or DL. The heterogeneity in the description of the clinical variables included in the models prevented the inclusion of additional predictors of greatest clinical interest. Only the best model from each study according to its DOR was selected. When the mean/median age was not available due to the heterogeneity among studies when presenting descriptive results, it was inferred from the information obtained. Thus, mean and median values were indistinctly considered; when both values were provided, an average of both was calculated. If mean values were absent, median values were considered and viceversa. If both values were absent from the validation cohort, mean/median age from the total cohort was considered. When this information was not available either, the study was not included in the meta-regression.

All the analyses were performed using R Statistical Software v4.2.2 and the packages mada and tidyverse.

## RESULTS

In total 615 articles were obtained according to the search strategy (**Figure 1**). After de-duplication, 397 studies were obtained and screened. According to the inclusion and exclusion criteria, 89 studies were included in the qualitative analysis (systematic review), all of them developing models for the prediction of EGFR, ALK, and/or KRAS. Out of those, 38 were found eligible for the quantitative part of the study (meta-analyses). As detailed in **Supplementary Table S2**, all papers passed this quality check and were therefore included.

**Figure 1.**
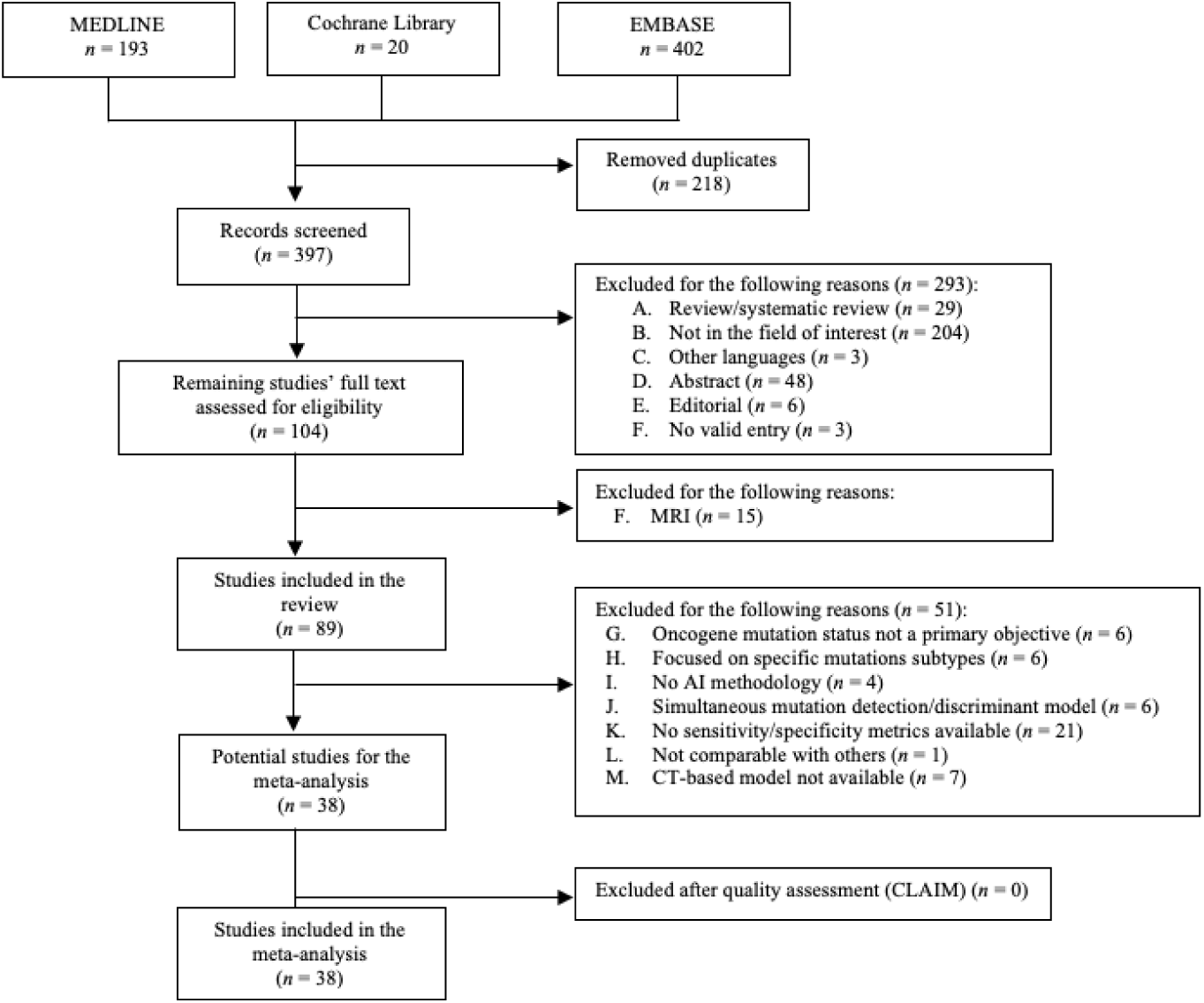
PRISMA flowchart. AI, artificial intelligence; CLAIM, Checklist for Artificial Intelligence in Medical Imaging; CT, computed tomography; MRI, magnetic resonance imaging.

### Qualitative analysis (systematic review)

#### Methodological characteristics of the studies

The methodological characteristics of the studies are summarized in **Table 1**. Most of the studies (*n* = 69/ 89) applied exclusively ML algorithms, while this methodology was also used to build comparator models in 10 articles in which DL techniques were the main methodological approach followed. Only three studies exclusively applied DL algorithms, while classical statistical models were used in seven publications. Among the 79 articles applying ML techniques, the most common classifier used was logistic regression (*n* = 38), followed by support vector machine (*n* = 35) and by random forest (*n* = 29). In terms of partitioning strategy, training-validation split was the most frequent technique (*n* = 71). External validation was only performed in a small set of studies (*n* = 9). Regarding imaging techniques, CT was the most frequently used modality (*n* = 61), followed by PET/CT (*n* = 22) and by PET alone (*n* = 4). Additionally, in one study^17^ PET/CT scans and contrast-enhanced CT images independently acquired were collected, while in another study^18^, PET/CT, CT, and contrast-enhanced diagnostic quality (CTD) images were used. Of the 61 studies conducted with CT scans, 39 included non-contrast-enhanced images, 18 contrast-enhanced images, in two contrast-related information was not specified and in two both contrast- and non-contrast-enhanced scans were included. Regarding tumor segmentation, a manual approach was followed in 48 studies, and automatic and semi-automatic segmentations were applied in two and 31 studies, respectively; three studies applied both methodologies (for verification or a different approach according to the imaging modality used) and five studies did not specify the method utilized for tumor segmentation.

**Table 1.**
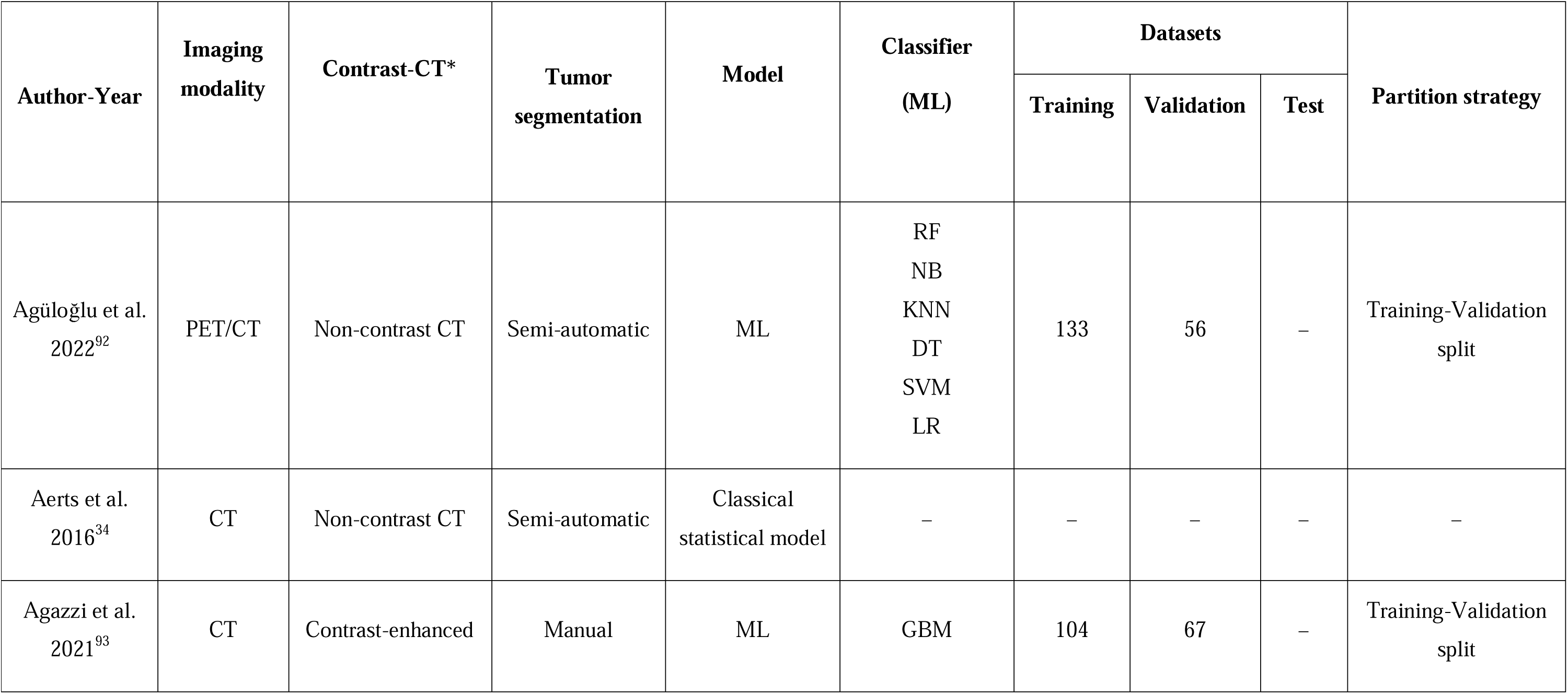
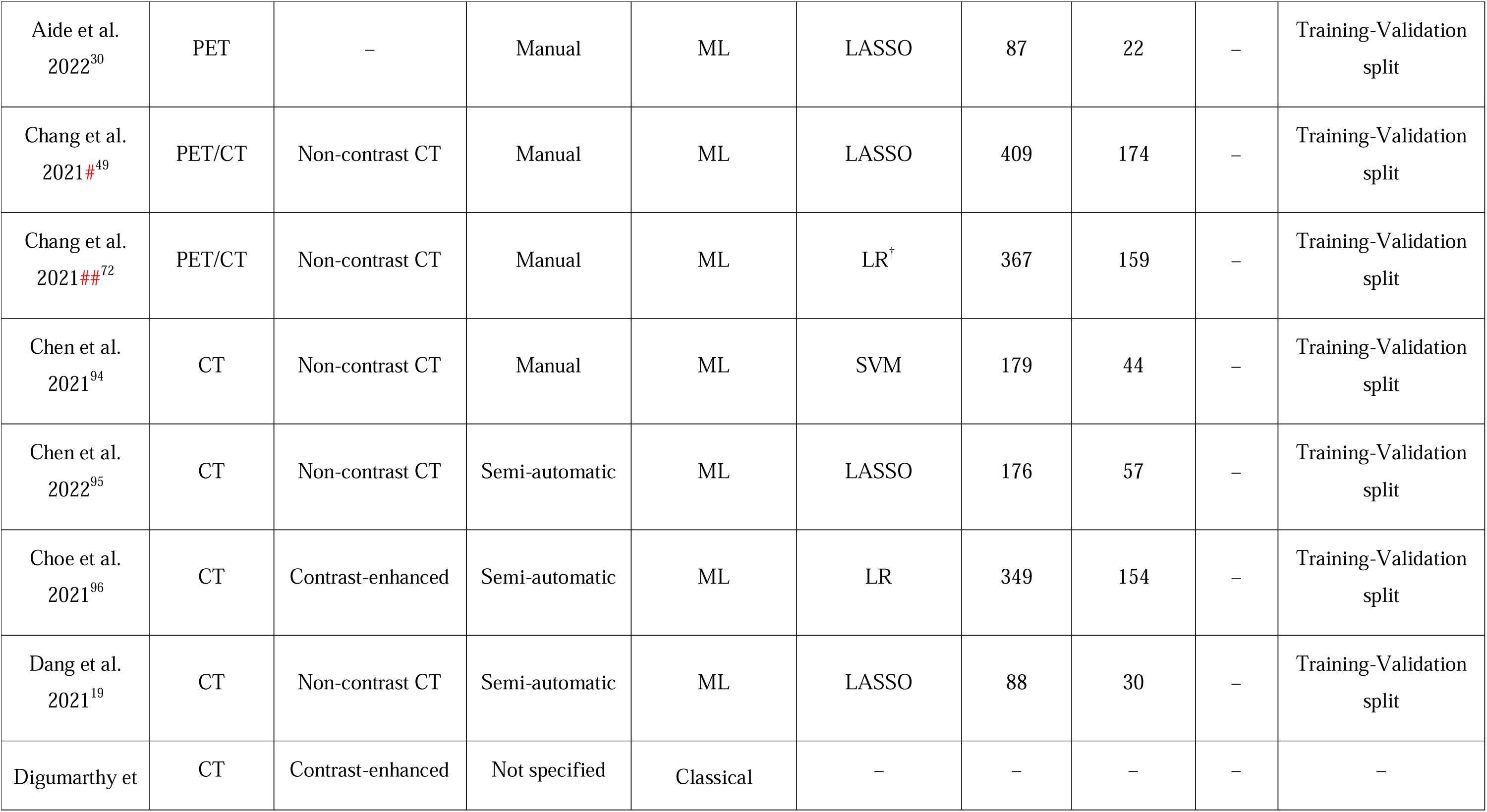
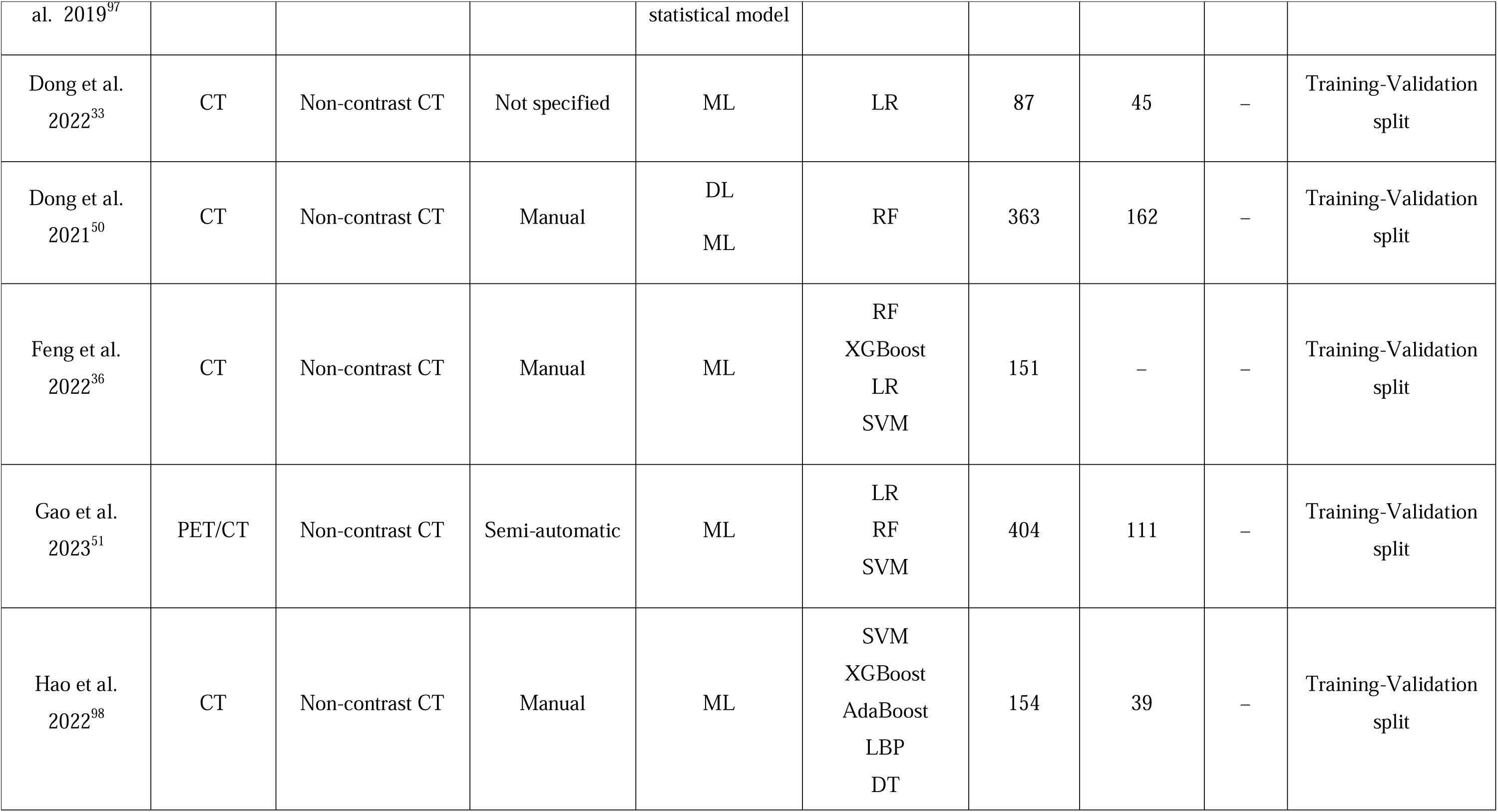
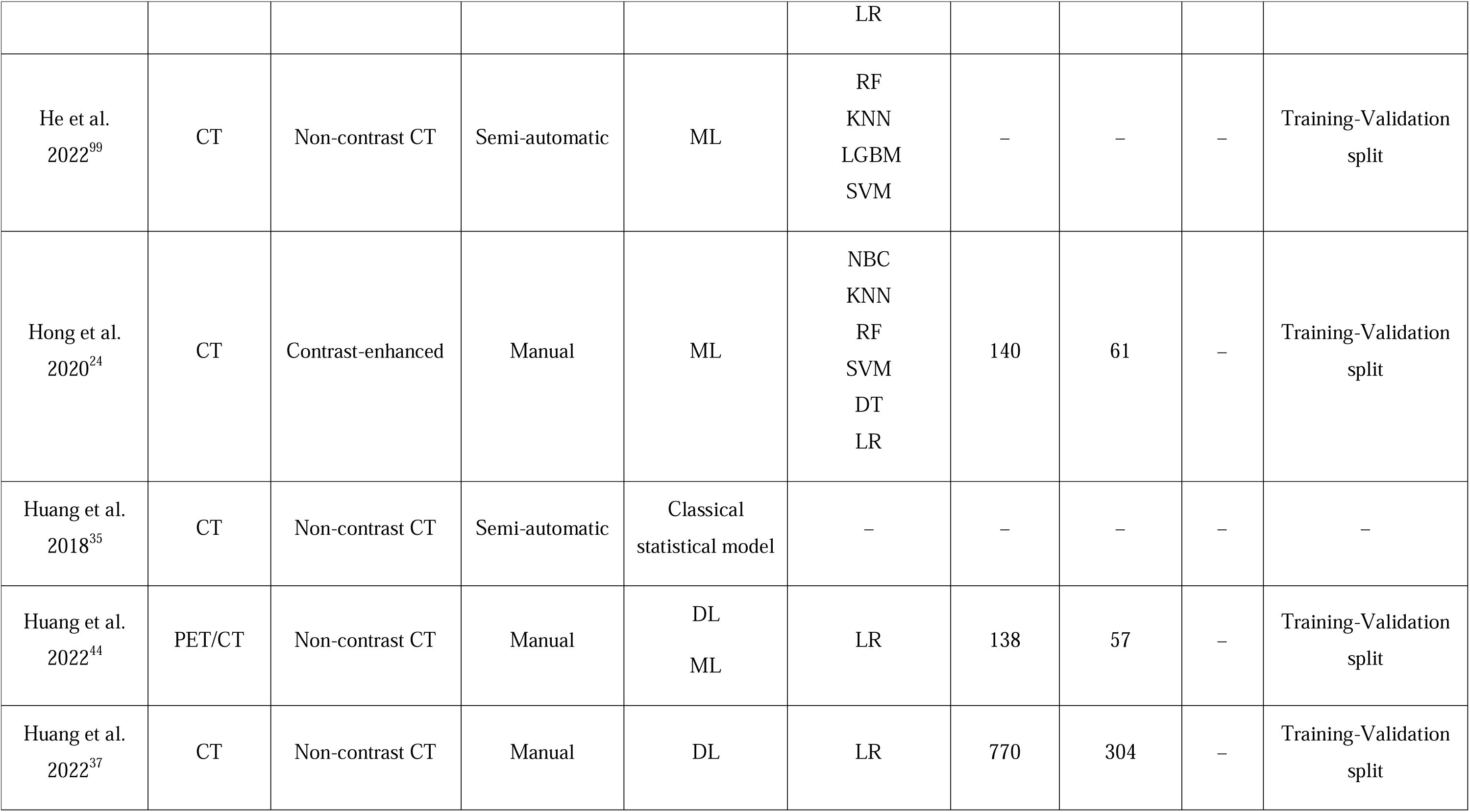
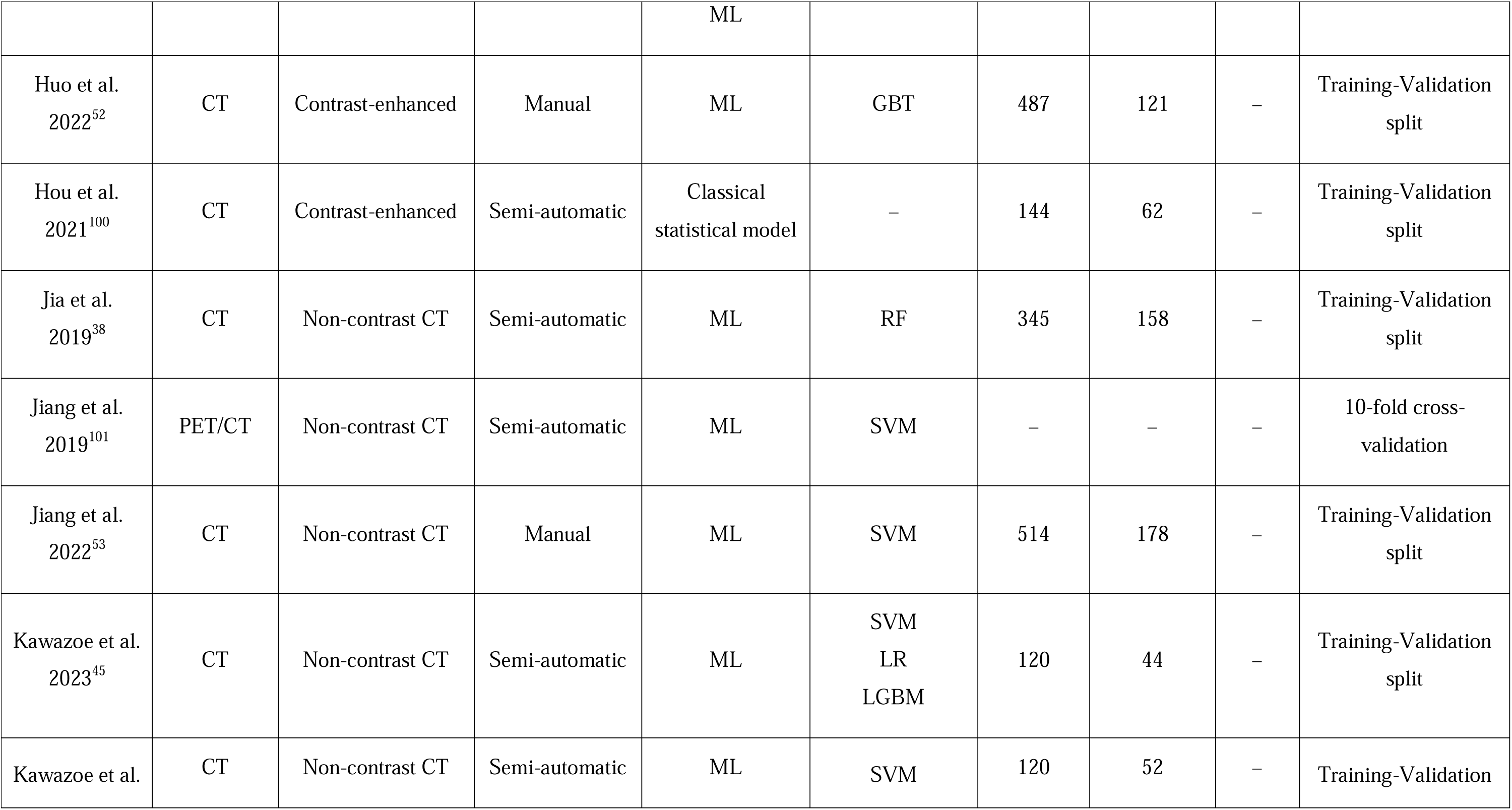
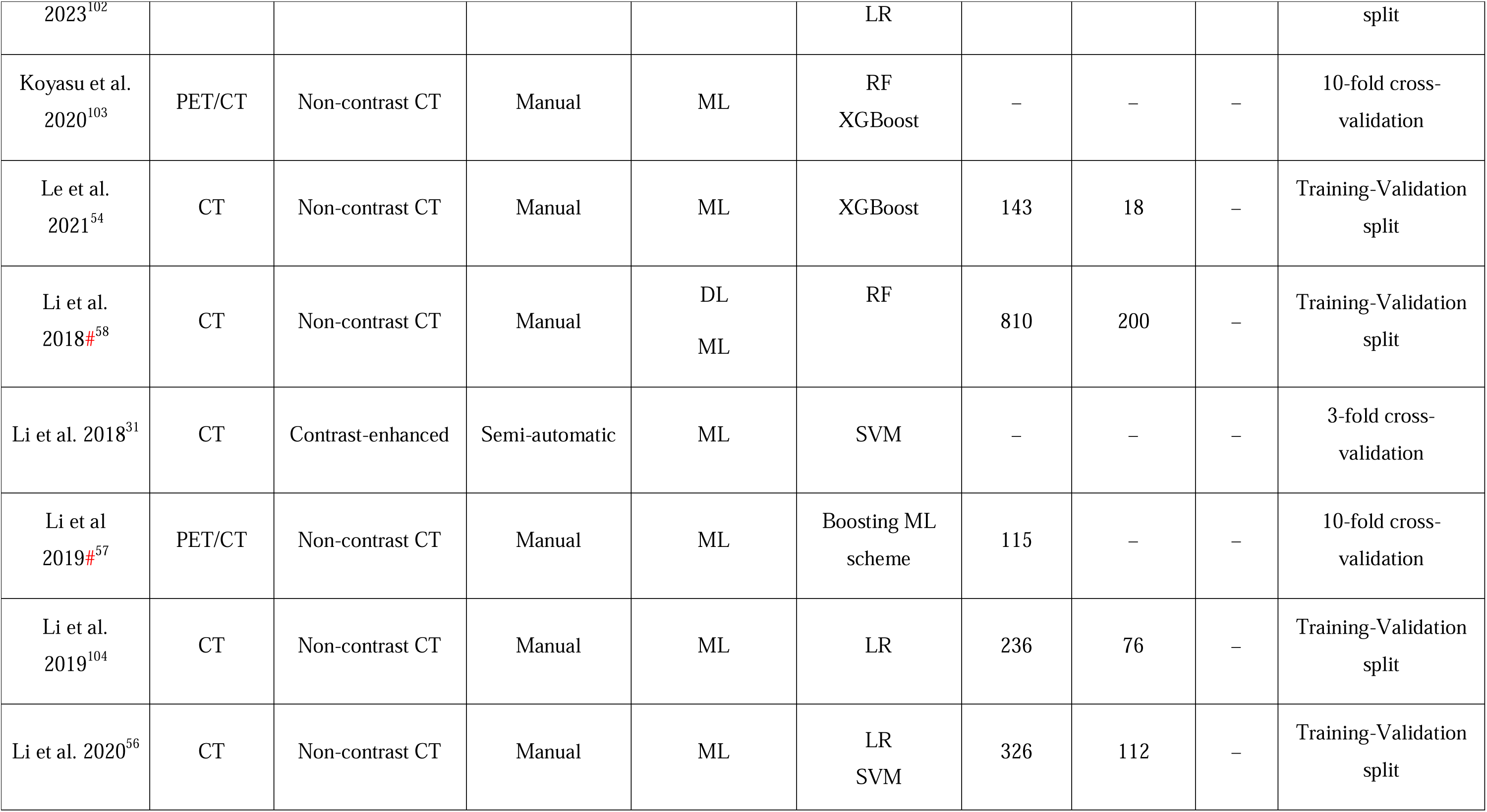
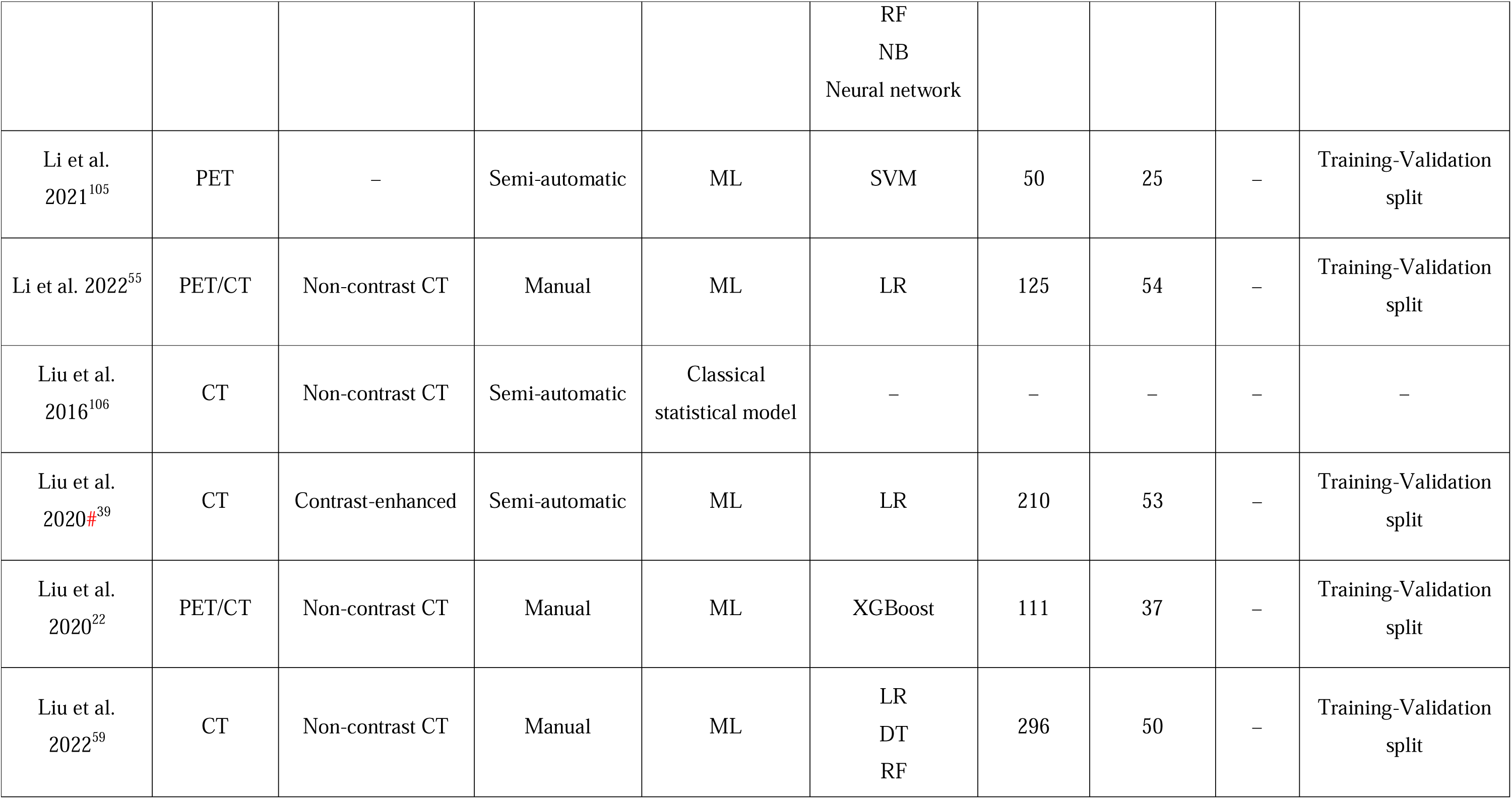
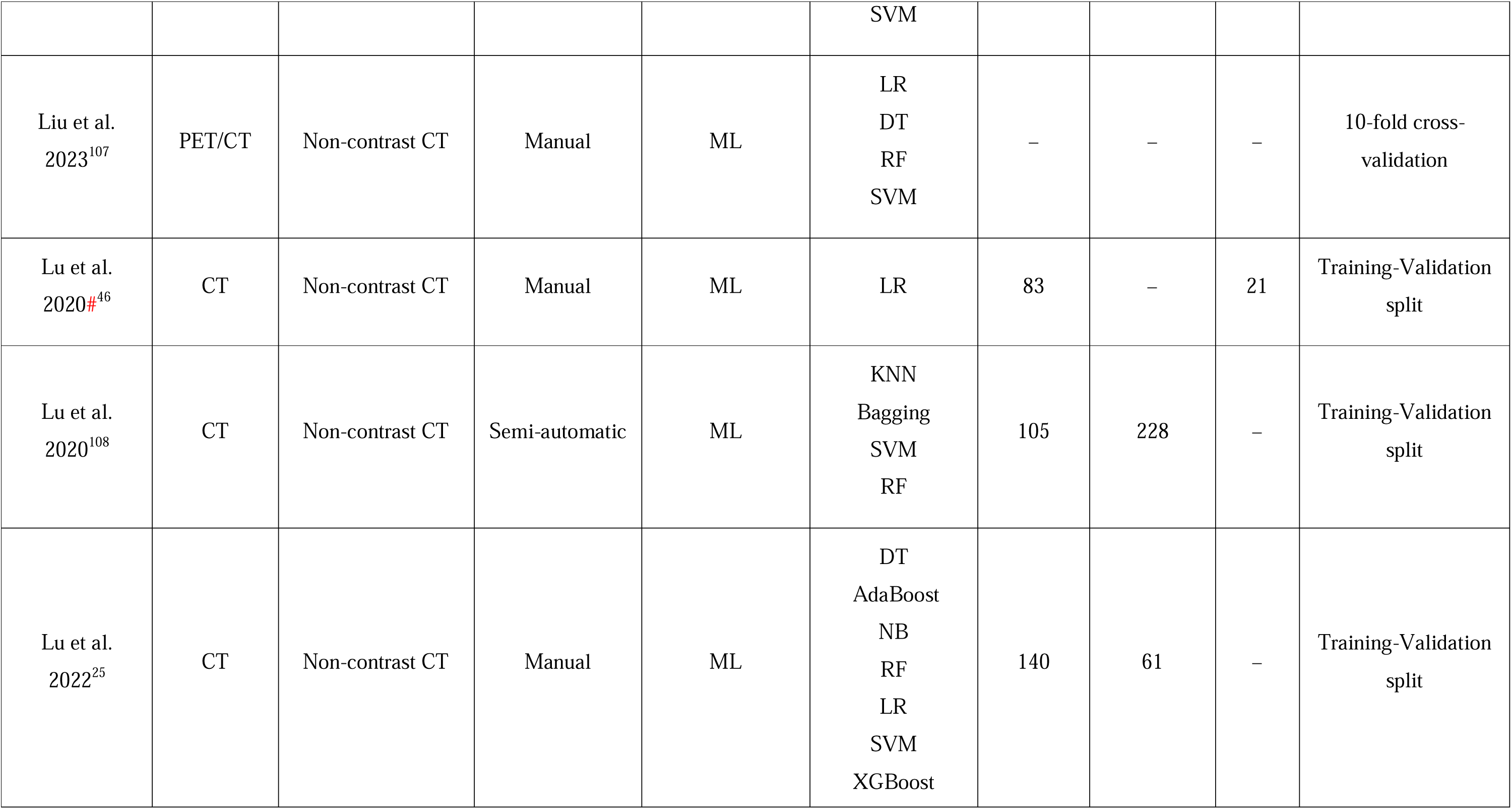
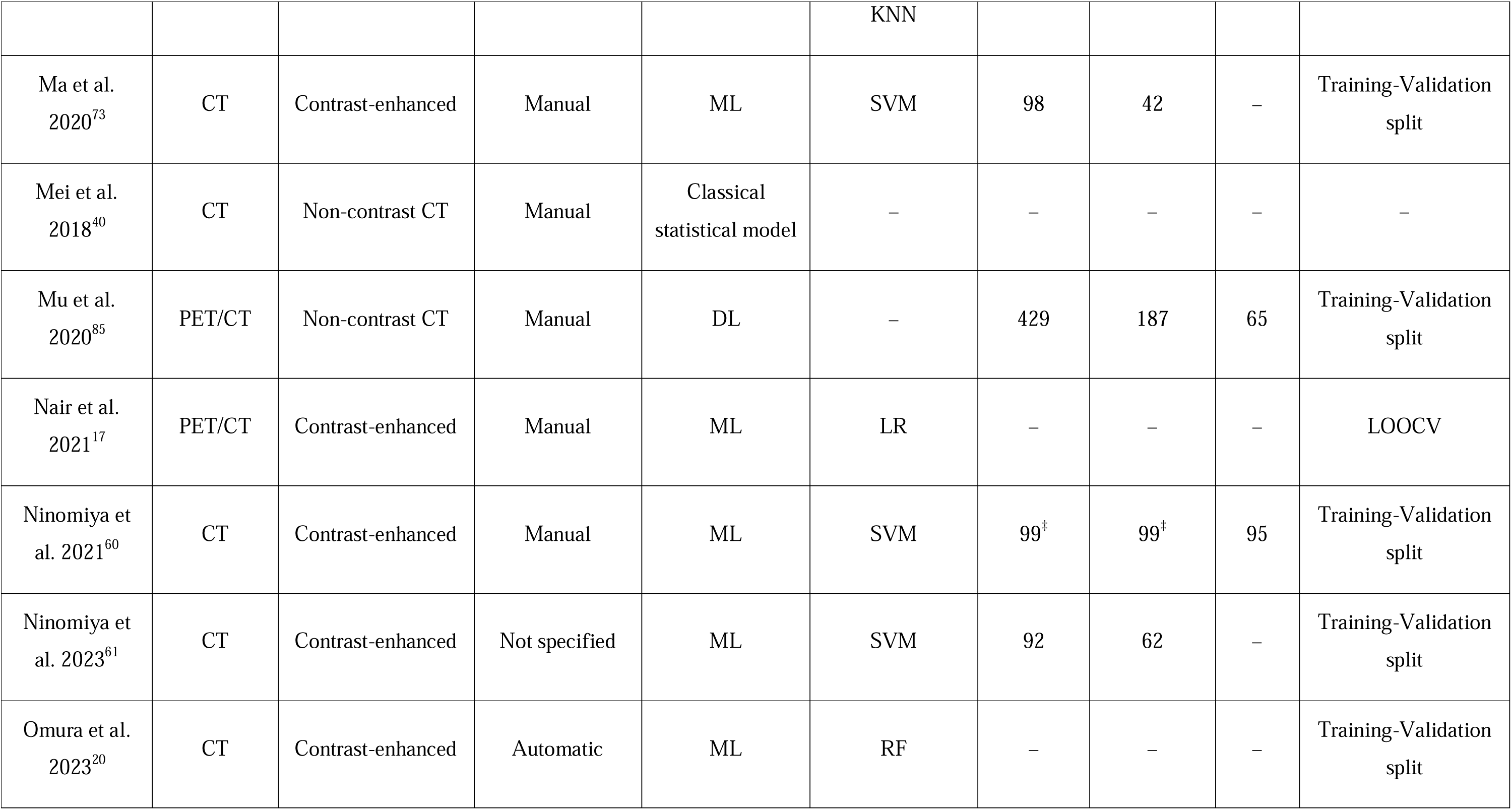
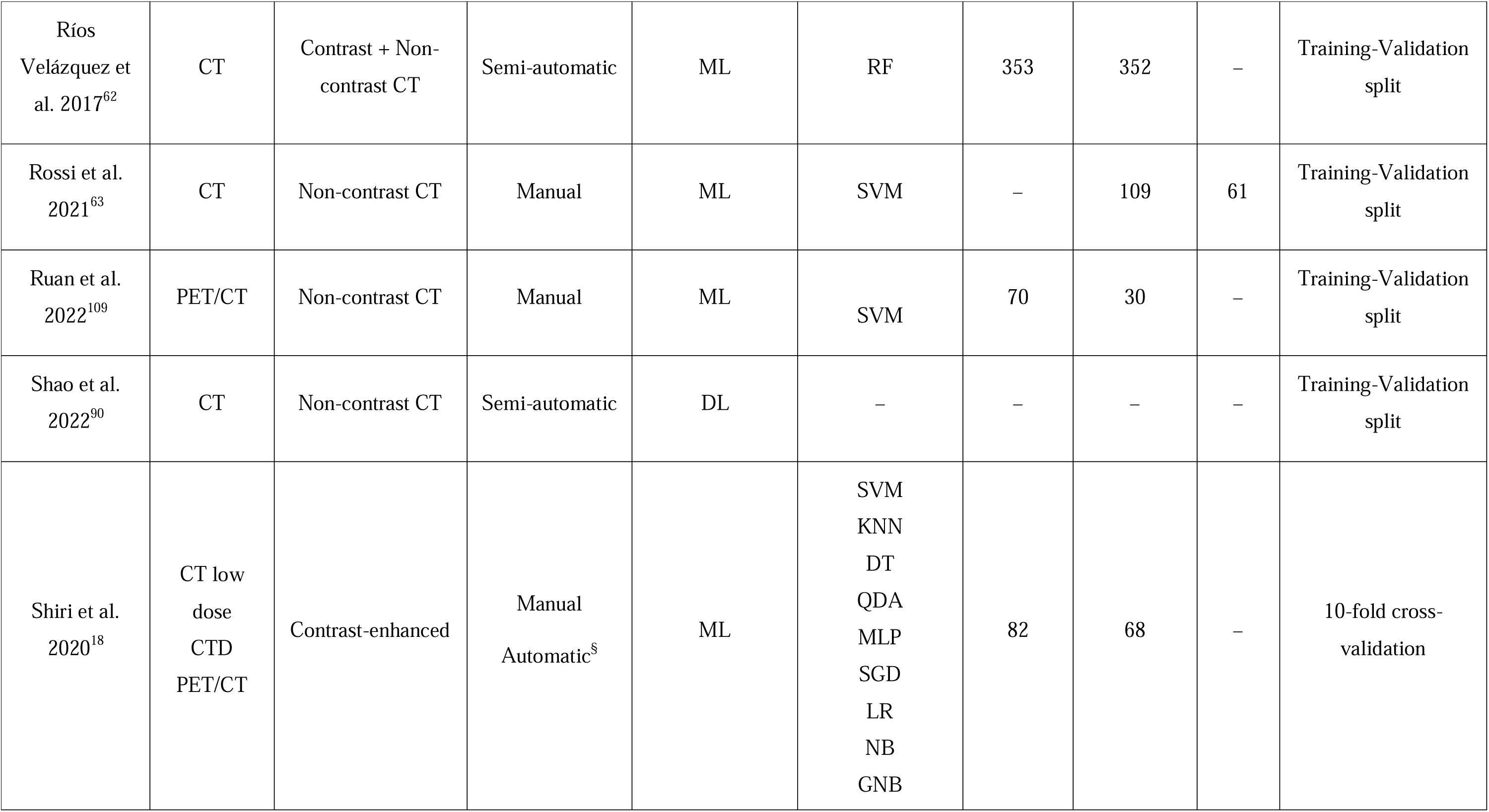
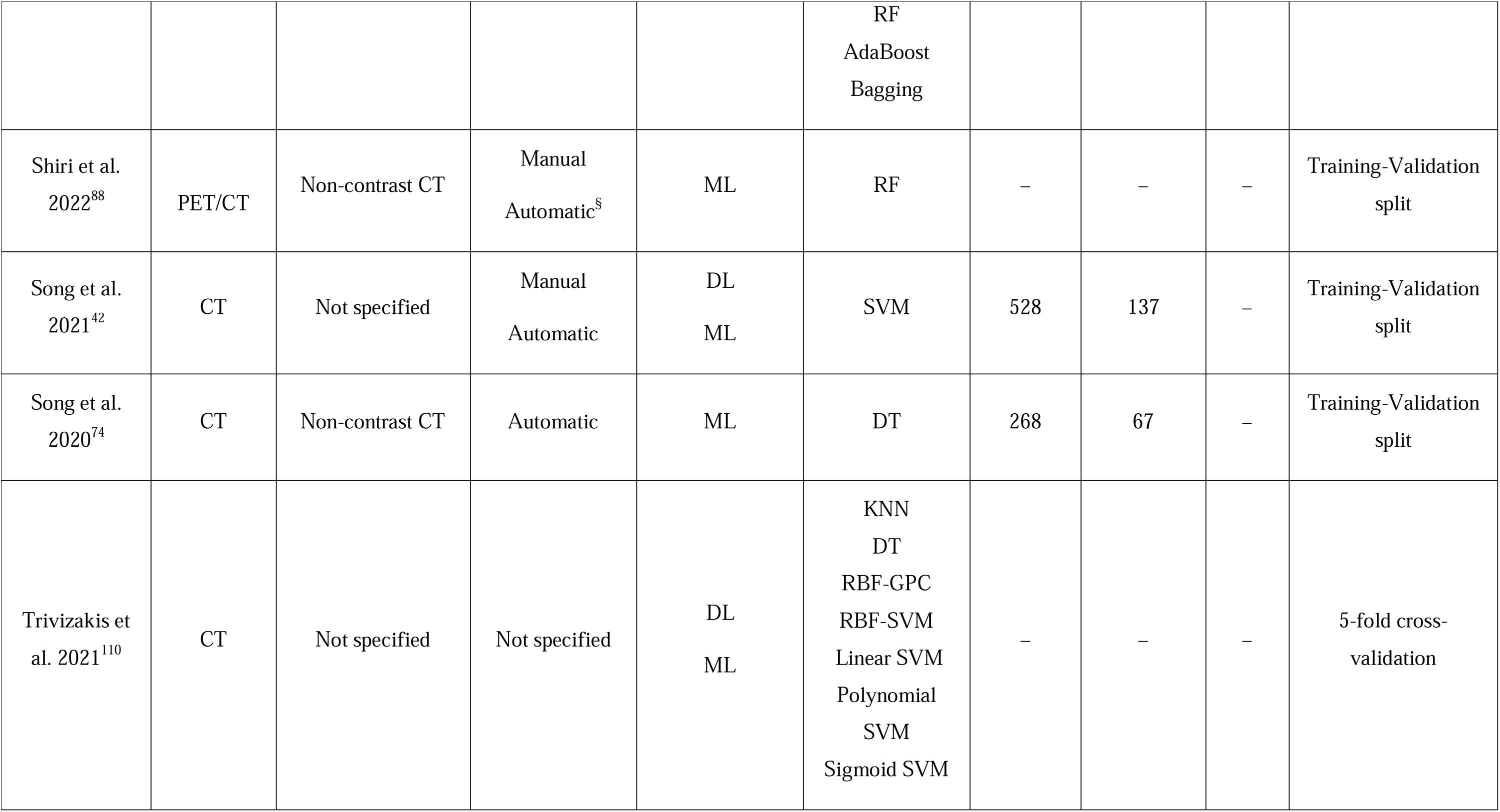
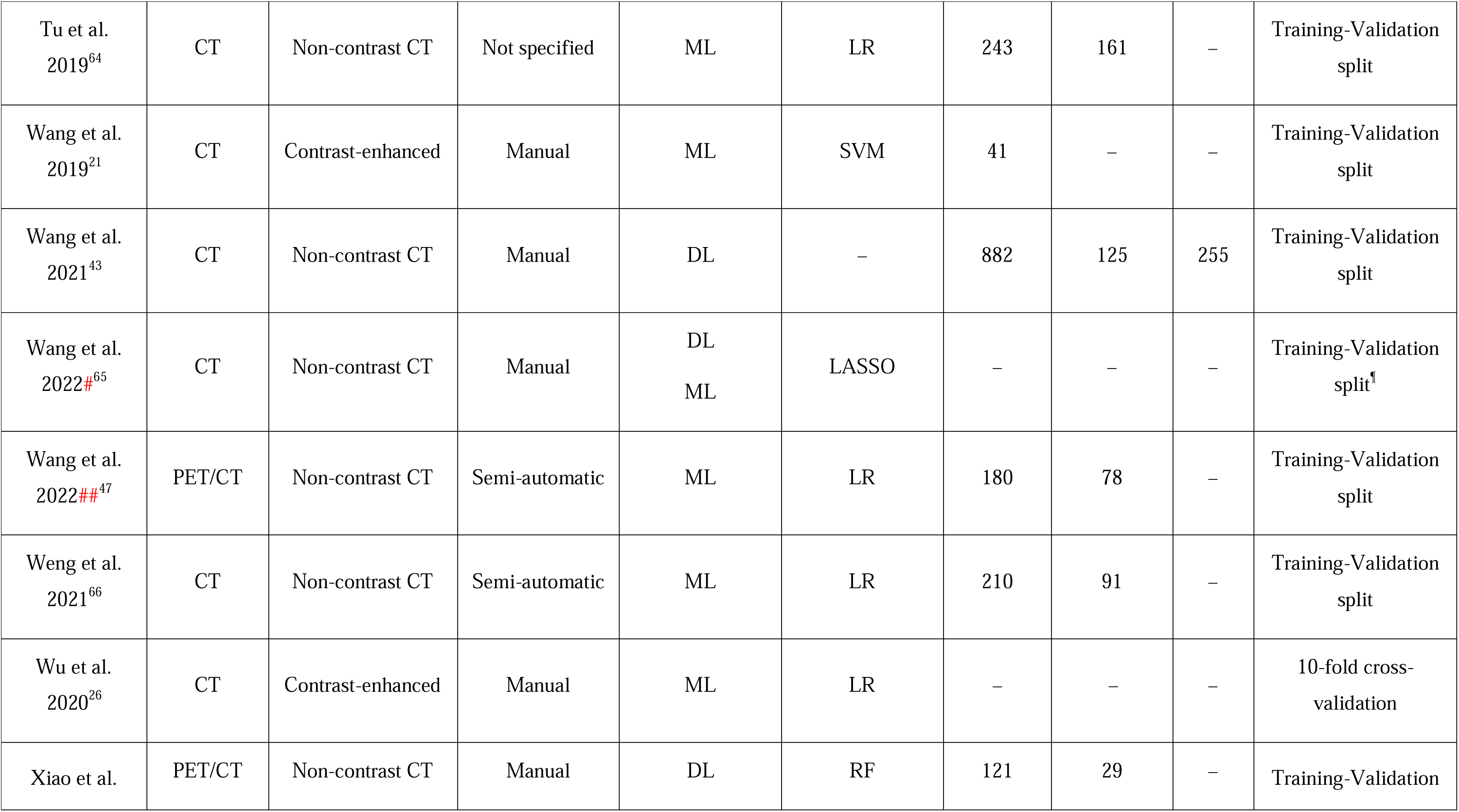
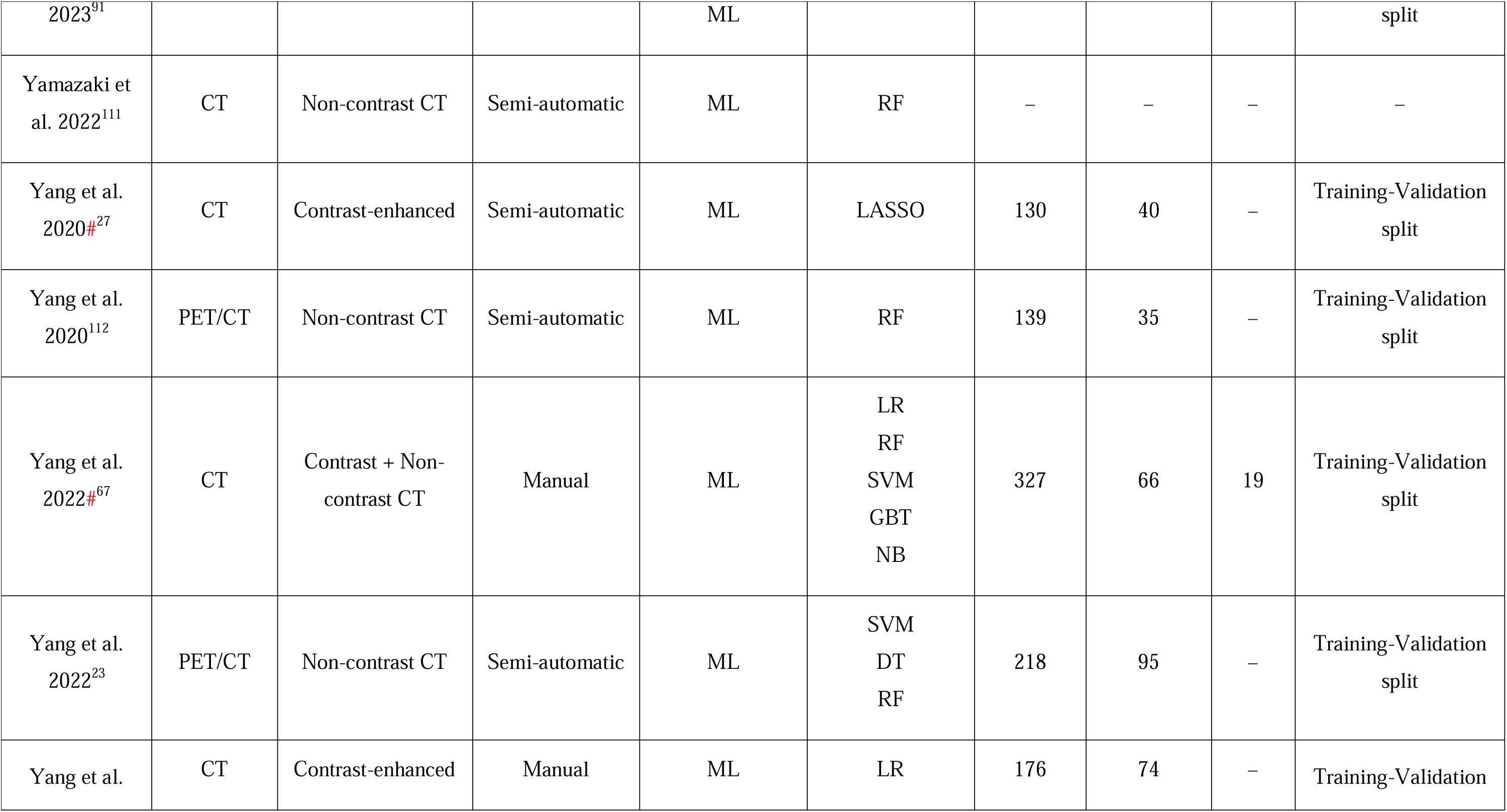
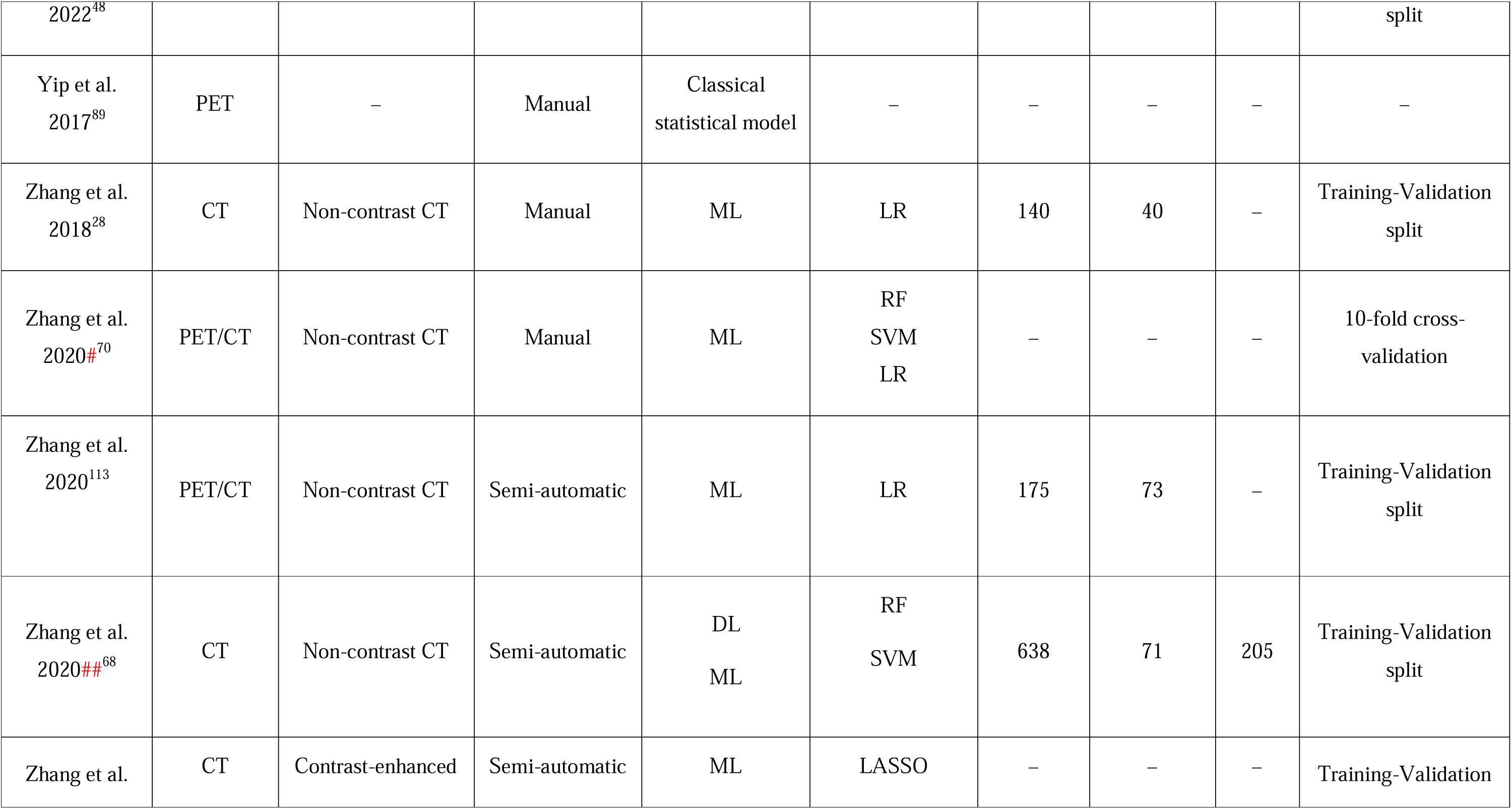
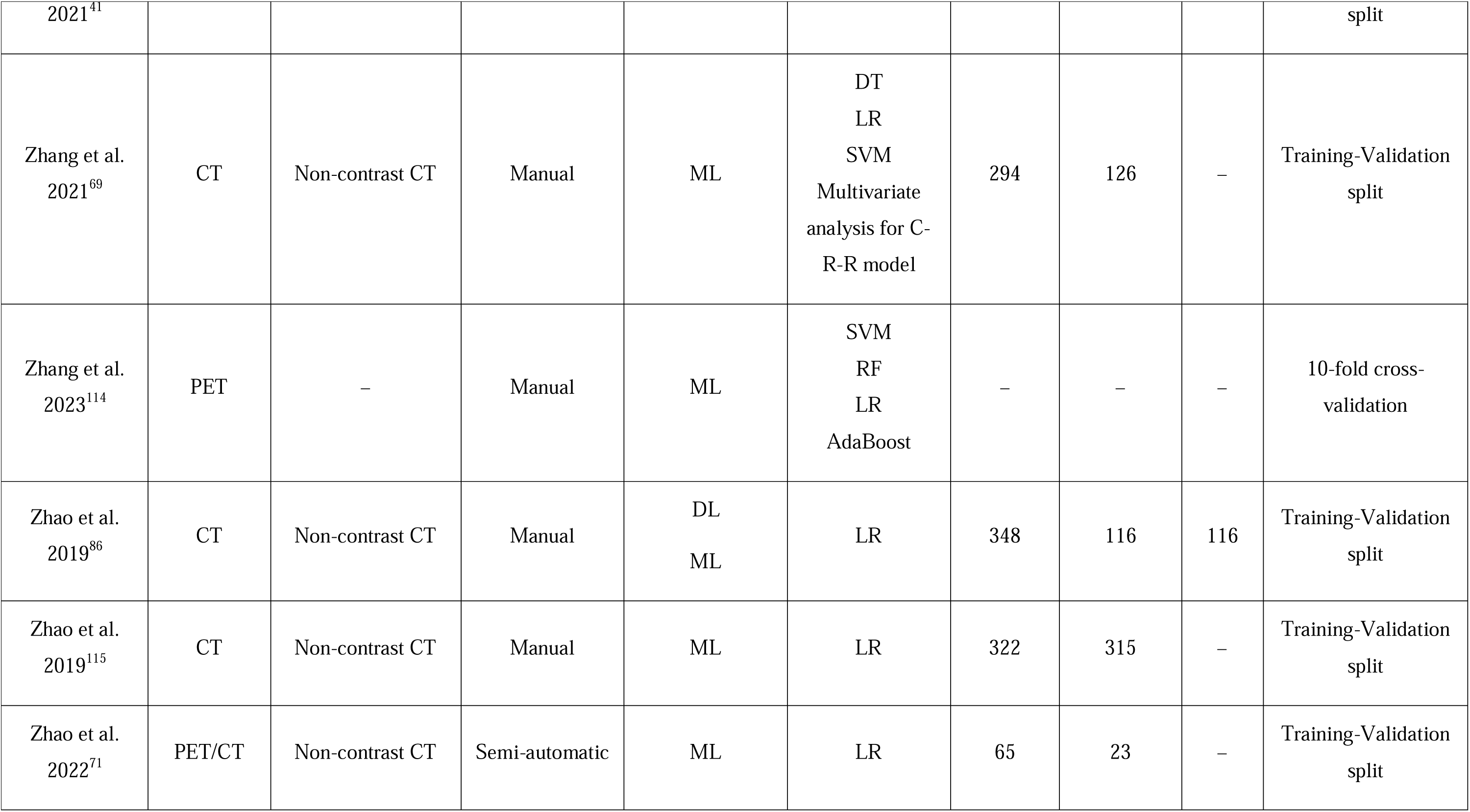
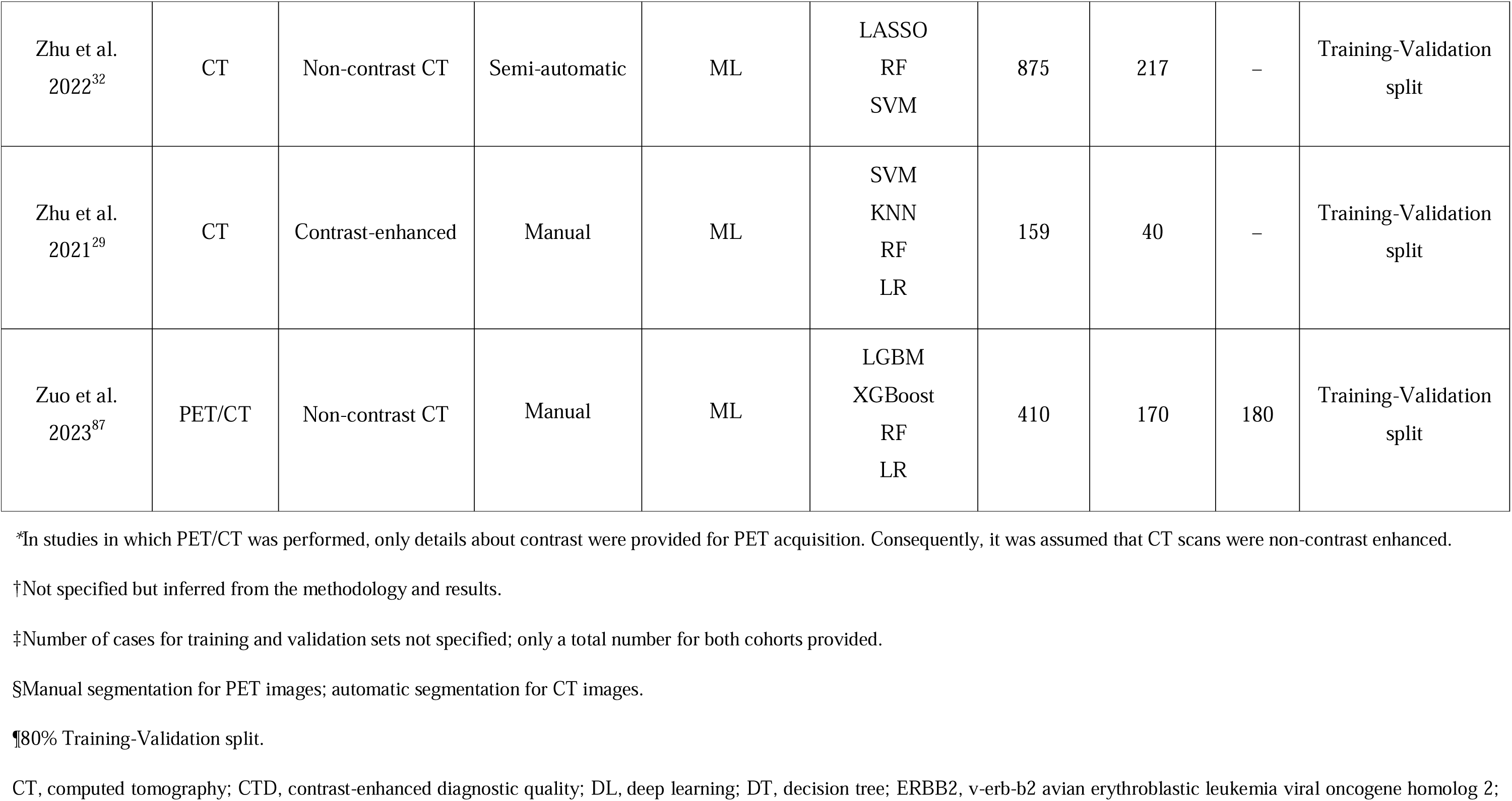
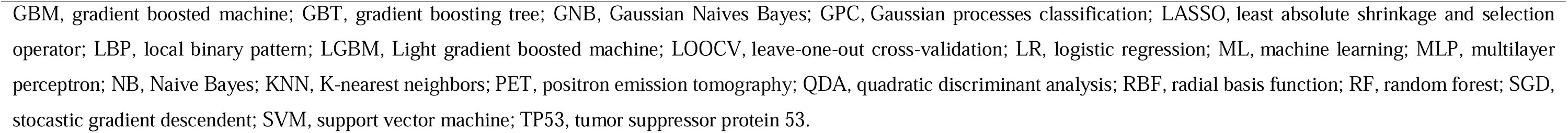
Methodological characteristics of the studies (*N* = 89) included in the systematic review. For those studies with the same name for the first author and published the same year, a hashtag was added to unequivocally indicate those that were included in the different meta-analyses and consequently, that are represented in the forest plots.

| Author-Year | Imaging modality | Contrast-CT* | Tumor segmentation | Model | Classifier (ML) | Datasets |  |  | Partition strategy |
| --- | --- | --- | --- | --- | --- | --- | --- | --- | --- |
|  |  |  |  |  |  | Training | Validation | Test |  |
| Agüloğlu et al. 2022<sup>92</sup> | PET/CT | Non-contrast CT | Semi-automatic | ML | RF<br>NB<br>KNN<br>DT<br>SVM<br>LR | 133 | 56 | – | Training-Validation split |
| Aerts et al. 2016<sup>34</sup> | CT | Non-contrast CT | Semi-automatic | Classical statistical model | – | – | – | – | – |
| Agazzi et al. 2021<sup>93</sup> | CT | Contrast-enhanced | Manual | ML | GBM | 104 | 67 | – | Training-Validation split |
| Aide et al.<br>2022<sup>30</sup> | PET | – | Manual | ML | LASSO | 87 | 22 | – | Training-Validation<br>split |
| Chang et al.<br>2021<sup>#49</sup> | PET/CT | Non-contrast CT | Manual | ML | LASSO | 409 | 174 | – | Training-Validation<br>split |
| Chang et al.<br>2021<sup>##72</sup> | PET/CT | Non-contrast CT | Manual | ML | LR<sup>†</sup> | 367 | 159 | – | Training-Validation<br>split |
| Chen et al.<br>2021<sup>94</sup> | CT | Non-contrast CT | Manual | ML | SVM | 179 | 44 | – | Training-Validation<br>split |
| Chen et al.<br>2022<sup>95</sup> | CT | Non-contrast CT | Semi-automatic | ML | LASSO | 176 | 57 | – | Training-Validation<br>split |
| Choe et al.<br>2021<sup>96</sup> | CT | Contrast-enhanced | Semi-automatic | ML | LR | 349 | 154 | – | Training-Validation<br>split |
| Dang et al.<br>2021<sup>19</sup> | CT | Non-contrast CT | Semi-automatic | ML | LASSO | 88 | 30 | – | Training-Validation<br>split |
| Digumarthy et | CT | Contrast-enhanced | Not specified | Classical | – | – | – | – | – |
| al. 2019<sup>97</sup> |  |  |  | statistical model |  |  |  |  |  |
| Dong et al.<br>2022<sup>33</sup> | CT | Non-contrast CT | Not specified | ML | LR | 87 | 45 | – | Training-Validation<br>split |
| Dong et al.<br>2021<sup>50</sup> | CT | Non-contrast CT | Manual | DL<br>ML | RF | 363 | 162 | – | Training-Validation<br>split |
| Feng et al.<br>2022<sup>36</sup> | CT | Non-contrast CT | Manual | ML | RF<br>XGBoost<br>LR<br>SVM | 151 | – | – | Training-Validation<br>split |
| Gao et al.<br>2023<sup>51</sup> | PET/CT | Non-contrast CT | Semi-automatic | ML | LR<br>RF<br>SVM | 404 | 111 | – | Training-Validation<br>split |
| Hao et al.<br>2022<sup>98</sup> | CT | Non-contrast CT | Manual | ML | SVM<br>XGBoost<br>AdaBoost<br>LBP<br>DT | 154 | 39 | – | Training-Validation<br>split |
|  |  |  |  |  | LR |  |  |  |  |
| He et al.<br>2022<sup>99</sup> | CT | Non-contrast CT | Semi-automatic | ML | RF<br>KNN<br>LGBM<br>SVM | – | – | – | Training-Validation<br>split |
| Hong et al.<br>2020<sup>24</sup> | CT | Contrast-enhanced | Manual | ML | NBC<br>KNN<br>RF<br>SVM<br>DT<br>LR | 140 | 61 | – | Training-Validation<br>split |
| Huang et al.<br>2018<sup>35</sup> | CT | Non-contrast CT | Semi-automatic | Classical<br>statistical model | – | – | – | – | – |
| Huang et al.<br>2022<sup>44</sup> | PET/CT | Non-contrast CT | Manual | DL<br>ML | LR | 138 | 57 | – | Training-Validation<br>split |
| Huang et al.<br>2022<sup>37</sup> | CT | Non-contrast CT | Manual | DL | LR | 770 | 304 | – | Training-Validation<br>split |
|  |  |  |  | ML |  |  |  |  |  |
| Huo et al.<br>2022<sup>52</sup> | CT | Contrast-enhanced | Manual | ML | GBT | 487 | 121 | – | Training-Validation<br>split |
| Hou et al.<br>2021<sup>100</sup> | CT | Contrast-enhanced | Semi-automatic | Classical<br>statistical model | – | 144 | 62 | – | Training-Validation<br>split |
| Jia et al.<br>2019<sup>38</sup> | CT | Non-contrast CT | Semi-automatic | ML | RF | 345 | 158 | – | Training-Validation<br>split |
| Jiang et al.<br>2019<sup>101</sup> | PET/CT | Non-contrast CT | Semi-automatic | ML | SVM | – | – | – | 10-fold cross-validation |
| Jiang et al.<br>2022<sup>53</sup> | CT | Non-contrast CT | Manual | ML | SVM | 514 | 178 | – | Training-Validation<br>split |
| Kawazoe et al.<br>2023<sup>45</sup> | CT | Non-contrast CT | Semi-automatic | ML | SVM<br>LR<br>LGBM | 120 | 44 | – | Training-Validation<br>split |
| Kawazoe et al. | CT | Non-contrast CT | Semi-automatic | ML | SVM | 120 | 52 | – | Training-Validation |
| 2023<sup>102</sup> |  |  |  |  | LR |  |  |  | split |
| Koyasu et al.<br>2020<sup>103</sup> | PET/CT | Non-contrast CT | Manual | ML | RF<br>XGBoost | – | – | – | 10-fold cross-validation |
| Le et al.<br>2021<sup>54</sup> | CT | Non-contrast CT | Manual | ML | XGBoost | 143 | 18 | – | Training-Validation<br>split |
| Li et al.<br>2018<sup>#58</sup> | CT | Non-contrast CT | Manual | DL<br>ML | RF | 810 | 200 | – | Training-Validation<br>split |
| Li et al. 2018<sup>31</sup> | CT | Contrast-enhanced | Semi-automatic | ML | SVM | – | – | – | 3-fold cross-validation |
| Li et al<br>2019<sup>#57</sup> | PET/CT | Non-contrast CT | Manual | ML | Boosting ML<br>scheme | 115 | – | – | 10-fold cross-validation |
| Li et al.<br>2019<sup>104</sup> | CT | Non-contrast CT | Manual | ML | LR | 236 | 76 | – | Training-Validation<br>split |
| Li et al. 2020<sup>56</sup> | CT | Non-contrast CT | Manual | ML | LR<br>SVM | 326 | 112 | – | Training-Validation<br>split |
|  |  |  |  |  | RF<br>NB<br>Neural network |  |  |  |  |
| Li et al.<br>2021<sup>105</sup> | PET | – | Semi-automatic | ML | SVM | 50 | 25 | – | Training-Validation<br>split |
| Li et al. 2022<sup>55</sup> | PET/CT | Non-contrast CT | Manual | ML | LR | 125 | 54 | – | Training-Validation<br>split |
| Liu et al.<br>2016<sup>106</sup> | CT | Non-contrast CT | Semi-automatic | Classical<br>statistical model | – | – | – | – | – |
| Liu et al.<br>2020<sup>#39</sup> | CT | Contrast-enhanced | Semi-automatic | ML | LR | 210 | 53 | – | Training-Validation<br>split |
| Liu et al.<br>2020<sup>22</sup> | PET/CT | Non-contrast CT | Manual | ML | XGBoost | 111 | 37 | – | Training-Validation<br>split |
| Liu et al.<br>2022<sup>59</sup> | CT | Non-contrast CT | Manual | ML | LR<br>DT<br>RF | 296 | 50 | – | Training-Validation<br>split |
|  |  |  |  |  | SVM |  |  |  |  |
| Liu et al.<br>2023<sup>107</sup> | PET/CT | Non-contrast CT | Manual | ML | LR<br>DT<br>RF<br>SVM | – | – | – | 10-fold cross-validation |
| Lu et al.<br>2020<sup>#46</sup> | CT | Non-contrast CT | Manual | ML | LR | 83 | – | 21 | Training-Validation split |
| Lu et al.<br>2020<sup>108</sup> | CT | Non-contrast CT | Semi-automatic | ML | KNN<br>Bagging<br>SVM<br>RF | 105 | 228 | – | Training-Validation split |
| Lu et al.<br>2022<sup>25</sup> | CT | Non-contrast CT | Manual | ML | DT<br>AdaBoost<br>NB<br>RF<br>LR<br>SVM<br>XGBoost | 140 | 61 | – | Training-Validation split |
|  |  |  |  |  | KNN |  |  |  |  |
| Ma et al.<br>2020<sup>73</sup> | CT | Contrast-enhanced | Manual | ML | SVM | 98 | 42 | – | Training-Validation<br>split |
| Mei et al.<br>2018<sup>40</sup> | CT | Non-contrast CT | Manual | Classical<br>statistical model | – | – | – | – | – |
| Mu et al.<br>2020<sup>85</sup> | PET/CT | Non-contrast CT | Manual | DL | – | 429 | 187 | 65 | Training-Validation<br>split |
| Nair et al.<br>2021<sup>17</sup> | PET/CT | Contrast-enhanced | Manual | ML | LR | – | – | – | LOOCV |
| Ninomiya et<br>al. 2021<sup>60</sup> | CT | Contrast-enhanced | Manual | ML | SVM | 99<sup>‡</sup> | 99<sup>‡</sup> | 95 | Training-Validation<br>split |
| Ninomiya et<br>al. 2023<sup>61</sup> | CT | Contrast-enhanced | Not specified | ML | SVM | 92 | 62 | – | Training-Validation<br>split |
| Omura et al.<br>2023<sup>20</sup> | CT | Contrast-enhanced | Automatic | ML | RF | – | – | – | Training-Validation<br>split |
| Ríos Velázquez et al. 2017<sup>62</sup> | CT | Contrast + Non-contrast CT | Semi-automatic | ML | RF | 353 | 352 | – | Training-Validation split |
| Rossi et al. 2021<sup>63</sup> | CT | Non-contrast CT | Manual | ML | SVM | – | 109 | 61 | Training-Validation split |
| Ruan et al. 2022<sup>109</sup> | PET/CT | Non-contrast CT | Manual | ML | SVM | 70 | 30 | – | Training-Validation split |
| Shao et al. 2022<sup>90</sup> | CT | Non-contrast CT | Semi-automatic | DL | – | – | – | – | Training-Validation split |
| Shiri et al. 2020<sup>18</sup> | CT low dose<br>CTD<br>PET/CT | Contrast-enhanced | Manual<br>Automatic<sup>§</sup> | ML | SVM<br>KNN<br>DT<br>QDA<br>MLP<br>SGD<br>LR<br>NB<br>GNB | 82 | 68 | – | 10-fold cross-validation |
|  |  |  |  |  | RF<br>AdaBoost<br>Bagging |  |  |  |  |
| Shiri et al.<br>2022<sup>88</sup> | PET/CT | Non-contrast CT | Manual<br>Automatic<sup>§</sup> | ML | RF | – | – | – | Training-Validation<br>split |
| Song et al.<br>2021<sup>42</sup> | CT | Not specified | Manual<br>Automatic | DL<br>ML | SVM | 528 | 137 | – | Training-Validation<br>split |
| Song et al.<br>2020<sup>74</sup> | CT | Non-contrast CT | Automatic | ML | DT | 268 | 67 | – | Training-Validation<br>split |
| Trivizakis et<br>al. 2021<sup>110</sup> | CT | Not specified | Not specified | DL<br>ML | KNN<br>DT<br>RBF-GPC<br>RBF-SVM<br>Linear SVM<br>Polynomial<br>SVM<br>Sigmoid SVM | – | – | – | 5-fold cross-validation |
| Tu et al.<br>2019<sup>64</sup> | CT | Non-contrast CT | Not specified | ML | LR | 243 | 161 | – | Training-Validation<br>split |
| Wang et al.<br>2019<sup>21</sup> | CT | Contrast-enhanced | Manual | ML | SVM | 41 | – | – | Training-Validation<br>split |
| Wang et al.<br>2021<sup>43</sup> | CT | Non-contrast CT | Manual | DL | – | 882 | 125 | 255 | Training-Validation<br>split |
| Wang et al.<br>2022<sup>#65</sup> | CT | Non-contrast CT | Manual | DL<br>ML | LASSO | – | – | – | Training-Validation<br>split<sup>¶</sup> |
| Wang et al.<br>2022<sup>##47</sup> | PET/CT | Non-contrast CT | Semi-automatic | ML | LR | 180 | 78 | – | Training-Validation<br>split |
| Weng et al.<br>2021<sup>66</sup> | CT | Non-contrast CT | Semi-automatic | ML | LR | 210 | 91 | – | Training-Validation<br>split |
| Wu et al.<br>2020<sup>26</sup> | CT | Contrast-enhanced | Manual | ML | LR | – | – | – | 10-fold cross-validation |
| Xiao et al. | PET/CT | Non-contrast CT | Manual | DL | RF | 121 | 29 | – | Training-Validation |
| 2023<sup>91</sup> |  |  |  | ML |  |  |  |  | split |
| Yamazaki et al. 2022<sup>111</sup> | CT | Non-contrast CT | Semi-automatic | ML | RF | – | – | – | – |
| Yang et al. 2020<sup>#27</sup> | CT | Contrast-enhanced | Semi-automatic | ML | LASSO | 130 | 40 | – | Training-Validation split |
| Yang et al. 2020<sup>112</sup> | PET/CT | Non-contrast CT | Semi-automatic | ML | RF | 139 | 35 | – | Training-Validation split |
| Yang et al. 2022<sup>#67</sup> | CT | Contrast + Non-contrast CT | Manual | ML | LR<br>RF<br>SVM<br>GBT<br>NB | 327 | 66 | 19 | Training-Validation split |
| Yang et al. 2022<sup>23</sup> | PET/CT | Non-contrast CT | Semi-automatic | ML | SVM<br>DT<br>RF | 218 | 95 | – | Training-Validation split |
| Yang et al. | CT | Contrast-enhanced | Manual | ML | LR | 176 | 74 | – | Training-Validation |
| 2022<sup>48</sup> |  |  |  |  |  |  |  |  | split |
| Yip et al.<br>2017<sup>89</sup> | PET | – | Manual | Classical<br>statistical model | – | – | – | – | – |
| Zhang et al.<br>2018<sup>28</sup> | CT | Non-contrast CT | Manual | ML | LR | 140 | 40 | – | Training-Validation<br>split |
| Zhang et al.<br>2020<sup>#70</sup> | PET/CT | Non-contrast CT | Manual | ML | RF<br>SVM<br>LR | – | – | – | 10-fold cross-validation |
| Zhang et al.<br>2020<sup>113</sup> | PET/CT | Non-contrast CT | Semi-automatic | ML | LR | 175 | 73 | – | Training-Validation<br>split |
| Zhang et al.<br>2020<sup>##68</sup> | CT | Non-contrast CT | Semi-automatic | DL<br>ML | RF<br>SVM | 638 | 71 | 205 | Training-Validation<br>split |
| Zhang et al. | CT | Contrast-enhanced | Semi-automatic | ML | LASSO | – | – | – | Training-Validation |
| 2021<sup>41</sup> |  |  |  |  |  |  |  |  | split |
| Zhang et al.<br>2021<sup>69</sup> | CT | Non-contrast CT | Manual | ML | DT<br>LR<br>SVM<br>Multivariate<br>analysis for C-R-R model | 294 | 126 | – | Training-Validation<br>split |
| Zhang et al.<br>2023<sup>114</sup> | PET | – | Manual | ML | SVM<br>RF<br>LR<br>AdaBoost | – | – | – | 10-fold cross-validation |
| Zhao et al.<br>2019<sup>86</sup> | CT | Non-contrast CT | Manual | DL<br>ML | LR | 348 | 116 | 116 | Training-Validation<br>split |
| Zhao et al.<br>2019<sup>115</sup> | CT | Non-contrast CT | Manual | ML | LR | 322 | 315 | – | Training-Validation<br>split |
| Zhao et al.<br>2022<sup>71</sup> | PET/CT | Non-contrast CT | Semi-automatic | ML | LR | 65 | 23 | – | Training-Validation<br>split |
| Zhu et al.<br>2022<sup>32</sup> | CT | Non-contrast CT | Semi-automatic | ML | LASSO<br>RF<br>SVM | 875 | 217 | – | Training-Validation<br>split |
| Zhu et al.<br>2021<sup>29</sup> | CT | Contrast-enhanced | Manual | ML | SVM<br>KNN<br>RF<br>LR | 159 | 40 | – | Training-Validation<br>split |
| Zuo et al.<br>2023<sup>87</sup> | PET/CT | Non-contrast CT | Manual | ML | LGBM<br>XGBoost<br>RF<br>LR | 410 | 170 | 180 | Training-Validation<br>split |

\*In studies in which PET/CT was performed, only details about contrast were provided for PET acquisition. Consequently, it was assumed that CT scans were non-contrast enhanced.
†Not specified but inferred from the methodology and results.
‡Number of cases for training and validation sets not specified; only a total number for both cohorts provided.
§Manual segmentation for PET images; automatic segmentation for CT images.
¶80% Training-Validation split.
CT, computed tomography; CTD, contrast-enhanced diagnostic quality; DL, deep learning; DT, decision tree; ERBB2, v-erb-b2 avian erythroblastic leukemia viral oncogene homolog 2;
GBM, gradient boosted machine; GBT, gradient boosting tree; GNB, Gaussian Naives Bayes; GPC, Gaussian processes classification; LASSO, least absolute shrinkage and selection operator; LBP, local binary pattern; LGBM, Light gradient boosted machine; LOOCV, leave-one-out cross-validation; LR, logistic regression; ML, machine learning; MLP, multilayer perceptron; NB, Naive Bayes; KNN, K-nearest neighbors; PET, positron emission tomography; QDA, quadratic discriminant analysis; RBF, radial basis function; RF, random forest; SGD, stochastic gradient descent; SVM, support vector machine; TP53, tumor suppressor protein 53.

#### Clinical characteristics of the studies

The 89 studies evaluated in the qualitative synthesis included a total of 32,084 patients with NSCLC. Although most of the studies included >200 patients, in 42 publications, the sample size did not reach this figure and in 11 studies sample size was even lower than 100. All studies were retrospective and mostly unicentric (*n* = 72); the number of participant centers was not specified in one study^19^. In general, basic clinical and demographic information collected included sex, age, smoking status, TNM stage, histology, and treatment status at the moment of image acquisition, although this information was not available in 13, 9, 22, 34, 22 and 24 studies out of the 89 assessed, respectively. The clinical characteristics of the patients included in the 89 studies are depicted in **Table 2**. The median [range]/ mean ± standard deviation (SD) age of patients was 61.78 [59–64.17] years and 61.71 ± 3.64 years, respectively. In terms of sex, the total population was balanced, with 13,574 females and 14,066 males. The smoking history was available for 23,200 patients, and many were non-smokers (*n* = 12,813); while smoking history was unknown for 1,146 patients. Out of the 55 studies detailing information about the TNM stage, the majority of them (*n* = 40) included information about the four stages (I-IV), either provided per group or grouped in stages I-II and stages III-IV. Among the 15 studies that did not include patients of all stages, two studies included only early stage patients (stages I and II)^20, 21^, two included patients stage II-IV^22, 23^, six included only patients of stages III and IV^24-29^ (three of them with a majority of stage IV patients^24, 27, 29^), and five included patients of stages I-III without including the most advanced stage^19, 30-33^. A total of 65 studies included patients with adenocarcinoma: 43 exclusively including this histology subtype and 22 including other NSCLC histology types as well. Finally, in most of the cases (*n* = 65), images were acquired before patients received any treatment, with two studies also including post-treatment images^34, 35^. In 24 studies, no information on treatment was detailed, although in some of them image acquisition before surgery^31, 36-41^, before polymerase chain reaction (PCR)^42^, or before pathological diagnosis^43^ was detailed as an inclusion criterion. In five studies^19, 44-47^, authors specify that patients had not received radiotherapy or chemotherapy, but no information on targeted therapy was provided. Finally, only one study^48^ out of the 89 included in the systematic review, which did not meet the inclusion criteria to be considered for the meta-analysis, included patients who had received treatment with tyrosine kinase inhibitors (TKIs).

**Table 2.**
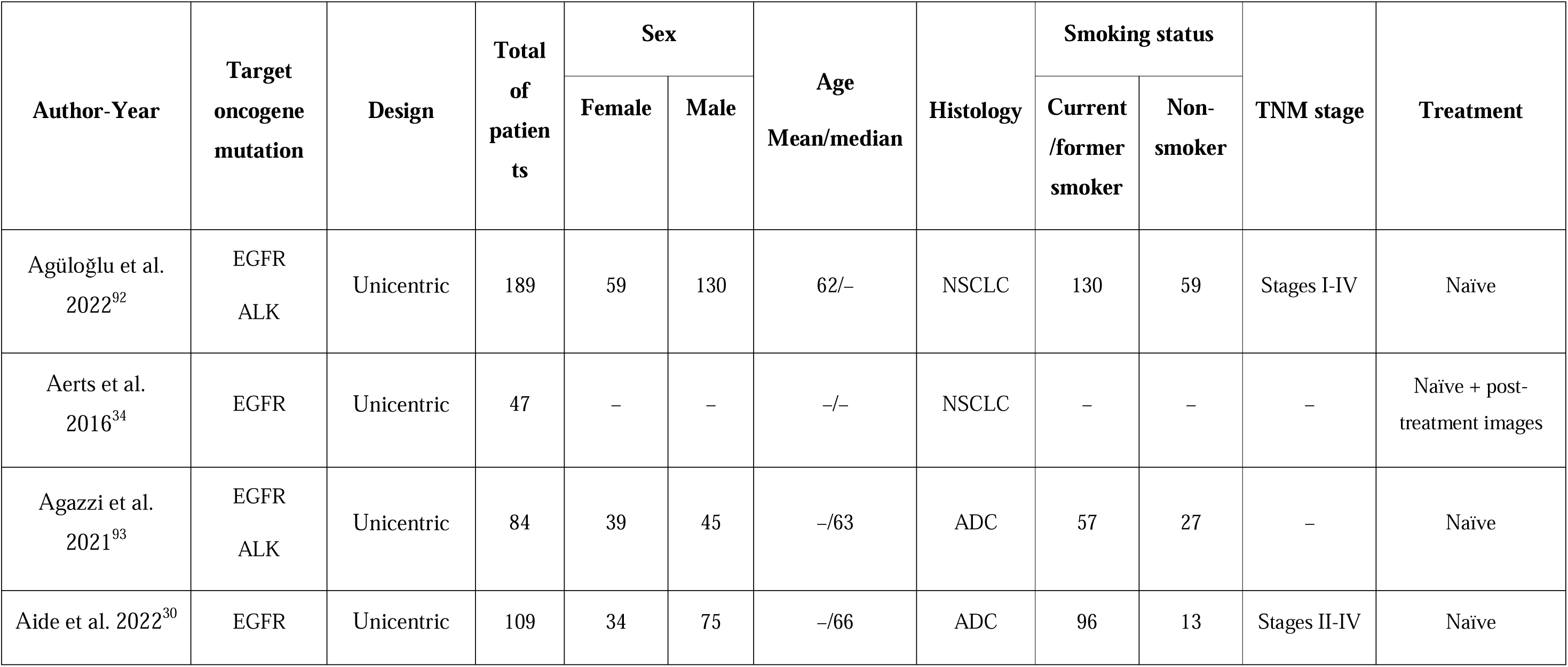
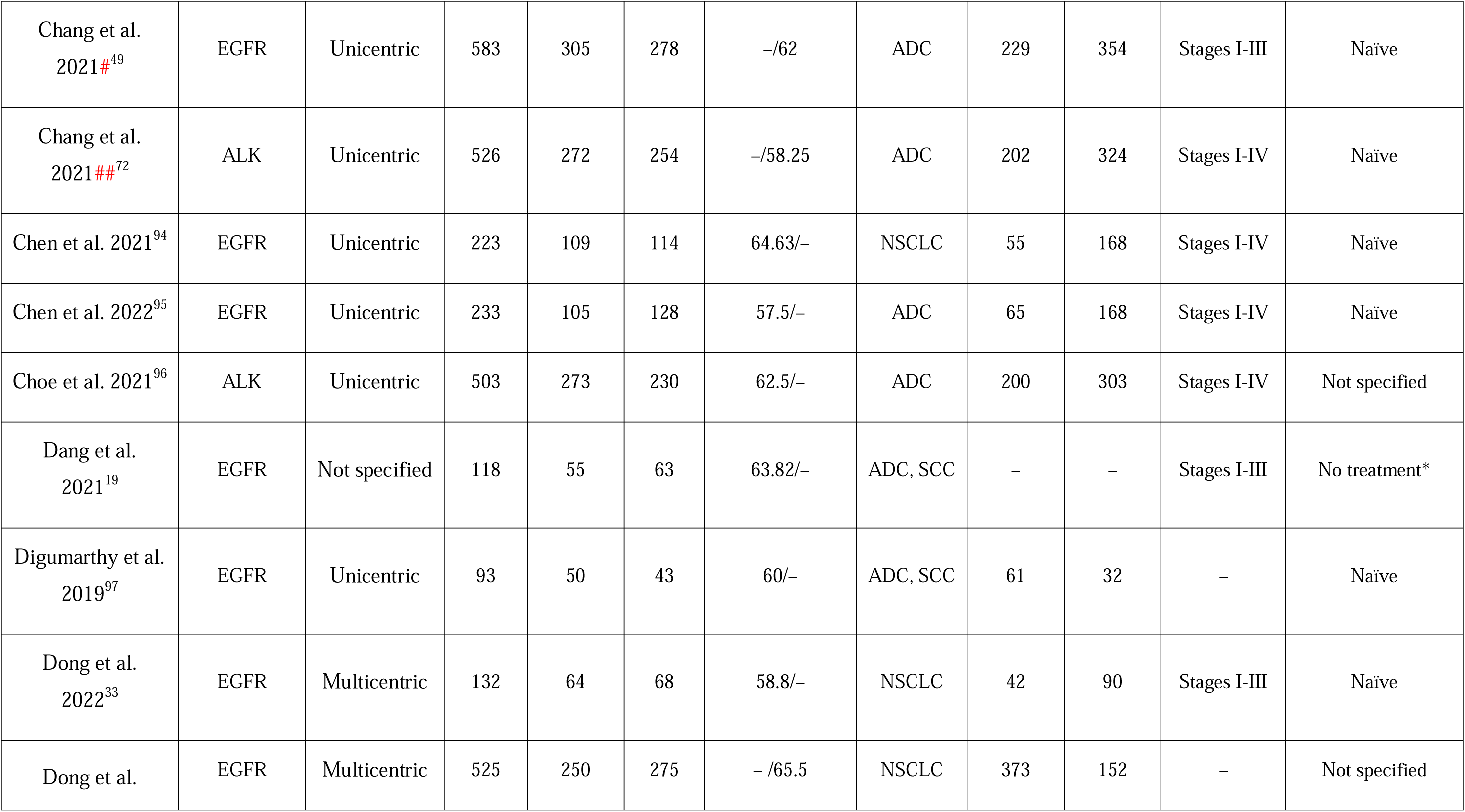
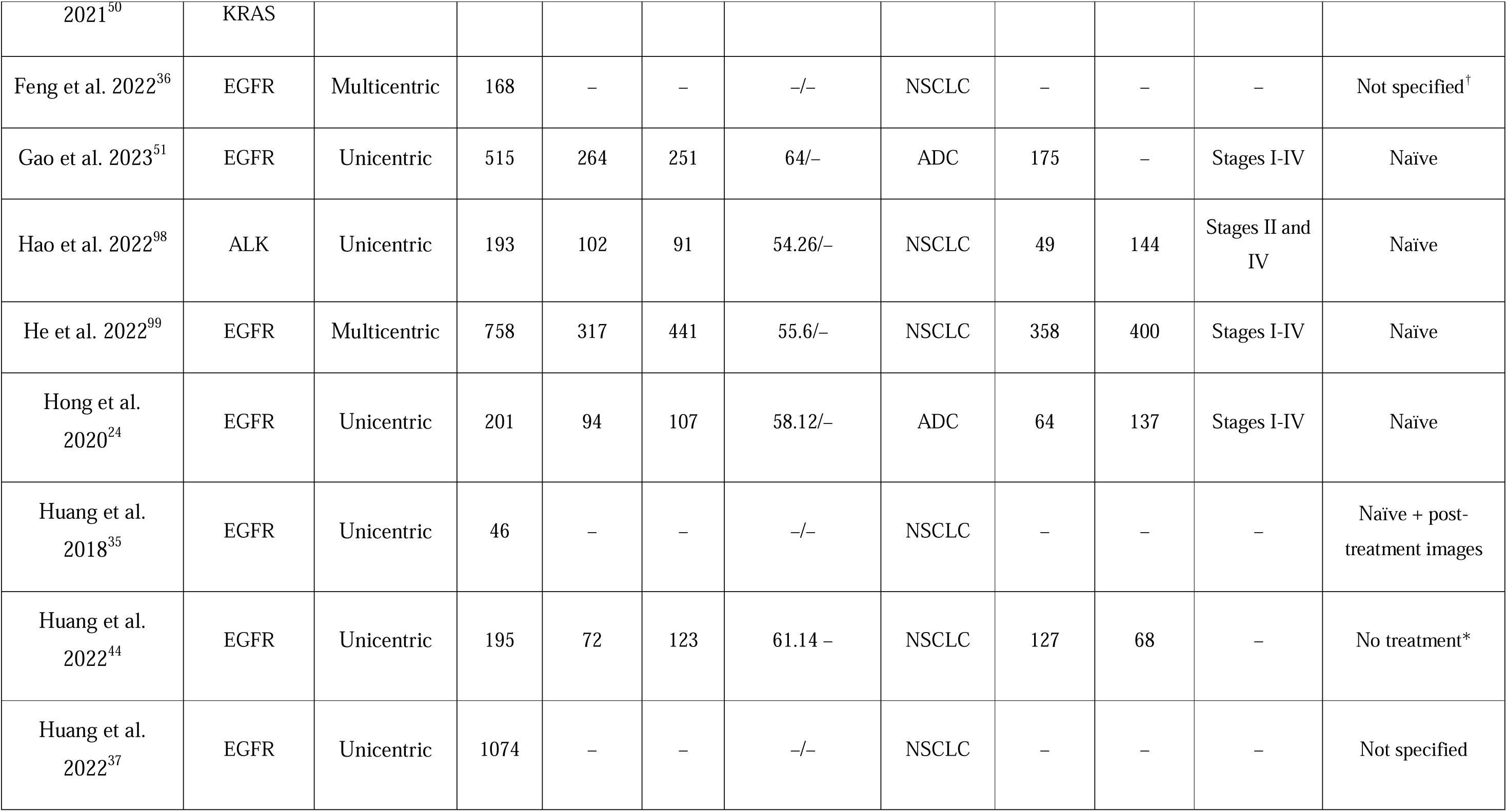
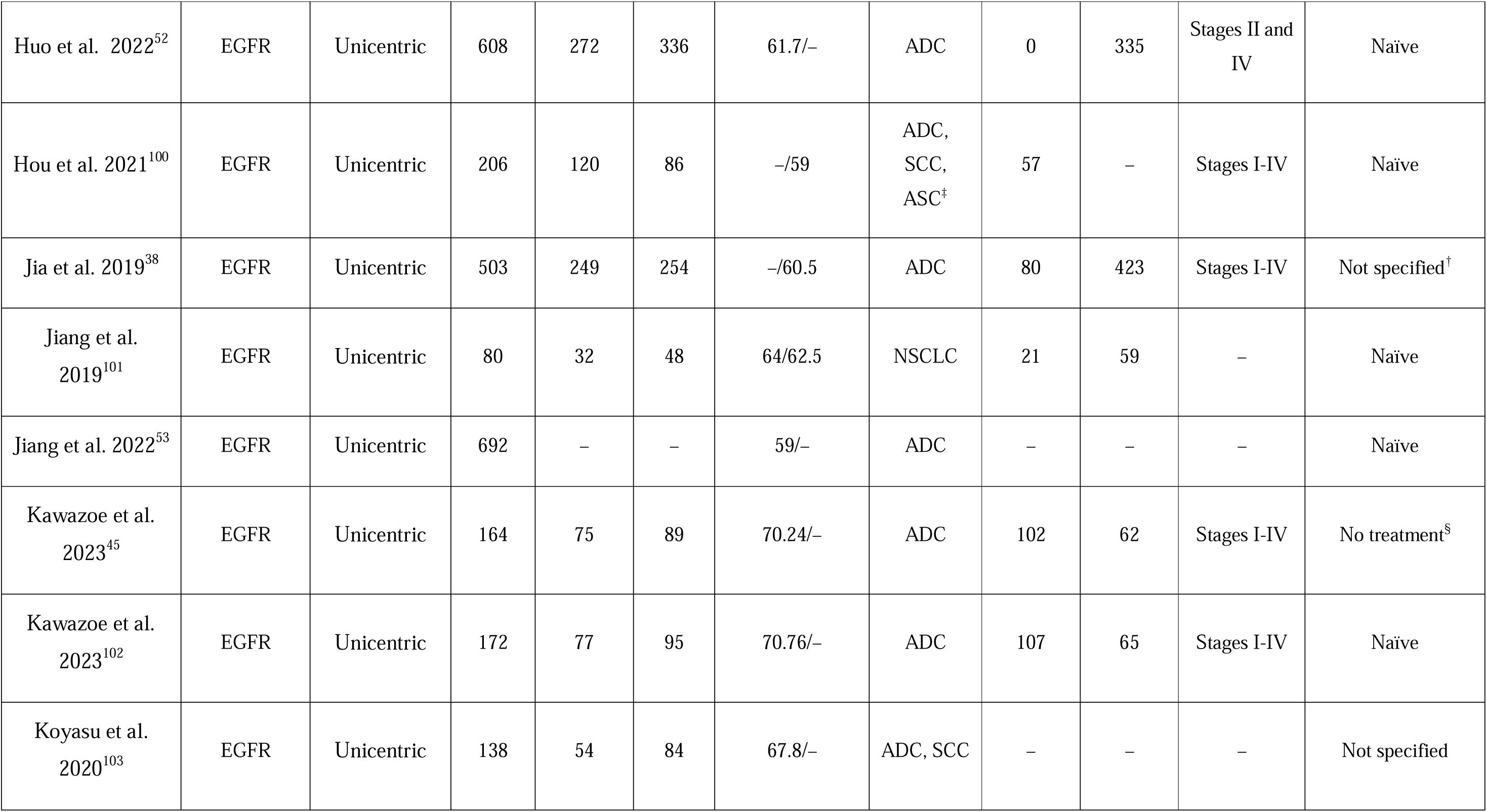
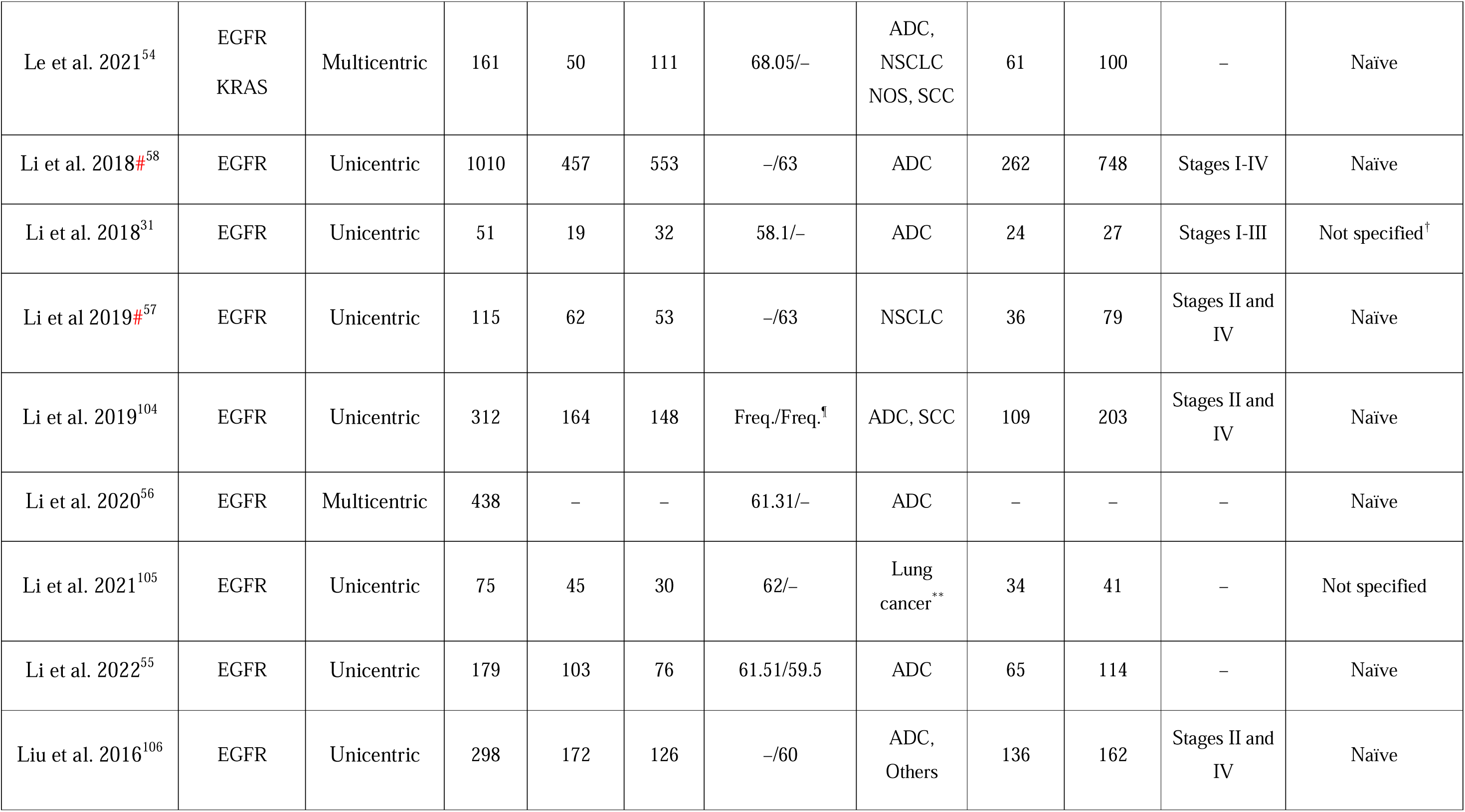
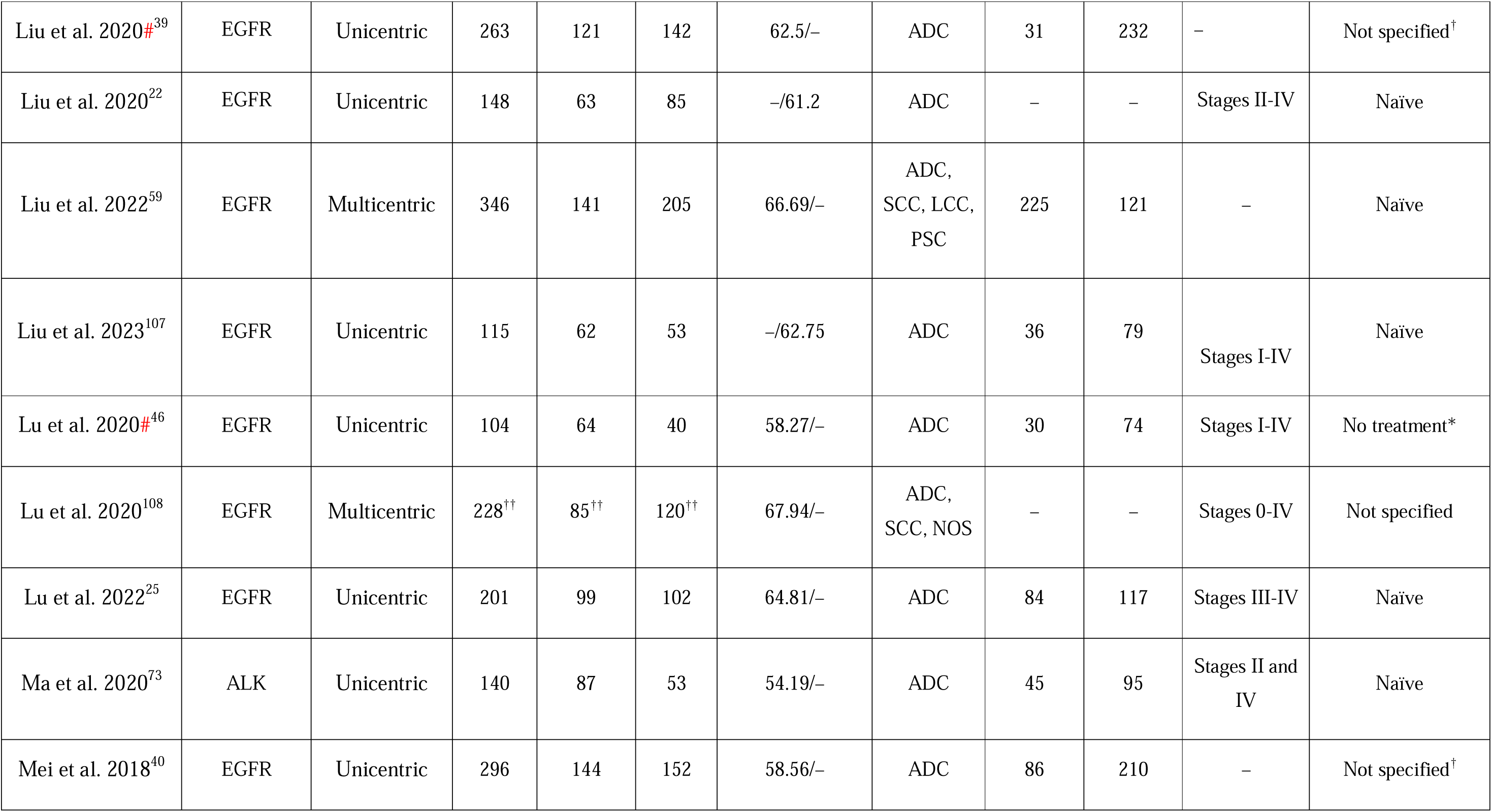
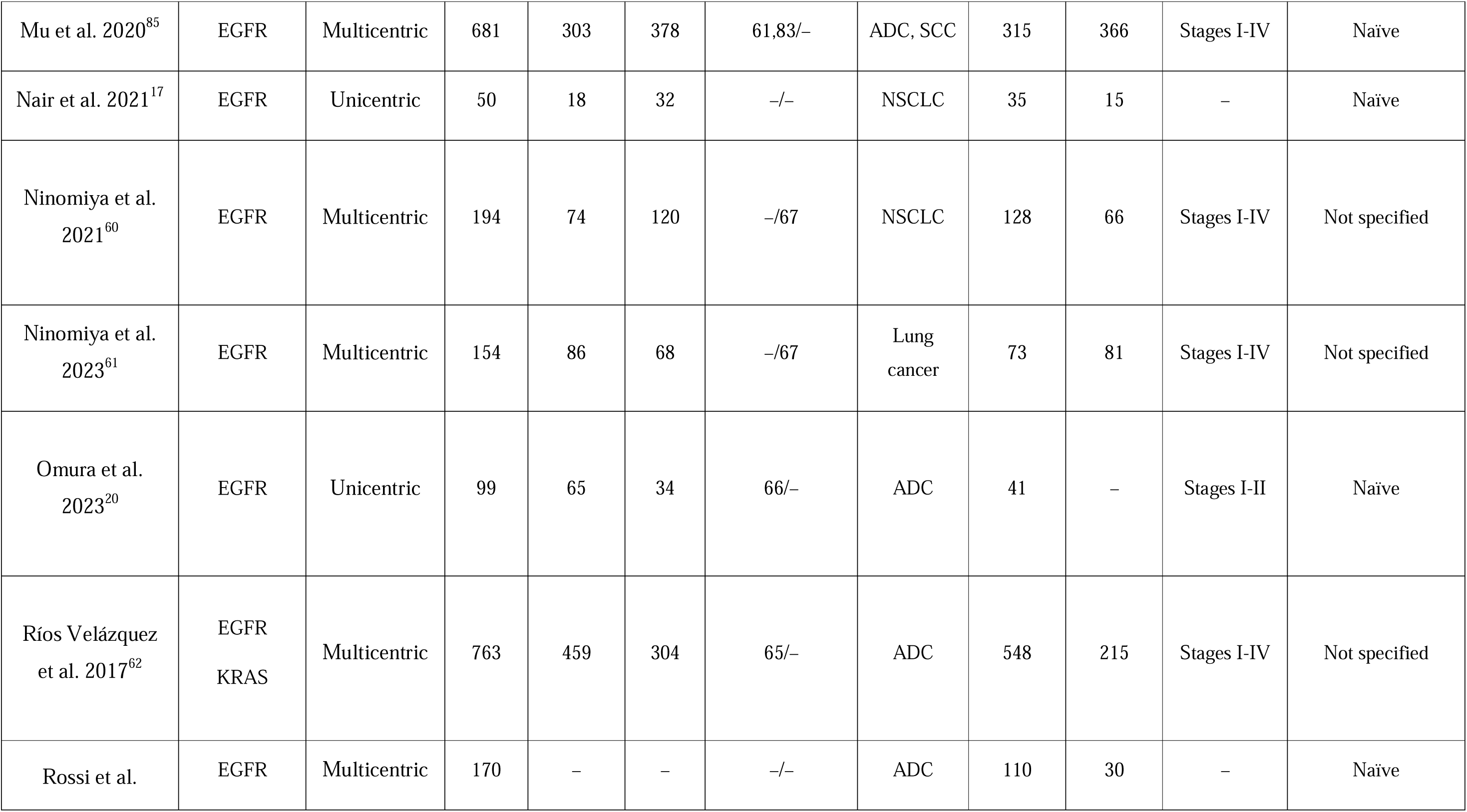
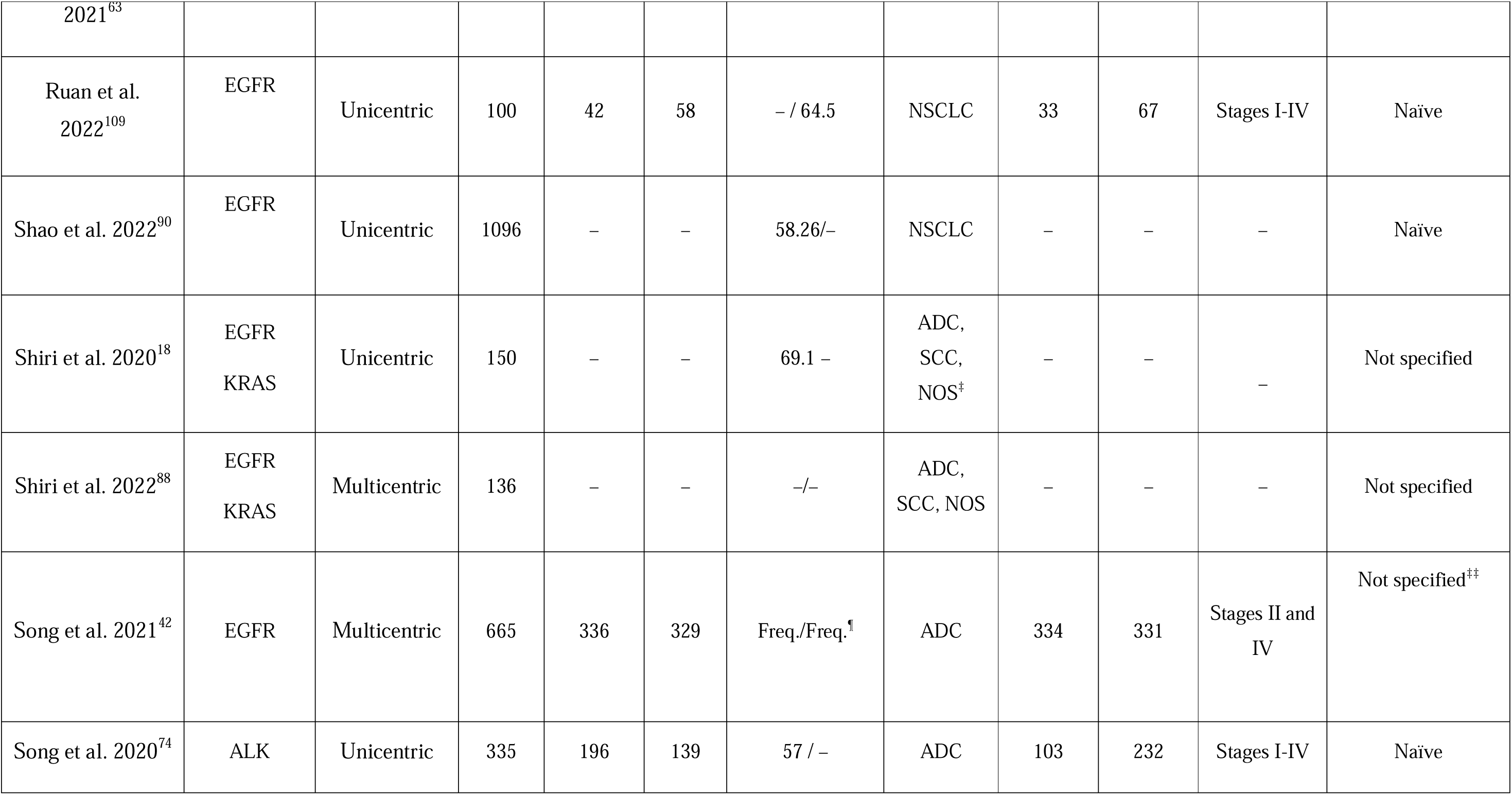
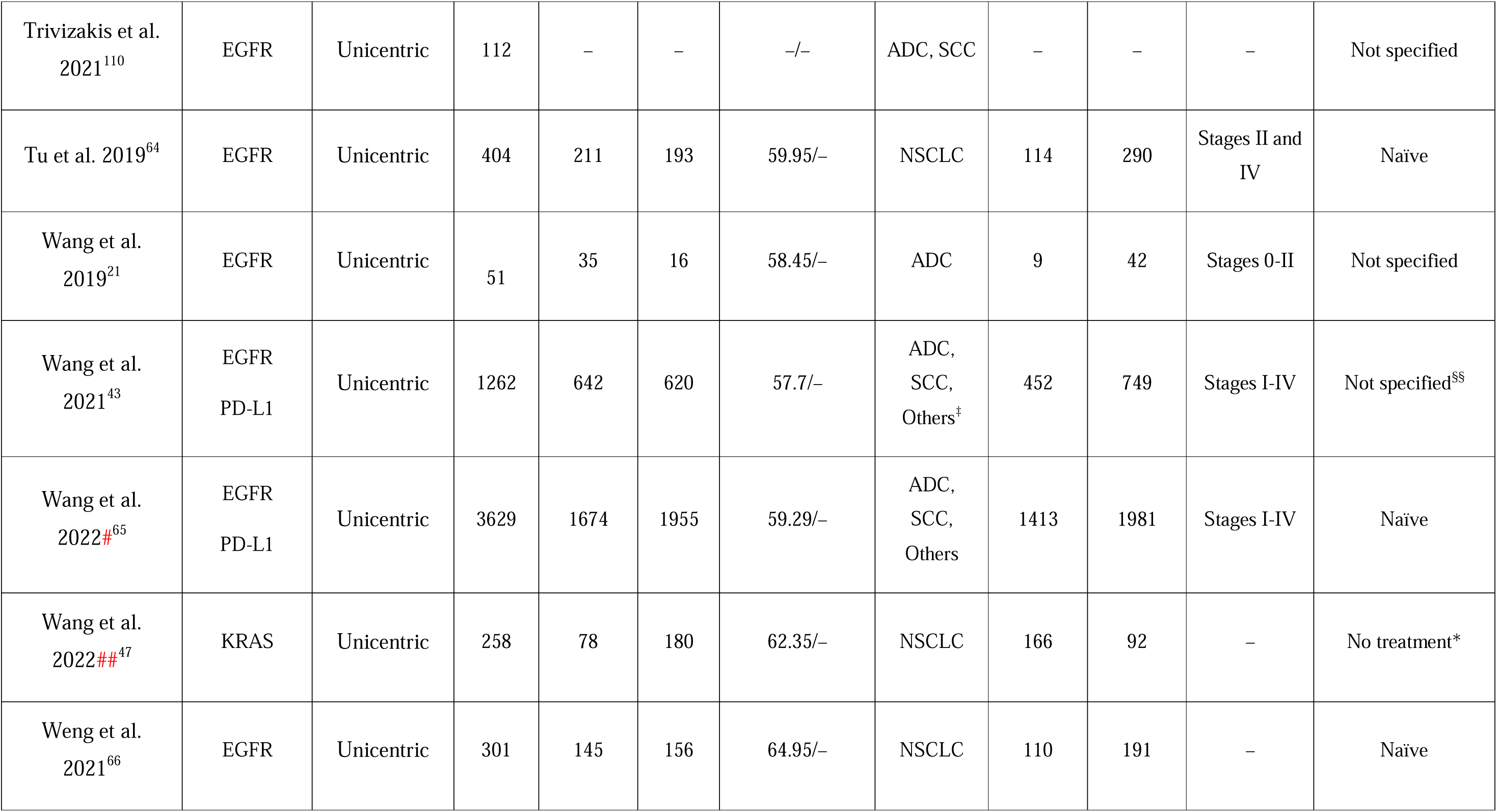
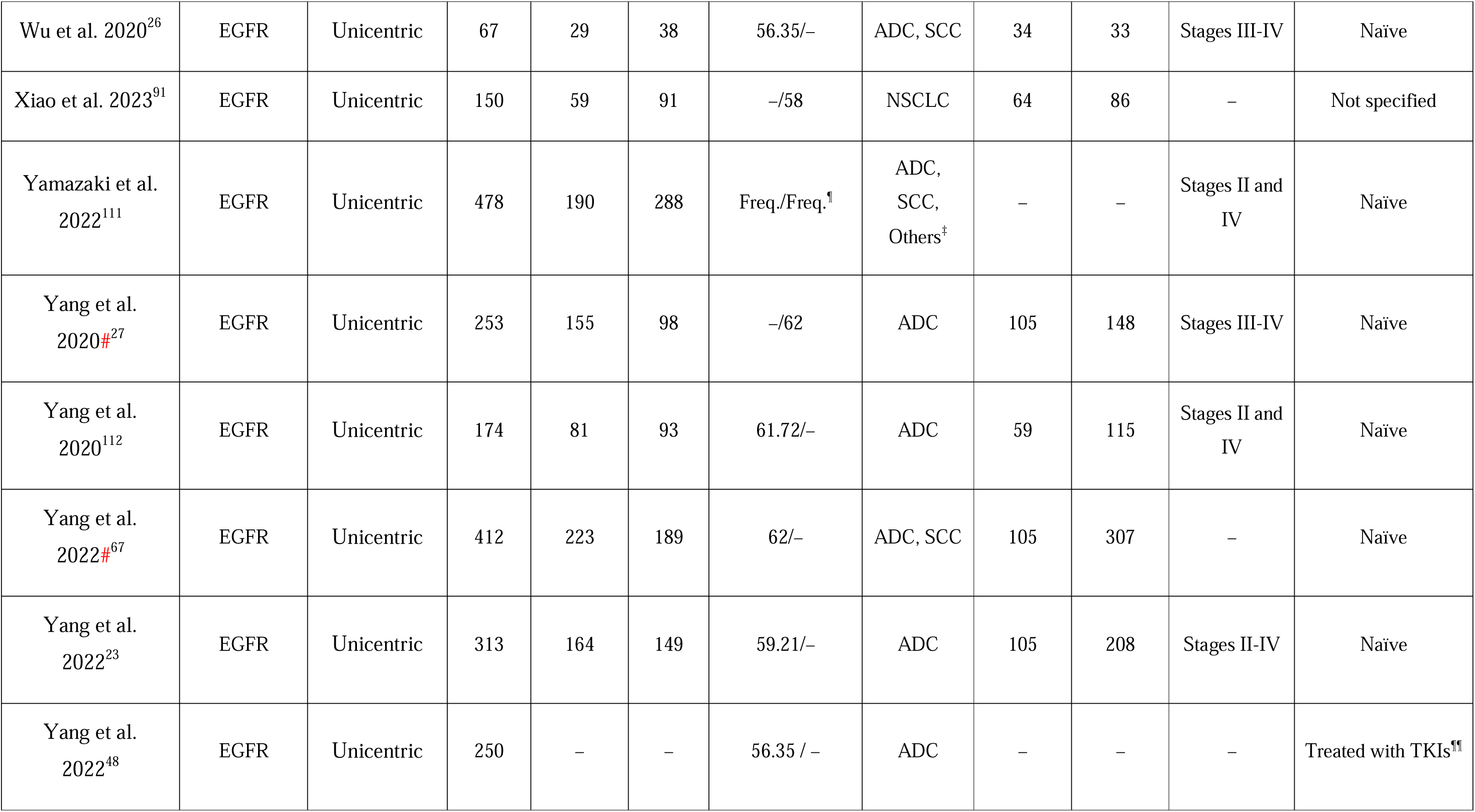
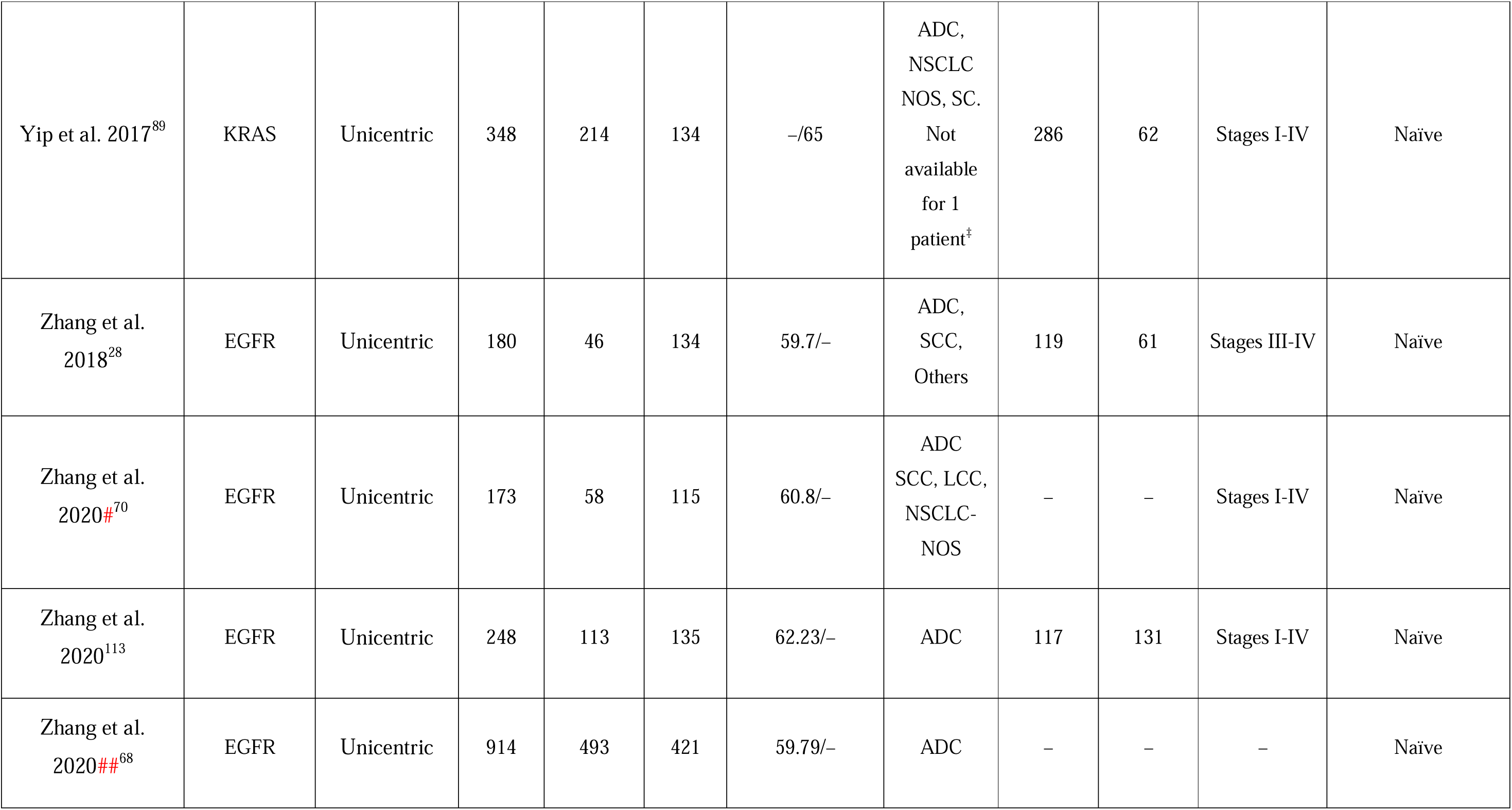
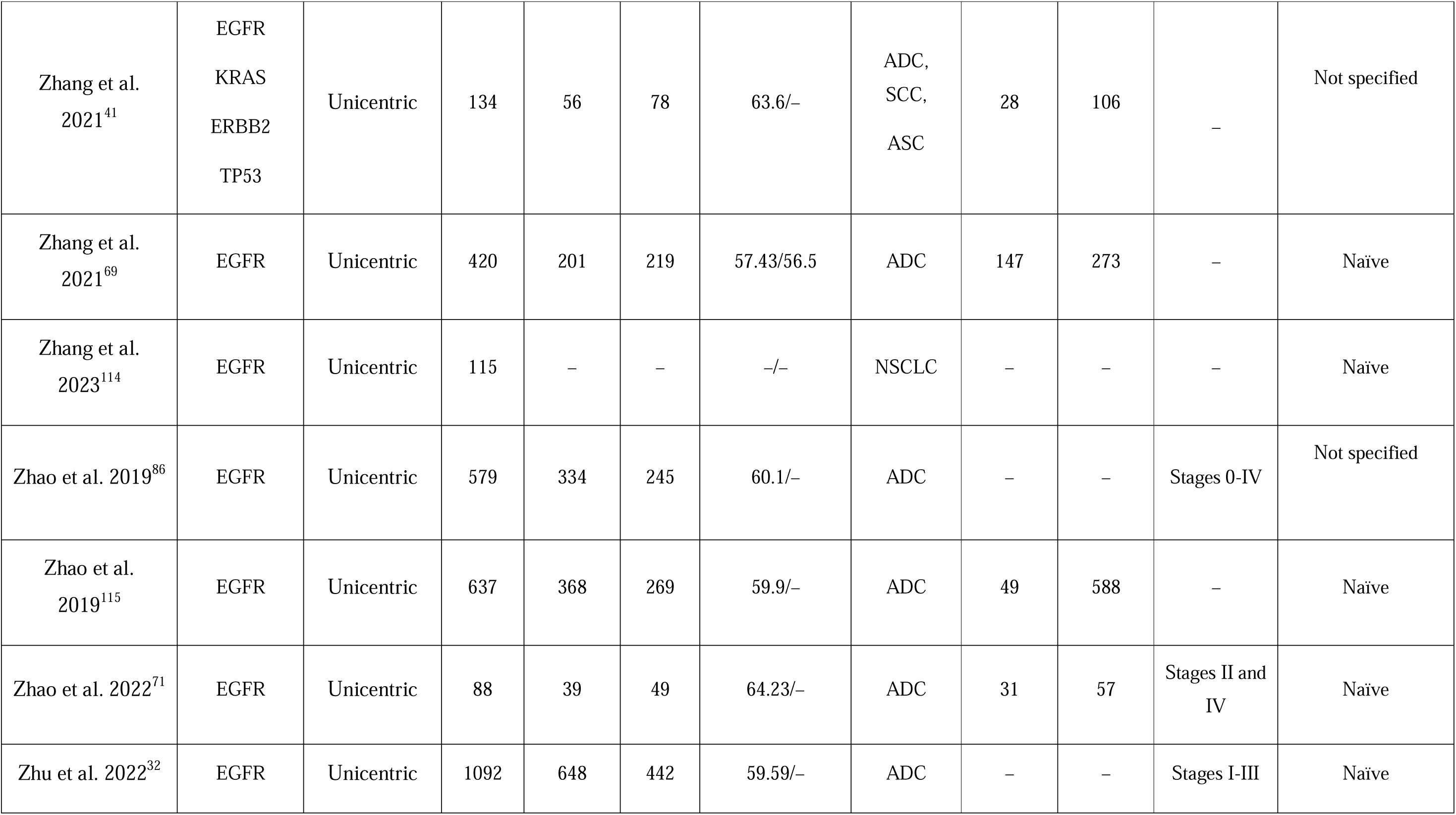
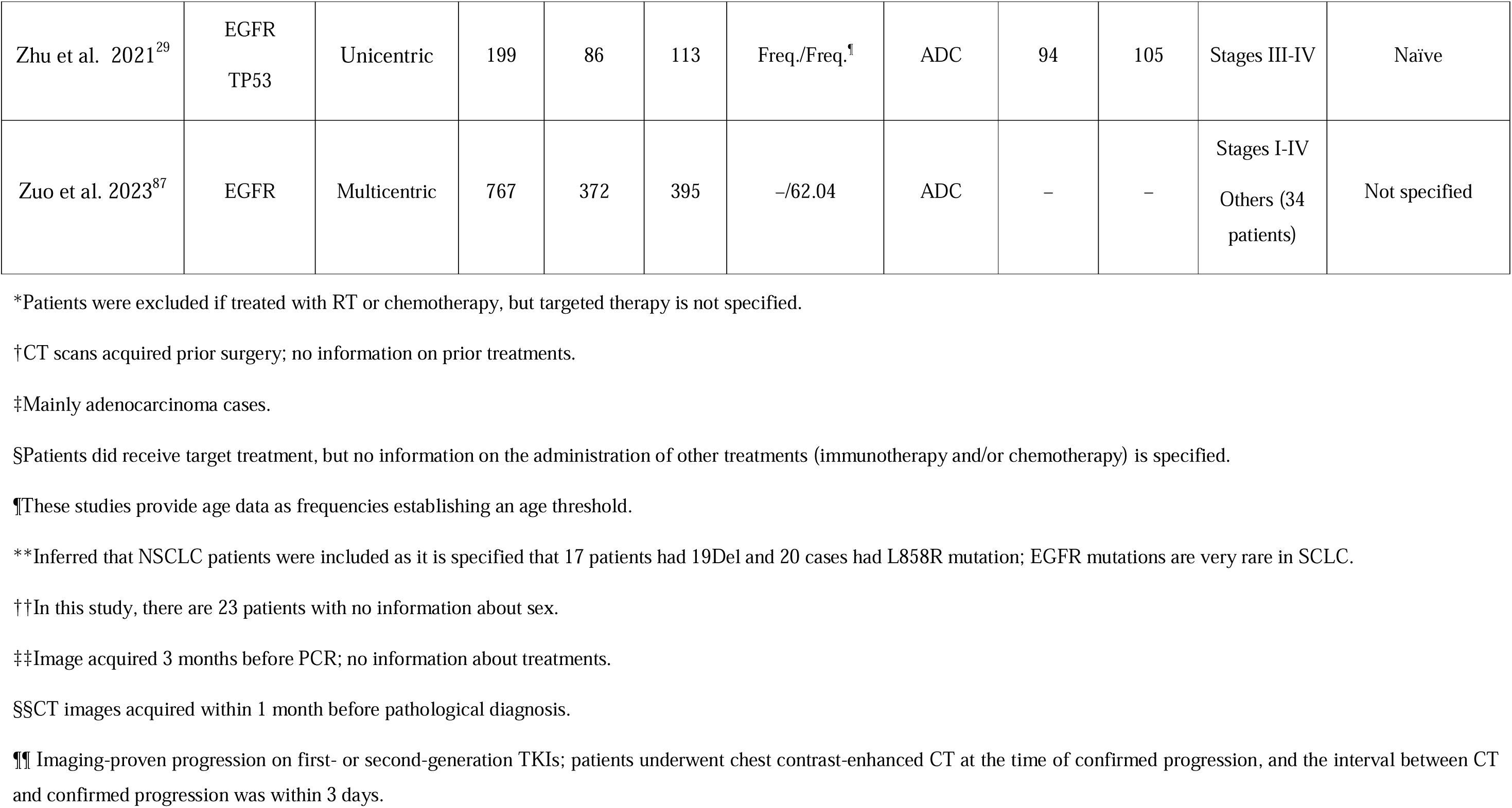
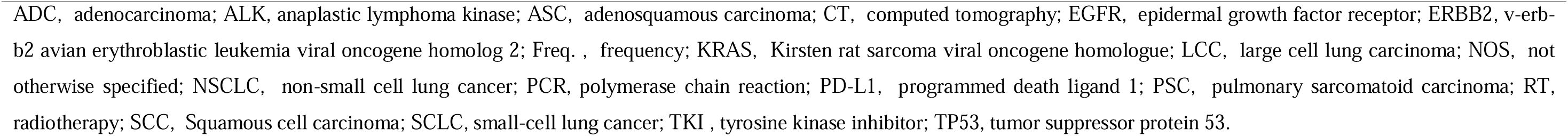
Clinical characteristics of the studies (*N* = 89) included in the systematic review. For those studies with the same name for the first author and published the same year, a hashtag was added to unequivocally indicate those that were included in the different meta-analyses and consequently, that are represented in the forest plots.

### Quantitative analysis (meta-analysis)

A total of 38 studies met the inclusion criteria for the quantitative assessment (*n* = 17,066 patients). Three main different meta-analyses including radiomics-based models were conducted: 1) a meta-analysis including studies focused on the detection of EGFR (*n* = 34 studies)^17, 25-28, 32, 33, 36, 38, 39, 46, 49-71^; 2) a meta-analysis including studies focused on the detection of ALK (*n* = 3 studies)^72-74^; 3) a meta-analysis including studies focused on the detection of KRAS (*n* = 4 studies)^47, 50, 54, 62^. In three studies, authors developed models for the detection of both EGFR and KRAS^50, 54, 62^. Furthermore, a separate meta-analysis was conducted for combined models (radiomics features + clinical variables) for the prediction of EGFR (not enough studies for ALK or KRAS mutations). Studies included in all the meta-analyses conducted are summarized in **Supplementary Table S3.** Details on the radiomics features included in the EGFR, ALK, and KRAS models are summarized in **Supplementary Table S4**, **Supplementary Table S5** and **Supplementary Table S6**, respectively. In terms of radiomics variables, models grouped different combinations of first order, shape, gray level co-occurrence matrix (GLCM), gray level size zone matrix (GLSZM), gray level run length matrix (GLRLM), neighboring gray tone difference matrix (NGTDM), and gray level dependence matrix (GLDM) features. Clinical data included sex, smoking history, and/or histological type in the majority of studies.

#### EGFR

Results of the meta-analysis focused on models built with radiomics features are summarized in **Figure 2**. Note that this meta-analysis also included a study^50^ in which predictions were based on features extracted by a multi-channel and multi-task deep learning model with the ability to simultaneously detect EGFR and KRAS oncogene mutations; and consequently, did not include radiomics features (only single-task results for the independent prediction of EGFR and KRAS were considered for the quantitative analysis). A hierarchical sROC curve was plotted for the included 24 studies ^17, 25, 27, 32, 36, 39, 46, 49-51, 54, 55, 57-60, 62, 64-66, 68-71^ that evaluate the performance of AI algorithms in predicting EGFR mutation status in NSCLC (**Supplementary Figure S1**). Eight studies assessed more than one model^32, 36, 39, 50, 59, 60, 65, 70^. As observed, radiomics-based models exhibited high diagnostic performance in predicting EGFR mutation status with an overall AUC of 0.766. The AI algorithms’ sensitivity in determining the EGFR mutation status varied from 0.362 to 0.948, resulting in an estimate of 0.753 (95% CI 0.721–0.783). The FPR of these algorithms ranged from 0.022 to 0.761, with a estimate of 0.346 (95% CI 0.305–0.390). Detecting a positive case for EGFR mutation was almost six times more likely than not detecting it (DOR = 5.70 [95% CI 4.74–6.81]).

**Figure 2.**
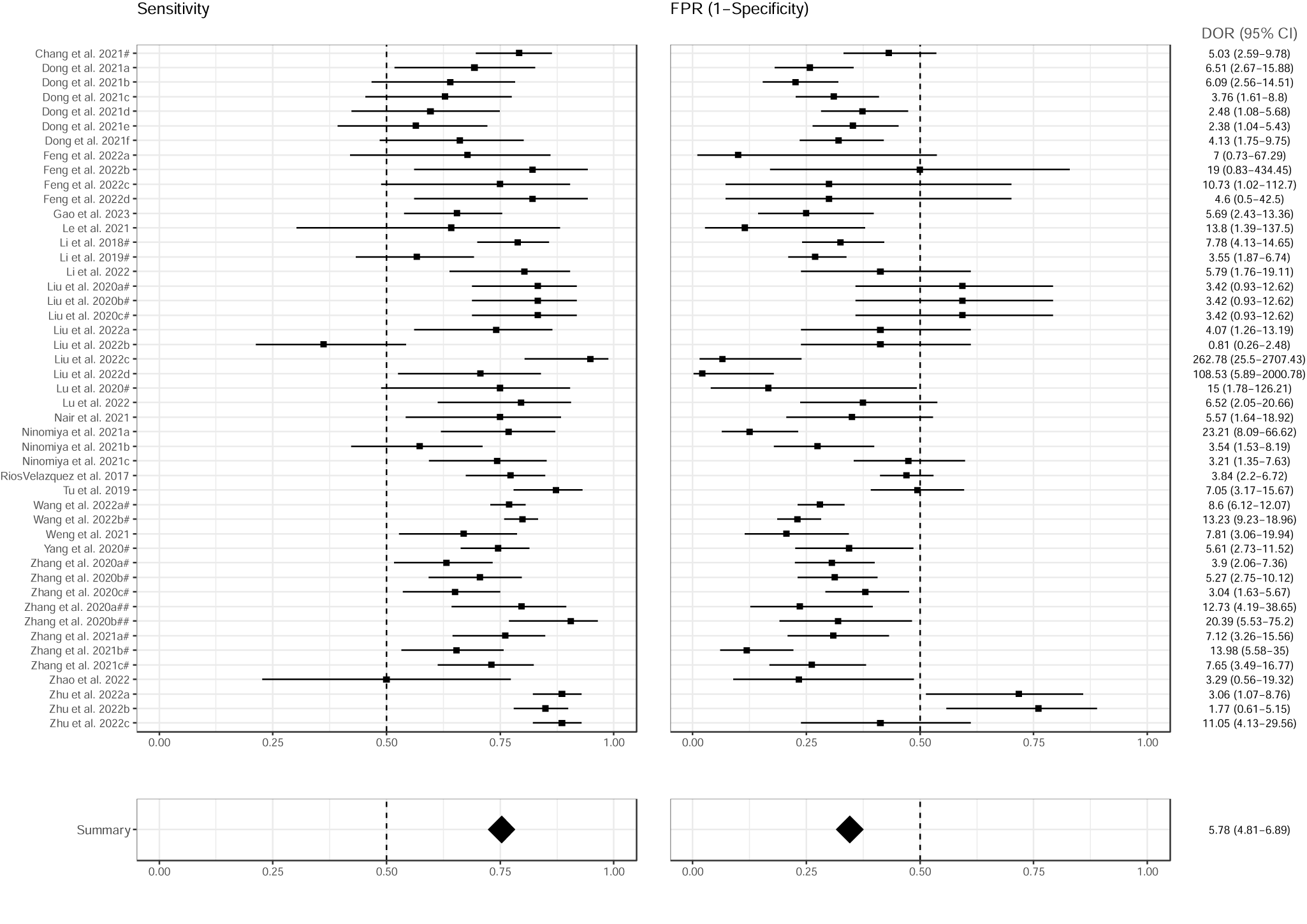
Forest plots of the included studies developing radiomics models using machine learning and/or deep learning methods for the prediction of EGFR mutation status. Numbers are estimated with 95% CIs in brackets and indicated by horizontal lines. For those studies with the same name for the first author and published the same year, a hashtag was added to unequivocally tag them as done in Tables 1 and 2 and in the reference list. EGFR, epidermal growth factor receptor; CI, confidence interval; DOR, diagnostic odds ration; FPR, false positive rate.

The effect of adding clinical variables to radiomics models or to models including both radiomics and deep features^65^ (models including clinical data and radiomic or deep features referred in this work as combined models) in the prediction of EGFR mutation was also analyzed. This meta-analysis included 23 studies^25, 26, 28, 32, 33, 38, 39, 46, 51-53, 56-58, 61-69^, of which four of them developed more than one model^32, 39, 56, 67^. Results are depicted in **Figure 3** and sROC curve in **Supplementary Figure S1**. Overall, the performance of combined models slightly improved compared to radiomics models, with an AUC of 0.811 and a sensitivity of 0.800 (95% CI 0.767–0.830; model’s sensitivity ranging from 0.523 to 0.944). The FPR resulted similar with a value of 0.335 (95% CI 0.279–0.396; model’s FPR ranging from 0.167 to 0.760.). Detecting a positive case for EGFR mutation with combined models was more than eight times more likely than not detecting it (DOR = 8.35 [95% CI 6.77–10.20]).

**Figure 3.**
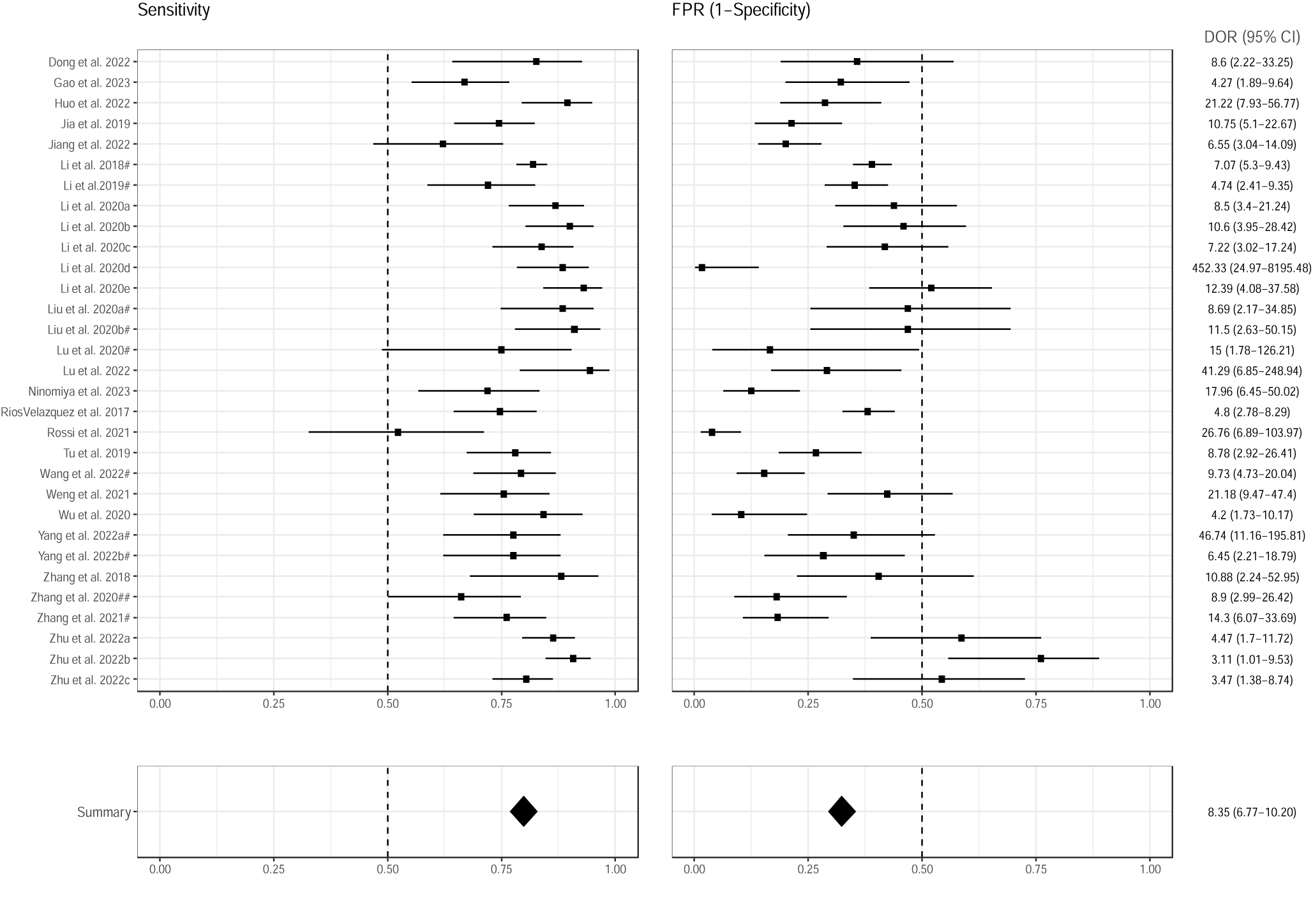
Forest plots of the included studies developing combined models (radiomics + clinical data) using machine learning and/or deep learning methods for the prediction of EGFR mutation status. Numbers are estimates with 95% CIs in brackets and indicated by horizontal lines. For those studies with the same name for the first author and published the same year, a hashtag was added to unequivocally tag them as done in Tables 1 and 2 and in the reference list. EGFR, epidermal growth factor receptor; CI, confidence interval; DOR, diagnostic odds ration; FPR, false positive rate.

#### ALK

The meta-analysis focused on radiomics-based models included three studies^72-74^, one of which developed two different models, one based on pre-contrast images and another one on post-contrast images^73^. An overall AUC of 0.831 was obtained for the prediction of ALK aberration, with a sensitivity ranging from 0.682 to 0.825, resulting in an estimate of 0.754 (95% CI 0.639–0.841). The FPR of these algorithms ranged from 0.167 to 0.277, with an estimate of 0.225 (95% CI 0.163–0.302). Detecting a positive case for ALK aberration was 11 times more likely than not detecting it (DOR = 5.70 [95% CI 5.83–19.10]) (**Figure 4** and **Supplementary Figure S2**). Given the lack of enough studies developing combined models, a meta-analysis to assess the effects of adding clinical variables in the prediction of ALK aberration was not possible. The only study^74^ that developed a model including age, sex, smoking history, smoking index, clinical stage, distal metastasis and pathological invasiveness of the tumor in combination with conventional CT features and different first order, GLCM, GLSZM, and GLRL radiomics features demonstrated increased performance in predicting ALK aberration of the combined model vs the radiomics-based model, but only in the primary cohort (AUC, 0.83–0.88, *p* = 0.01), not in the testing cohort (AUC, 0.80–0.88, *p* = 0.29).

**Figure 4.**
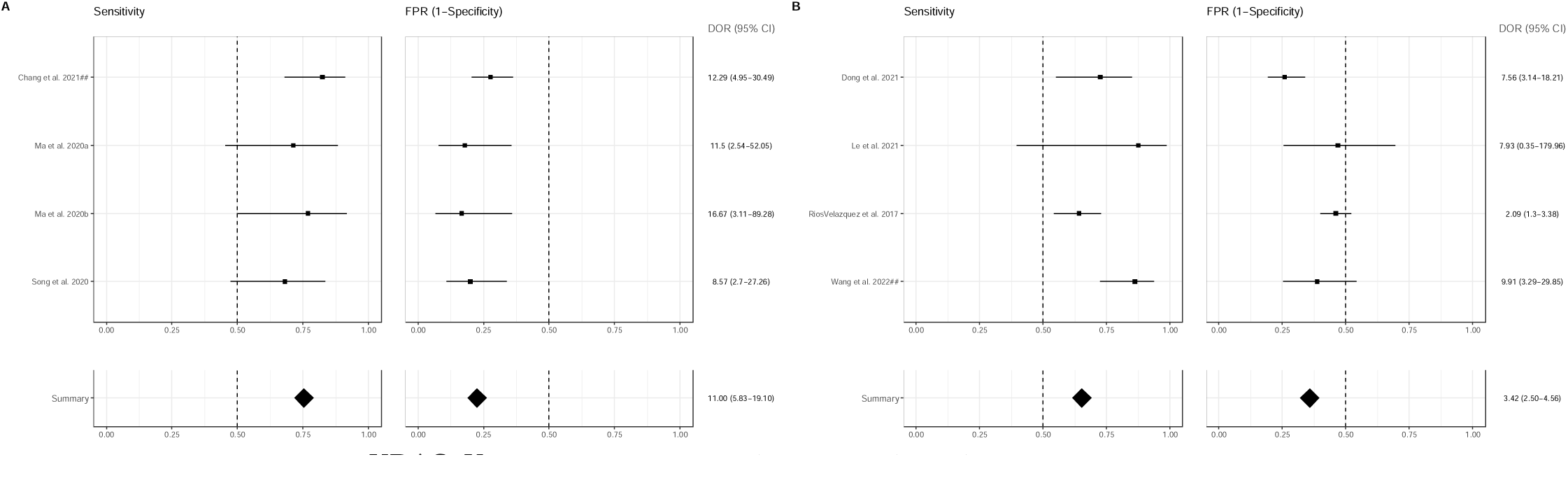
Forest plots of the included studies developing radiomics models using machine learning and/or deep learning methods for the prediction of **A)** ALK and **B)** KRAS mutation status. Numbers are estimates with 95% CIs in brackets and indicated by horizontal lines. For those studies with the same name for the first author and published the same year, a hashtag was added to unequivocally tag them as done in Tables 1 and 2 and in the reference list. ALK, anaplastic lymphoma kinase; CI, confidence interval; DOR, diagnostic odds ration; FPR, false positive rate; KRAS, Kirsten rat sarcoma viral oncogene homologue.

#### KRAS

Four studies met the inclusion criteria for the meta-analysis assessing models for KRAS mutation prediction^47, 50, 54, 62^, among which, three of them also developed models for EGFR mutation prediction^50, 54, 62^. KRAS/EGFR models were independently built except in one study, in which a multi-channel multi-task DL model for the prediction of both KRAS and EGFR mutations was developed^50^. However, and according to the inclusion criteria, only single-task metrics were considered for the quantitative analysis despite the multi-channel version displayed the highest performance for the simultaneous detection of both oncogenic driver mutations. Results of the meta-analysis evaluating radiomics-based models are shown in **Figure 4** and **Supplementary Figure S3**. KRAS mutation was predicted with an overall AUC of 0.732 and a sensitivity of 0.744 (95% CI 0.605–0.846; model’s sensitivity ranging from 0.641 to 0.875. The FPR was 0.376 (95% CI 0.274–0.491; model’s FPR ranging from 0.259 to 0.468). Detecting a positive case for KRAS mutation with radiomics-based models was more than five times more likely than not detecting it (DOR = 8.35 [95% CI 1.98–11.70]). Like ALK, the lack of enough KRAS studies made it impossible to perform a meta-analysis analyzing combined models. Only Ríos Velázquez et al.^62^ built a model including age, sex, smoking status, race, and clinical stage together with radiomics features that performed similar to the radiomics model (AUC = 0.69 [95% CI: 0.63–0.75] vs AUC = 0.63 [95% CI: 0.57–0.69]) and worse than a model developed only with clinical data AUC = 0.75 [95% CI: 0.69–0.80].

#### Meta-regression and subgroup analysis

The possible effects of different predictors on the predictive performance of the models was evaluated for EGFR mutation (not enough studies were available for ALK or KRAS mutations). Neither age, nor the use of contrast, nor the type of segmentation (manual/semi-automatic/automatic), nor the model (radiomics/combined), nor the AI methodology (machine learning/deep learning), yielded statistically significant results (**Supplementary Table S7**).

## DISCUSSION

At present, molecular testing performed on biopsied tissue remains the gold standard for diagnosis and genotyping in advanced NSCLC^75, 76^. However, given the associated limitations and inconveniences, such as the lack of enough tissue for successful testing^77, 78^, or the long turnaround times^76^, there is a need to validate and incorporate new procedures into routine clinical practice. In recent years, liquid biopsy has emerged as a promising alternative in NSCLC, especially in clinical scenarios^78^. Likewise, radiomics have shown encouraging results in prognosis and prediction in this setting^79^. In general, both methodologies possess great potential, since they are both simple, straightforward to do, and repeatable at patient follow-up visits, which makes it possible to gather important data about the type of tumor, its aggressiveness, its progression, and its response to therapy^80^. Radiomics has the additional advantage of only requiring medical images and capturing patient-level and tissue-level heterogeneity, such as CT scans in lung cancer, that are usually acquired as part of the patient’s standard journey, representing an affordable methodology both in terms of resources and costs. It is important that new techniques are properly validated to facilitate their standardization, prior to incorporation into the routine clinical workflow.

To our knowledge, this is the first systematic review and meta-analysis that analyzes the performance and applicability of different imaging-based models for the prediction of three of the most common oncogene mutations—EGFR, ALK and KRAS—in NSCLC from a clinical perspective and with a special focus on AI methodologies. So far, results were only available for EGFR studies and did not take clinical aspects into account^81^. Thus, the results of our different meta-analyses demonstrate that AI-based models developed with CT-derived radiomics features showed good performance in predicting EGFR, ALK, and KRAS mutations with a sensitivity of 0.753 [95% CI (0.721–0.783)], 0.754 [95% CI (0.639–0.841)] and 0.744 [95% CI (0.605–0.846)], respectively. Whether the inclusion of clinical variables increase models’ performance cannot be concluded from our results, although we believe that increasing the number of studies would probably confirm the trends observed in our quantitative analysis of EGFR mutation.

Our outcomes point to radiomics as a candidate screening tool for oncogene mutation status determination. We especially focused on CT-based models, aiming to obtain conclusions as applicable as possible to the standard clinical workflow since CT remains the most utilized imaging tool in NSCLC^82^. From our work, we conclude that in addition to additional validation of our findings that future studies should be conducted that consider the following important aspects. Firstly, a minimum sample size should be guaranteed to ensure the reliability of the results obtained with AI-based models^83, 84^. In both our systematic review and meta-analyses, more than half of the studies were conducted in >200 patients (*n* = 46/89 and *n* = 23/38 [*n* = 20/34 for EGFR, *n* = 2/3 for ALK and *n* = 3/4 for KRAS]), but still a sizable number had small sample sizes, which definitely limited the relevance of their conclusions. Multicentric designs would be also desirable to get more solid conclusions, an approach that few studies followed (*n* = 16/89 in the systematic review and *n* = 10/39 in the meta-analysis [*n* = 10/34 for EGFR, *n* = 0/3 for ALK and *n* = 3/4 for KRAS]). Secondly, including independent cohorts for external validations would reinforce the results, leading to more robust and reproducible models. Out of the 89 studies included in the qualitative analysis, only 9 used external cohorts for validation^43, 46, 60, 63, 67, 68, 85-87^, of which five were included in the EGFR meta-analysis^46, 60, 63, 67, 68^. Finally, it is important that patient populations reflect clinical practice. Thus, considering the potential applicability of the models for diagnostic purposes, studies should be conducted in treatment naïve populations to avoid possible therapy-related confounding effects, an inclusion criterion mostly applied in the studies evaluated in this work, but still missing in some of them. Additionally, studies should be carried out preferably in stage III-IV NSCLC patients (especially in those at stage IV, for whom clinical guidelines recommend molecular testing^75, 76^). As demonstrated in this work, most of the studies published so far do not provide information on TNM stage or include patients from all stages. Despite the heterogeneity of the studies evaluated, we believe that the evidence provided is enough as to demonstrate the potential of radiomics in oncogene mutation status determination. Thus, AI-based models using radiomics extracted from CT scans could be effective non-invasive screening tools to detect targetable driver mutations in NSCLC with good sensitivity and moderate specificity. These tools would not be intended to replace gold standard techniques, such as PCR or next-generation sequencing, but to allow for the potential earlier identification of ideal candidates to be genetically tested, saving time, costs, and samples. Consequently, a high sensitivity would ensure the identification of oncogene mutation positive patients for whom laboratory-based testing would be subsequently confirmed.

When the influence of different factors on the prediction of EGFR mutation was evaluated, no statistically significant results were obtained, probably due to the limited number of studies included and the presence of missing data. However, some of those factors might play an essential role and should be considered when developing accurate models to be potentially implemented into clinical practice. Indeed, some of the studies included in our qualitative analysis analyzed the impact of different methodological aspects on the performance of the models. For example, Huang et al.^35^ demonstrated that interobserver variability in tumor segmentation affects the use of radiomics to predict oncogene mutation status, which suggests that automatic or semi-automatic models might be more suitable. In the study by Shiri et al.^88^, the application to radiomics features of ComBat harmonization improved the performance of the models toward more successful prediction of EGFR and KRAS mutations. Likewise, other authors have pointed to the impact of the experimental settings on the robustness of radiomics features^89^, or the influence of CT slice thickness on the predictive performance of radiomics-based models^31^. It is also worth mentioning the relevance of using a particular AI methodology. Although we found no differences in the EGFR mutation predictive performance between ML and DL methods, most likely due to the limited number of available DL-based studies, the latter might offer some advantages over the former. Thus, while in radiomics analysis a process of lesion segmentation and subsequent feature extraction is required, which introduces certain degree of variability and can be a high time-consuming task, DL models only required a bounding box of the lesion, greatly reducing this effect. On the other hand, DL models, and in particular end-to-end convolutional neural network (CNN) models, such those developed in most of the DL studies included in our work^37, 42, 43, 50, 58, 68, 85, 86, 90, 91^, are generally more complex in terms of the number of parameters, allowing to solve more complicated problems than traditional ML models. Considering available evidence, it seems reasonable to think that methodologic approaches should be carefully revised when validation studies are designed and conducted.

Our study has also some limitations, mainly derived from the limitations of the publications included. Thus, it is based on retrospective studies displaying great heterogeneity in terms of methodology and patient clinical characteristics, which clearly hamper the impact of our conclusions. Additionally, the limited available evidence for ALK and KRAS mutations, makes it difficult to draw solid conclusions. Despite this, our work gathers the most up-to-date and complete evidence (all models developed in each of the studies were analyzed) on imaging-based models for the prediction of three of the most important oncogene mutations in NSCLC, following a clinical approach and a special focus on AI models. Our exhaustive review and meta-analyses are intended to provide solid evidence for future research in the field.

In conclusion, radiomics-based models offer a useful and non-invasive method for determining the status of EGFR mutations in NSCLC and seem to retain similar predictive value for ALK and KRAS mutations. Additionally, although the inclusion of clinical variables tends to increase the performance of the models, further validation is required.

## Data Availability

All data produced in the present work are contained in the manuscript.

## FIGURES

**Supplementary Figure S1.**
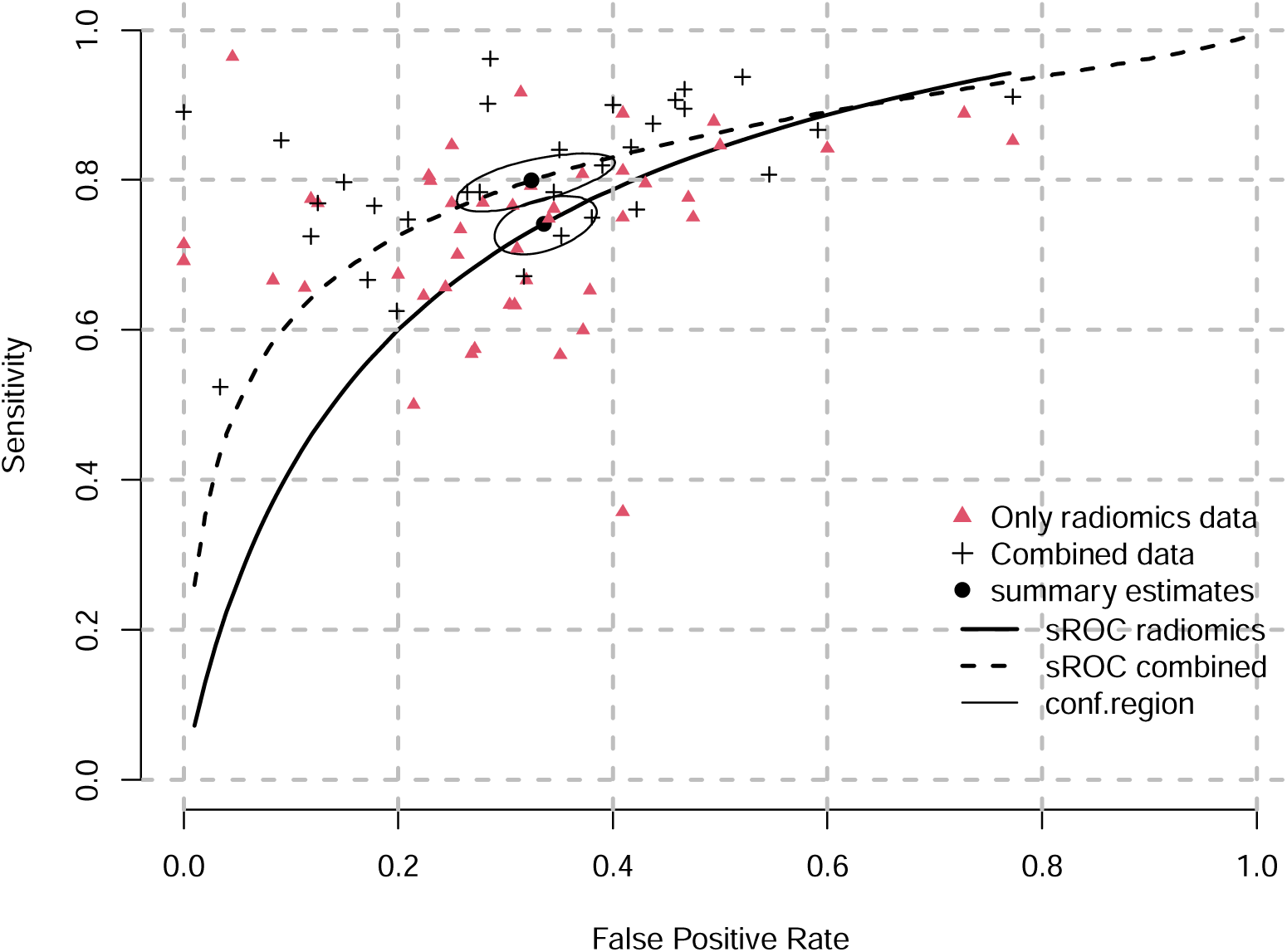
Hierarchical sROC curves of included studies for the comparative performance of radiomics models and combined models (radiomics + clinical data) using machine learning and/or deep learning methods for the prediction of EGFR mutation status (*n* = 24 and *n* = 23 studies, respectively). EGFR, epidermal growth factor receptor.

**Supplementary Figure S2.**
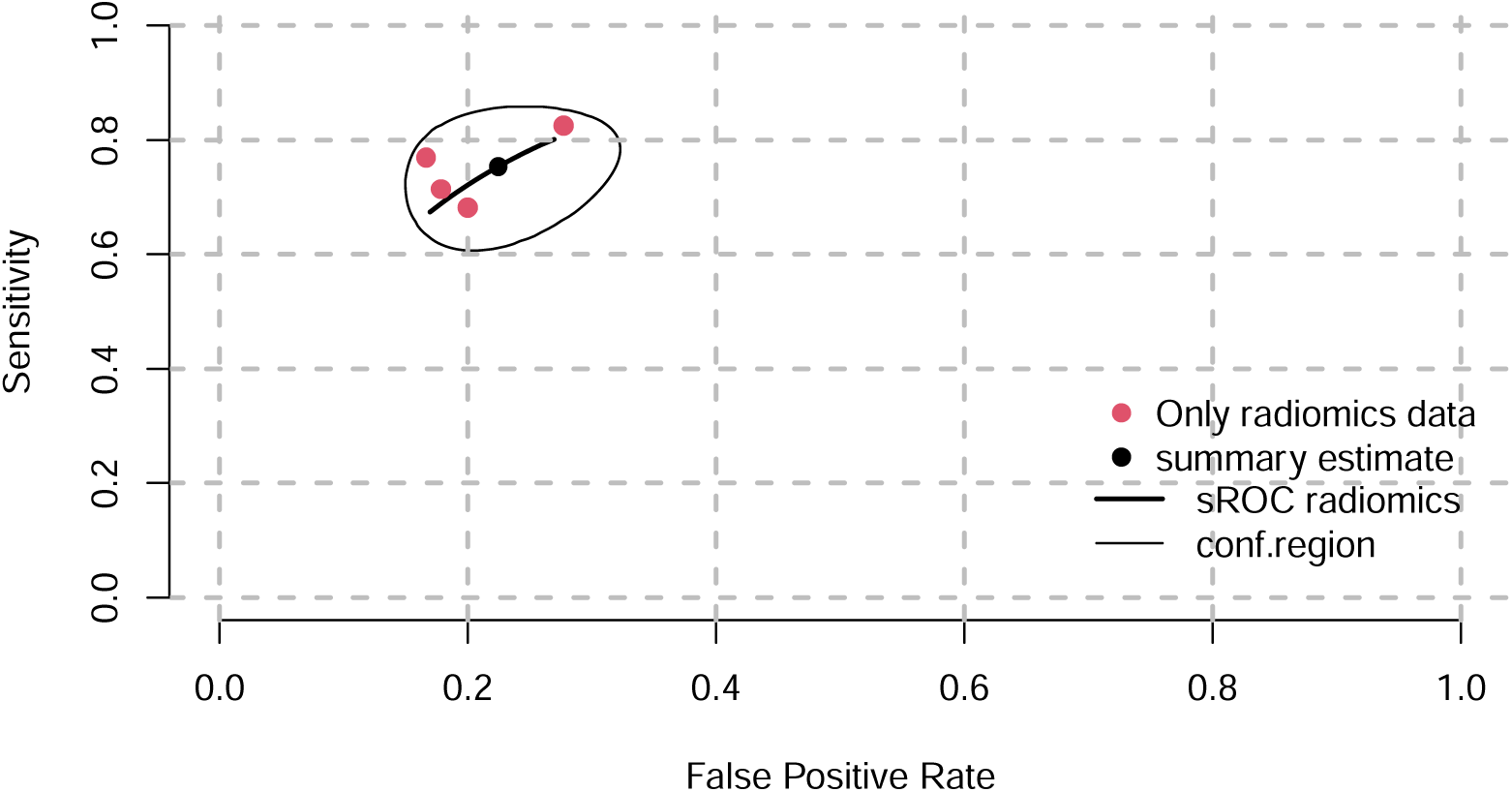
Hierarchical sROC curve of included studies for the performance of radiomics models for the prediction of ALK mutation status (*n* = 3). ALK, anaplastic lymphoma kinase.

**Supplementary Figure S3.**
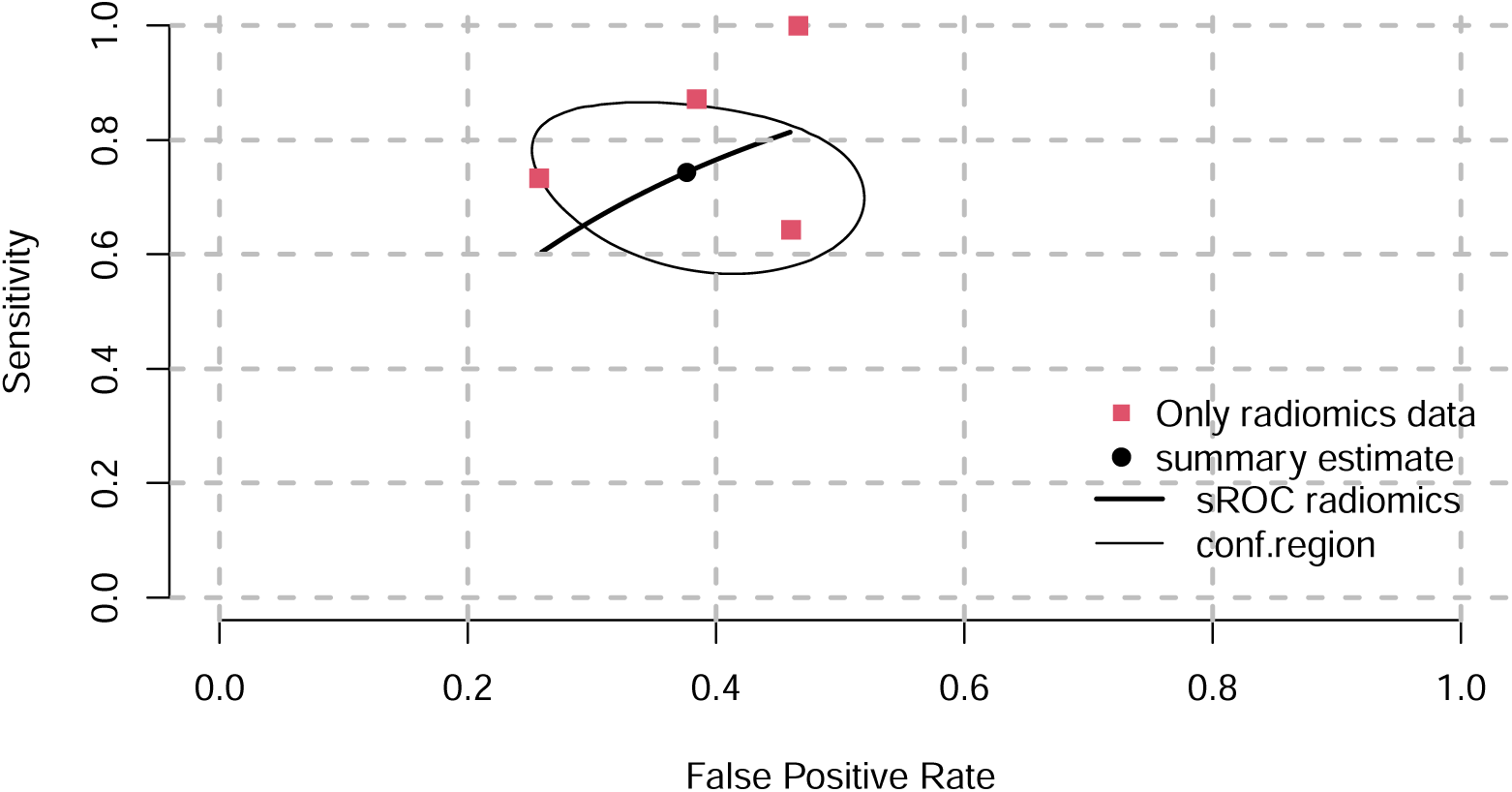
Hierarchical sROC curve of included studies for the performance of radiomics models for the prediction of KRAS mutation status (*n* = 4). KRAS, Kirsten rat sarcoma viral oncogene homologue.

## Supplementary tables

**Supplementary Table S1.** Search strategy applied for the qualitative analysis (systematic review).

| Databases | Search strategy |
| --- | --- |
| MEDLINE (via Pubmed) | ("radiomics"[TIAB] OR "radiomic"[TIAB] OR "texture analysis"[TIAB]) AND ("lung neoplasms"[MESH] OR "lung cancer"[TIAB] OR "NSCLC"[TIAB] OR "non-small cell lung cancer"[TIAB] OR "lung adenocarcinoma"[TIAB]) AND ("mutational status" OR "mutation" OR "molecular subtype" OR "ALK"[TIAB] OR "anaplastic lymphoma kinase"[TIAB] OR "BRAF"[TIAB] OR "EGFR"[TIAB] OR "Epidermal growth factor receptor"[TIAB] OR "ERRB2"[TIAB] OR "Receptor, ErbB-2"[MESH] OR "HER2"[TIAB] OR "KRAS"[TIAB] OR "Kirsten rat sarcoma virus"[TIAB] OR "Proto Oncogene Proteins c met"[TIAB] OR "NTRK"[TIAB] OR "ROS"[TIAB] OR "c-ros"[TIAB]) |
| COCHRANE LIBRARY | ("radiomics" OR "radiomic" OR "texture analysis") AND ("lung neoplasms" OR "lung cancer" OR "NSCLC" OR "non-small cell lung cancer") AND ("mutational status" OR "mutation" OR "molecular subtype" OR "ALK" OR "anaplastic lymphoma kinase" OR "BRAF" OR "EGFR" OR "ERRB2" OR "Receptor, ErbB-2" OR "HER2" OR "KRAS" OR "Kirsten rat sarcoma virus" OR "Proto Oncogene Proteins c met" OR "NTRK" OR "ROS" OR "c-ros") |
| EMBASE | ('radiomics':ab,ti OR 'radiomics'/exp OR 'radiomic':ab,ti OR 'texture analysis':ab,ti) AND ('lung cancer'/exp OR 'lung cancer':ab,ti OR 'NSCLC'/exp OR 'NSCLC':ab,ti OR 'non small cell lung cancer'/exp OR 'non small cell lung cancer':ab,ti OR 'lung adenocarcinoma'/exp OR 'lung adenocarcinoma':ab,ti) AND |
|  | ('mutational status':ab,ti OR ('mutational' NEAR/2 'status') OR<br>'mutation':ab,ti OR 'mutation'/exp OR 'molecular subtype':ab,ti OR<br>('molecular' NEAR/2 'subtype') OR 'ALK':ab,ti OR 'ALK gene'/exp<br>OR 'anaplastic lymphoma kinase':ab,ti OR 'anaplastic lymphoma<br>kinase'/exp OR 'BRAF':ab,ti OR 'BRAF gene'/exp OR 'EGFR':ab,ti<br>OR 'EGFR gene'/exp OR 'Epidermal growth factor receptor':ab,ti<br>OR 'Epidermal growth factor receptor gene'/exp OR 'ERRB2':ab,ti<br>OR 'ERRB2 gene'/exp OR 'epidermal growth factor receptor 2'/exp<br>OR 'epidermal growth factor receptor 2':ab,ti OR 'HER2':ab,ti OR<br>'KRAS':ab,ti OR 'KRAS gene'/exp OR 'Kirsten rat sarcoma<br>virus':ab,ti OR 'Kirsten rat sarcoma virus'/exp OR 'Proto Oncogene<br>Proteins c met' OR 'MET':ab,ti OR 'MET gene'/exp OR<br>'NTRK':ab,ti OR 'NTRK gene'/exp OR 'c ros oncogene 1':ab,ti OR<br>'ROS1':ab,ti OR 'ROS1 gene'/exp) |

**Supplementary Table S2.** Quality assessment results obtained after CLAIM evaluation.

| *N* | Study | Score |  |  | Mean score | Cut-off |
| --- | --- | --- | --- | --- | --- | --- |
|  |  | Reviewer 1<br>(A.J.P.) | Reviewer 2<br>(F.B.B.) | Reviewer 3<br>(A.P.P.) |  |  |
| 1 | Chang et al. 2021<sup>1</sup> | 28 | 24 | 27 | 26 | 17 |
| 2 | Chang et al. 2021<sup>2</sup> | 26 | 25 | 26 | 26 | 17 |
| 3 | Dong et al. 2021<sup>3</sup> | 23 | 22 | 19 | 21 | 18 |
| 4 | Dong et al. 2022<sup>4</sup> | 25 | 22 | 21 | 23 | 18 |
| 5 | Feng et al. 2022<sup>5</sup> | 20 | 22 | 23 | 22 | 18.5 |
| 6 | Gao et al. 2023<sup>6</sup> | 22 | 22 | 20 | 21 | 17.5 |
| 7 | Huo et al. 2022<sup>7</sup> | 22 | 25 | 25 | 24 | 17.5 |
| 8 | Jia et al. 2019<sup>8</sup> | 19 | 19 | 20 | 19 | 17 |
| 9 | Jiang et al. 2022<sup>9</sup> | 24 | 23 | 22 | 23 | 16.5 |
| 10 | Le et al. 2021<sup>10</sup> | 22 | 20 | 21 | 21 | 17.5 |
| 11 | Li et al. 2018<sup>11</sup> | 25 | 28 | 22 | 25 | 19 |
| 12 | Li et al. 2019<sup>12</sup> | 23 | 22 | 21 | 22 | 17 |
| 13 | Li et al. 2020<sup>13</sup> | 25 | 23 | 22 | 23 | 19 |
| 14 | Li et al. 2022<sup>14</sup> | 21 | 21 | 21 | 21 | 17 |
| 15 | Liu et al. 2020<sup>15</sup> | 22 | 25 | 23 | 23 | 17 |
| 16 | Liu et al. 2022<sup>16</sup> | 24 | 23 | 22 | 23 | 17 |
| 17 | Lu et al. 2020<sup>17</sup> | 27 | 30 | 26 | 28 | 17.5 |
| 18 | Lu et al. 2022<sup>18</sup> | 22 | 25 | 21 | 23 | 17.5 |
| 19 | Ma et al. 2020<sup>19</sup> | 24 | 26 | 22 | 24 | 17 |
| 20 | Nair et al. 2021<sup>20</sup> | 21 | 22 | 19 | 21 | 17.5 |
| 21 | Ninomiya et al. 2021<sup>21</sup> | 21 | 22 | 21 | 21 | 17.5 |
| 22 | Ninomiya et al. 2023<sup>22</sup> | 21 | 22 | 22 | 22 | 17 |
| 23 | Rios Velazquez et al. 2017<sup>23</sup> | 19 | 23 | 20 | 21 | 18 |
| 24 | Rossi et al. 2021<sup>24</sup> | 20 | 22 | 19 | 20 | 17.5 |
| 25 | Song et al. 2020<sup>25</sup> | 27 | 28 | 27 | 27 | 19 |
| 26 | Tu et al. 2019<sup>26</sup> | 19 | 21 | 20 | 20 | 17 |
| 27 | Wang et al. 2022<sup>27</sup> | 23 | 26 | 26 | 25 | 18 |
| 28 | Wang et al. 2022<sup>28</sup> | 24 | 22 | 22 | 23 | 19 |
| 29 | Weng et al. 2021<sup>29</sup> | 24 | 25 | 24 | 24 | 17.5 |
| 30 | Wu 2020<sup>30</sup> | 20 | 22 | 21 | 21 | 17 |
| 31 | Yang 2020<sup>31</sup> | 22 | 23 | 23 | 23 | 17 |
| 32 | Yang 2022<sup>32</sup> | 19 | 18 | 18 | 18 | 17 |
| 33 | Zhang 2018<sup>33</sup> | 26 | 27 | 23 | 25 | 17 |
| 34 | Zhang 2020<sup>34</sup> | 19 | 19 | 19 | 19 | 17 |
| 35 | Zhang 2020<sup>35</sup> | 22 | 23 | 23 | 23 | 17 |
| 36 | Zhang 2021<sup>36</sup> | 27 | 26 | 26 | 26 | 17 |
| 37 | Zhao 2022<sup>37</sup> | 23 | 24 | 23 | 23 | 19 |
| 38 | Zhu 2022<sup>38</sup> | 22 | 22 | 20 | 21 | 17.5 |

**Supplementary Table S3.** Studies included in the different meta-analyses conducted.

| *N* | **EGFR** |  | **ALK** | **KRAS** |
| --- | --- | --- | --- | --- |
|  | **Radiomics models** | **Combined models** | **Radiomics models** | **Radiomics models** |
| 1 | Chang et al. 2021<sup>1</sup> | Dong et al. 2022<sup>4</sup> | Chang et al. 2021<sup>2</sup> | Dong et al. 2021<sup>3</sup> |
| 2 | Dong et al. 2021<sup>3</sup> | Gao et al. 2023<sup>6</sup> | Ma et al. 2020<sup>19</sup> | Le et al. 2021<sup>10</sup> |
| 3 | Feng et al. 2022<sup>5</sup> | Huo et al. 2022<sup>7</sup> | Song et al. 2020<sup>25</sup> | RiosVelazquez et al. 2017<sup>23</sup> |
| 4 | Gao et al. 2023<sup>6</sup> | Jia et al. 2019<sup>8</sup> |  | Wang et al. 2022<sup>28</sup> |
| 5 | Le et al. 2021<sup>10</sup> | Jiang et al. 2022<sup>9</sup> |  |  |
| 6 | Li et al. 2018<sup>11</sup> | Li et al. 2018<sup>11</sup> |  |  |
| 7 | Li et al. 2019<sup>12</sup> | Li et al. 2019<sup>12</sup> |  |  |
| 8 | Li et al. 2022<sup>14</sup> | Li et al. 2020<sup>13</sup> |  |  |
| 9 | Liu et al. 2020<sup>15</sup> | Liu et al. 2020<sup>15</sup> |  |  |
| 10 | Liu et al. 2022<sup>16</sup> | Lu et al. 2020<sup>17</sup> |  |  |
| 11 | Lu et al. 2020<sup>17</sup> | Lu et al. 2022<sup>18</sup> |  |  |
| 12 | Lu et al. 2022<sup>18</sup> | Ninomiya et al. 2023<sup>22</sup> |  |  |
| 13 | Nair et al. 2021<sup>20</sup> | Rios Velazquez et al. 2017<sup>23</sup> |  |  |
| 14 | Ninomiya et al. 2021<sup>21</sup> | Rossi et al. 2021<sup>24</sup> |  |  |
| 15 | RiosVelazquez et al. 2017<sup>23</sup> | Tu et al. 2019<sup>26</sup> |  |  |
| 16 | Tu et al. 2019<sup>26</sup> | Wang et al. 2022<sup>27</sup> |  |  |
| 17 | Wang et al. 2022<sup>27</sup> | Weng et al. 2021<sup>29</sup> |  |  |
| 18 | Weng et al. 2021<sup>29</sup> | Wu 2020<sup>30</sup> |  |  |
| 19 | Yang 2020<sup>31</sup> | Yang 2022<sup>32</sup> |  |  |
| 20 | Zhang 2020<sup>34</sup> | Zhang 2018<sup>33</sup> |  |  |
| 21 | Zhang 2020<sup>35</sup> | Zhang 2020<sup>35</sup> |  |  |
| 22 | Zhang 2021<sup>36</sup> | Zhang 2021<sup>36</sup> |  |  |
| 23 | Zhao 2022<sup>37</sup> | Zhu 2022<sup>38</sup> |  |  |
| 24 | Zhu 2022<sup>38</sup> |  |  |  |

ALK, anaplastic lymphoma kinase; EGFR, epidermal growth factor receptor; KRAS, Kirsten rat sarcoma viral oncogene homologue.

**Supplementary Table S4.**
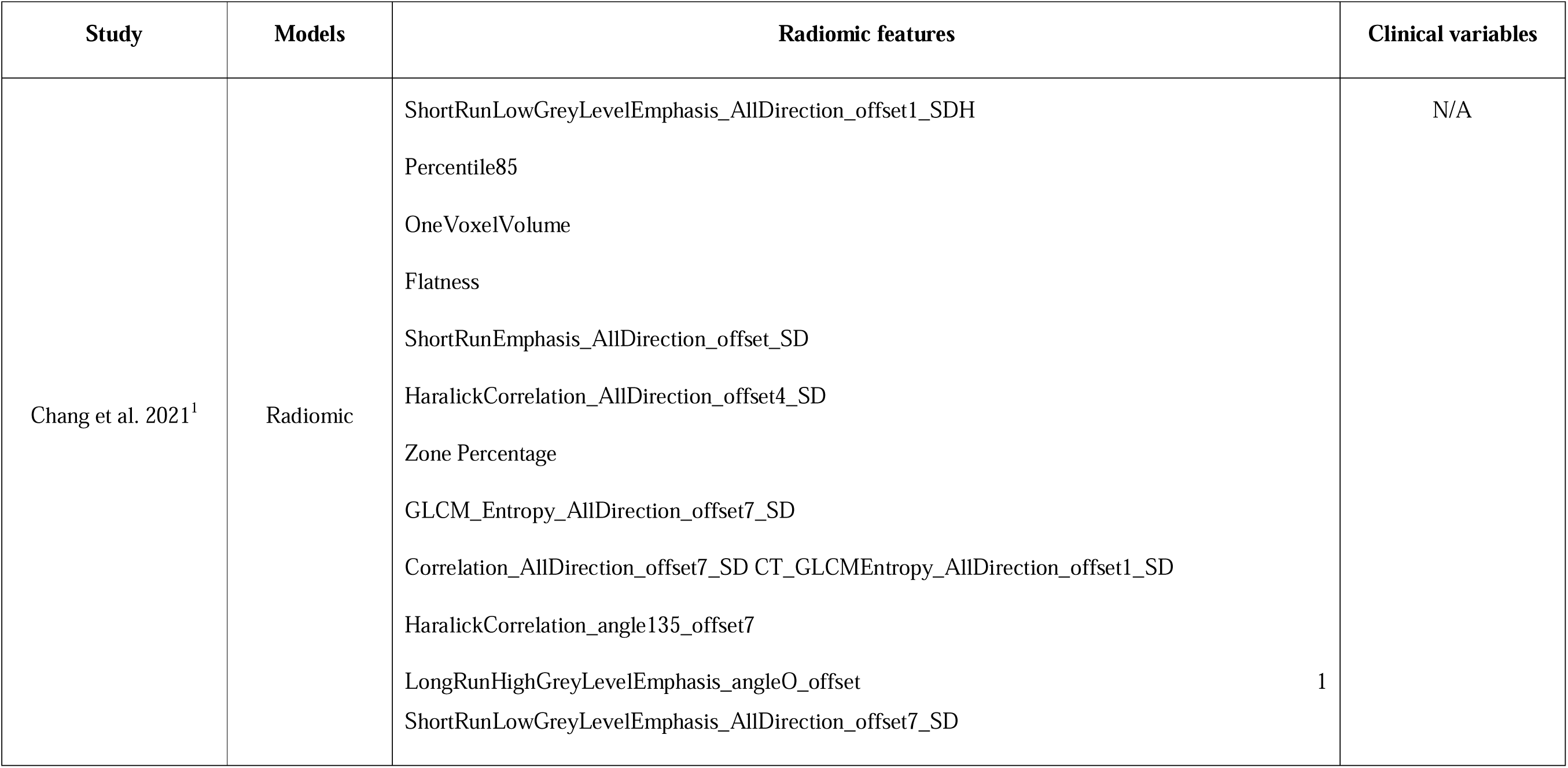
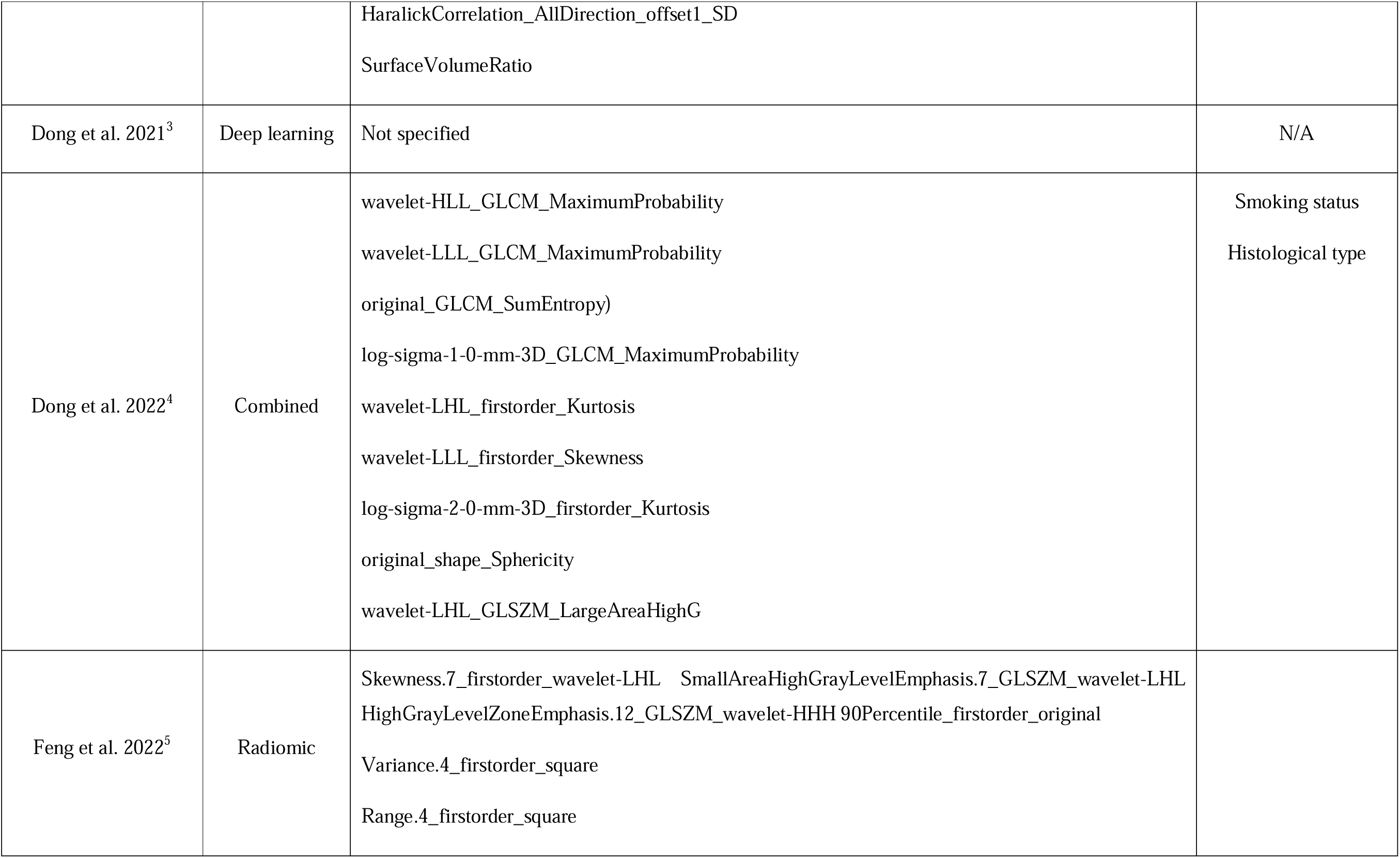
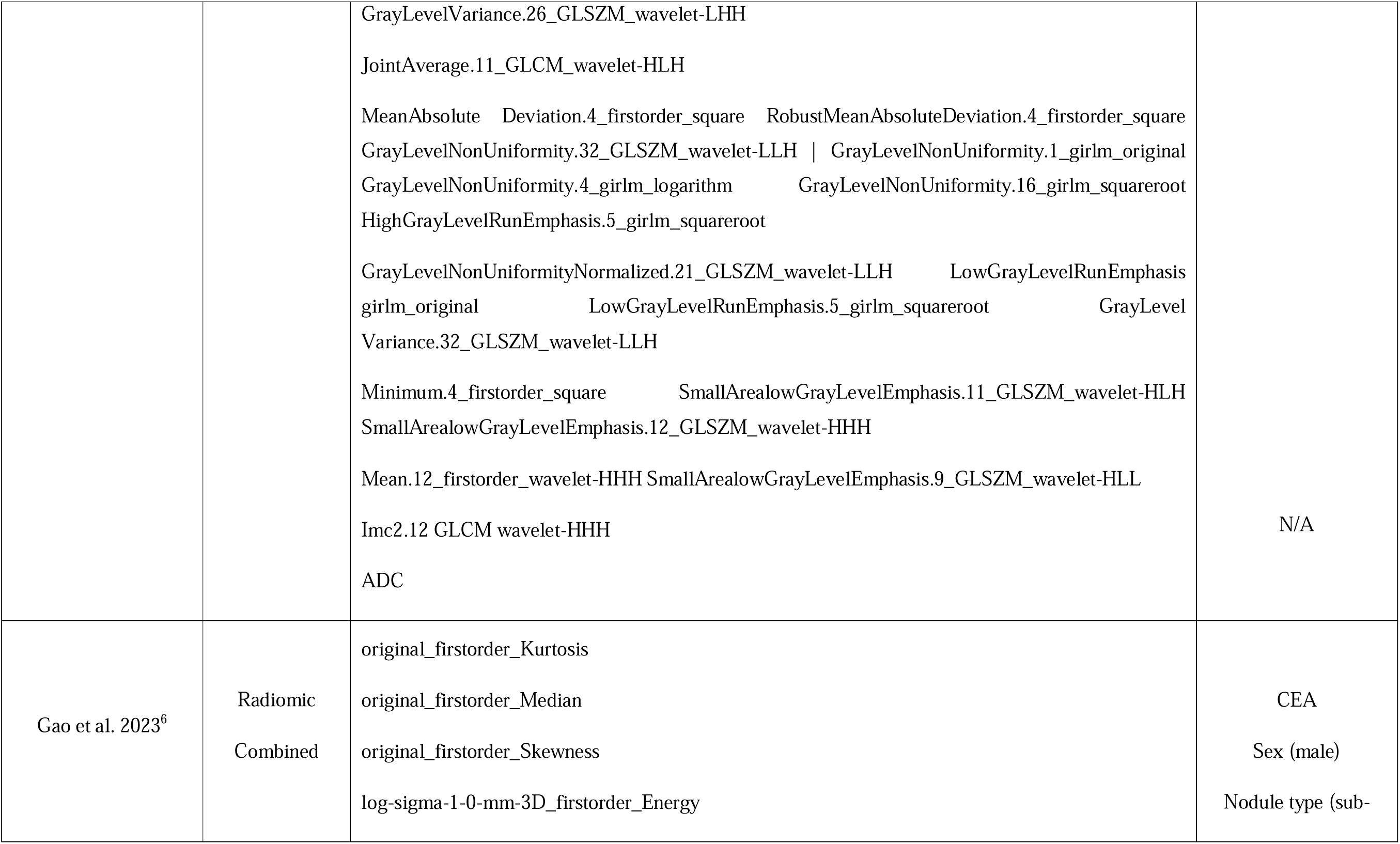
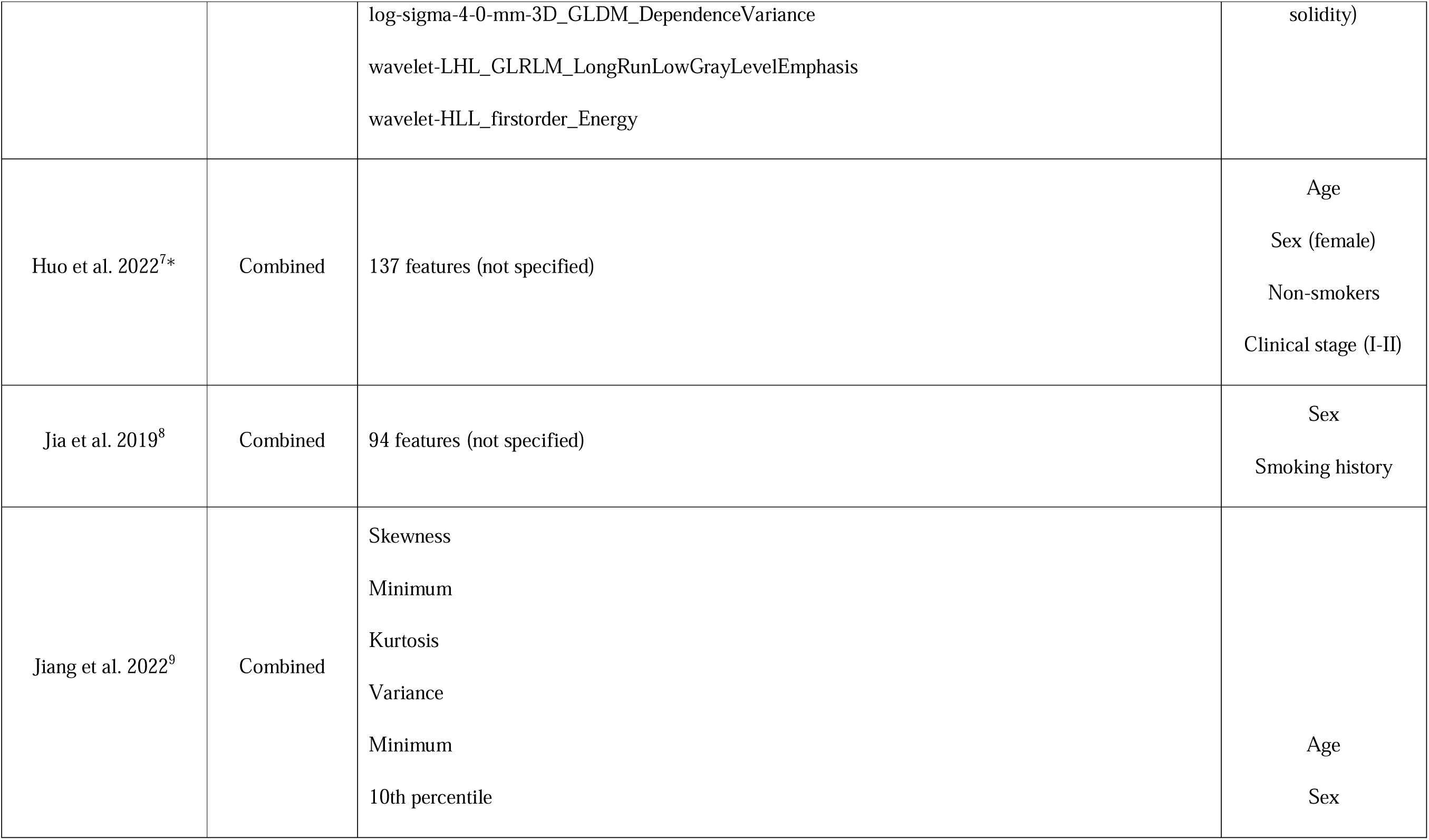
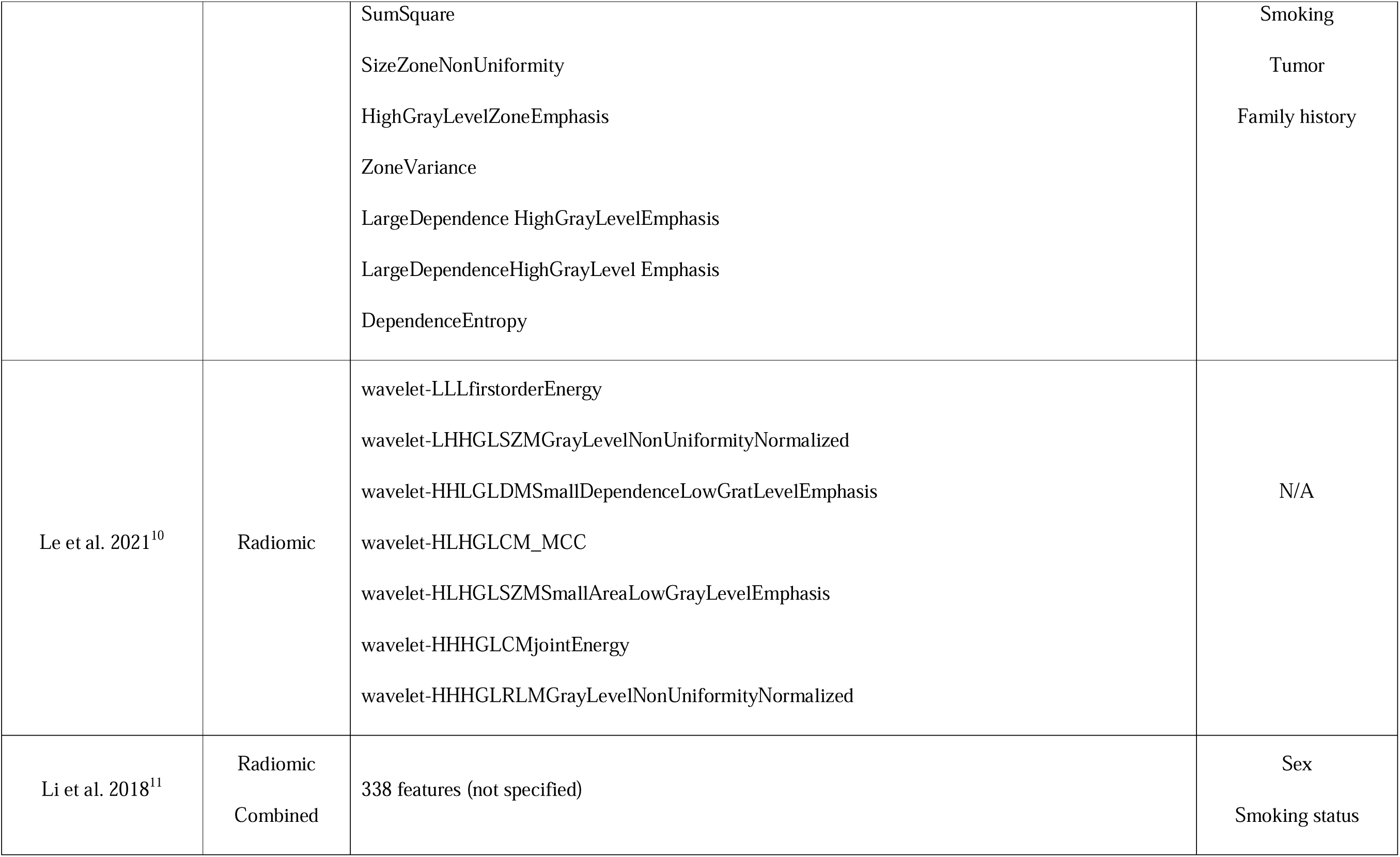
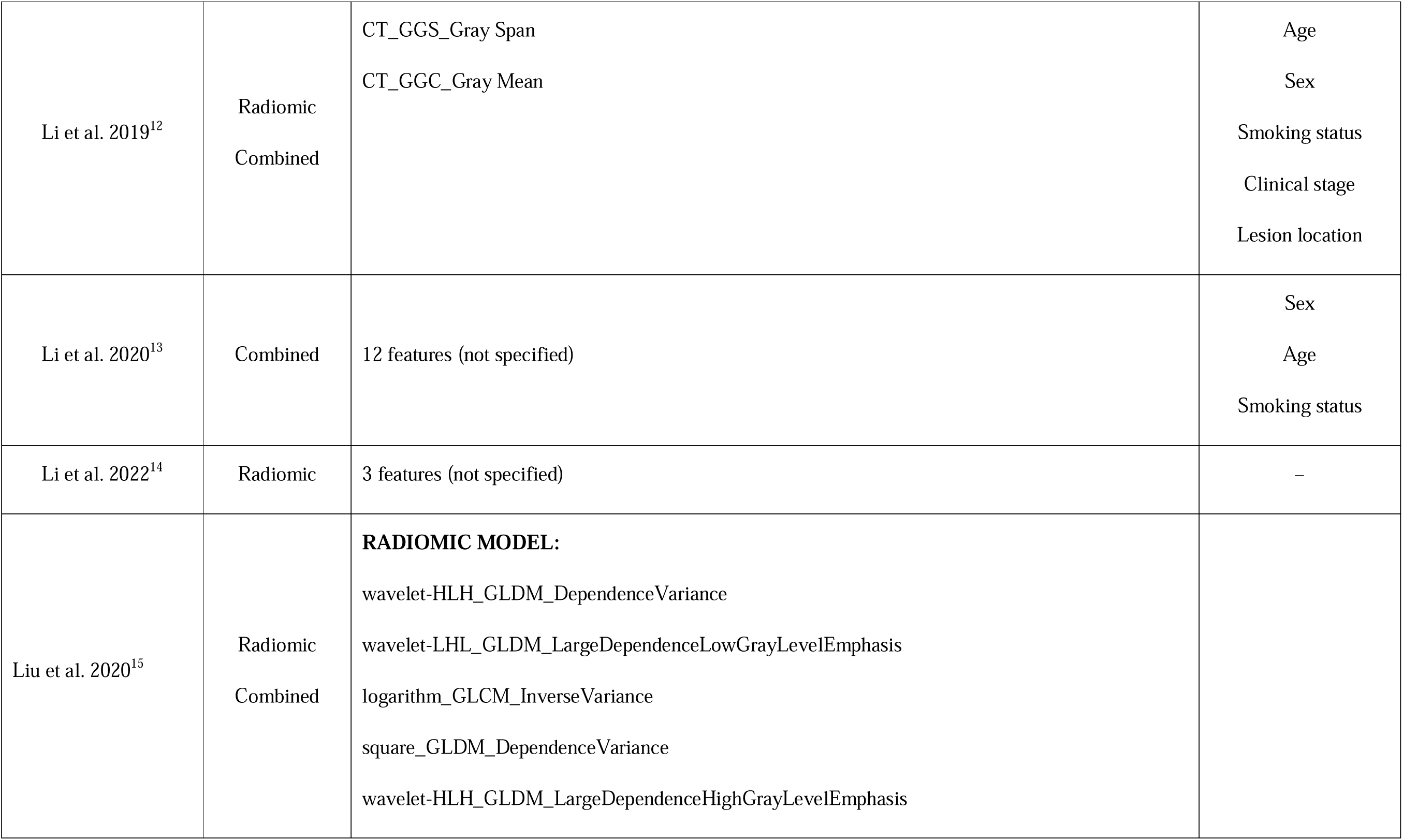
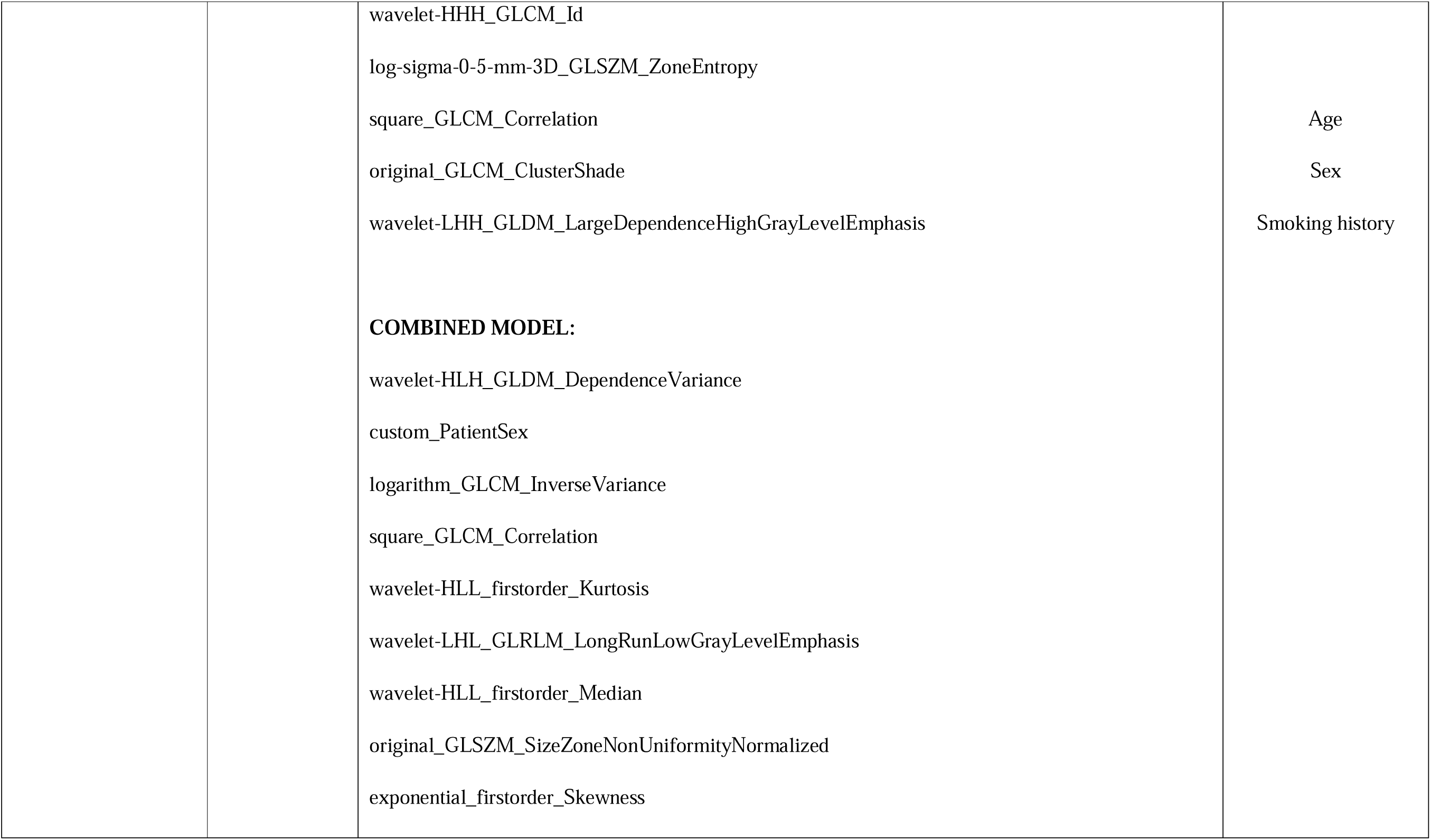
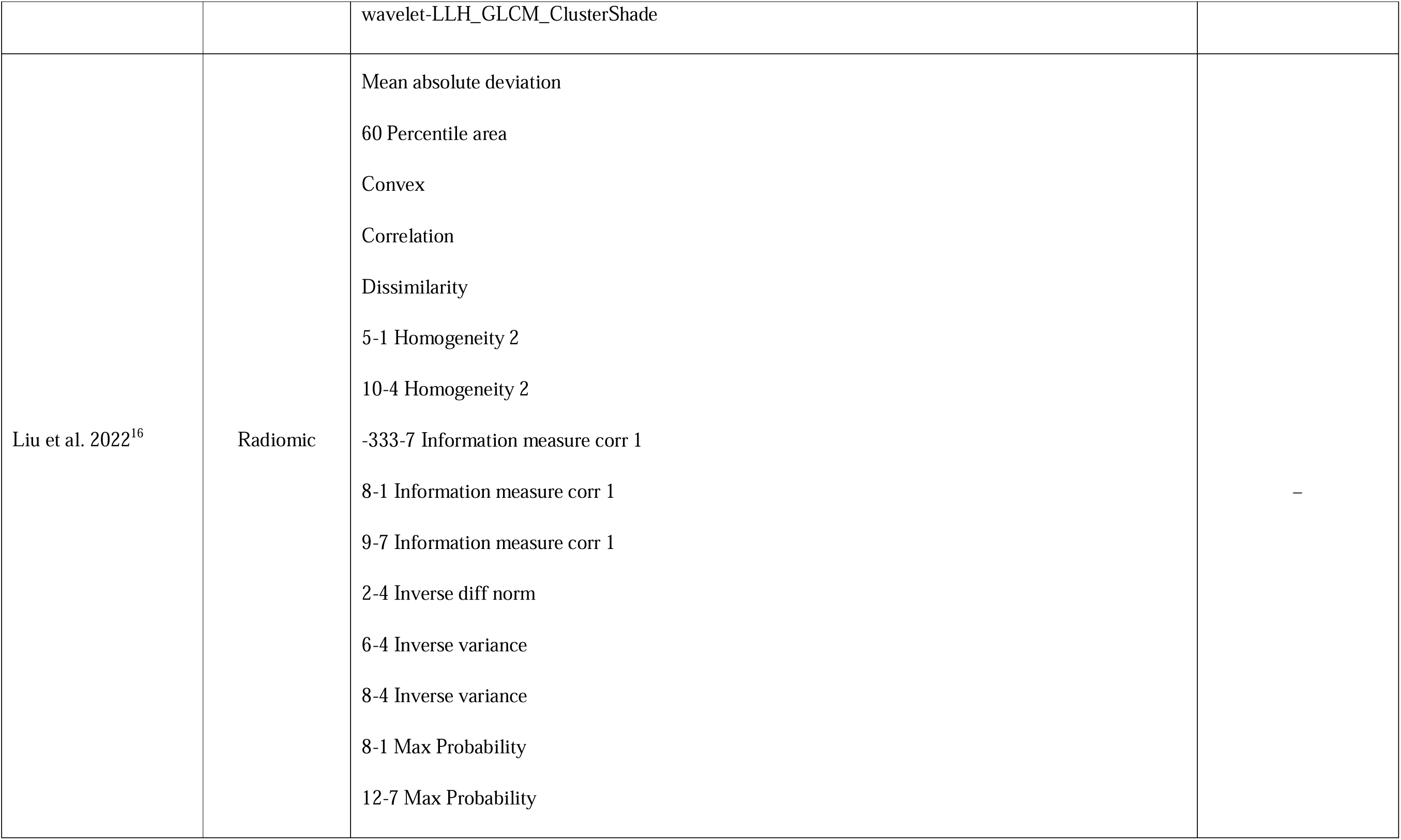
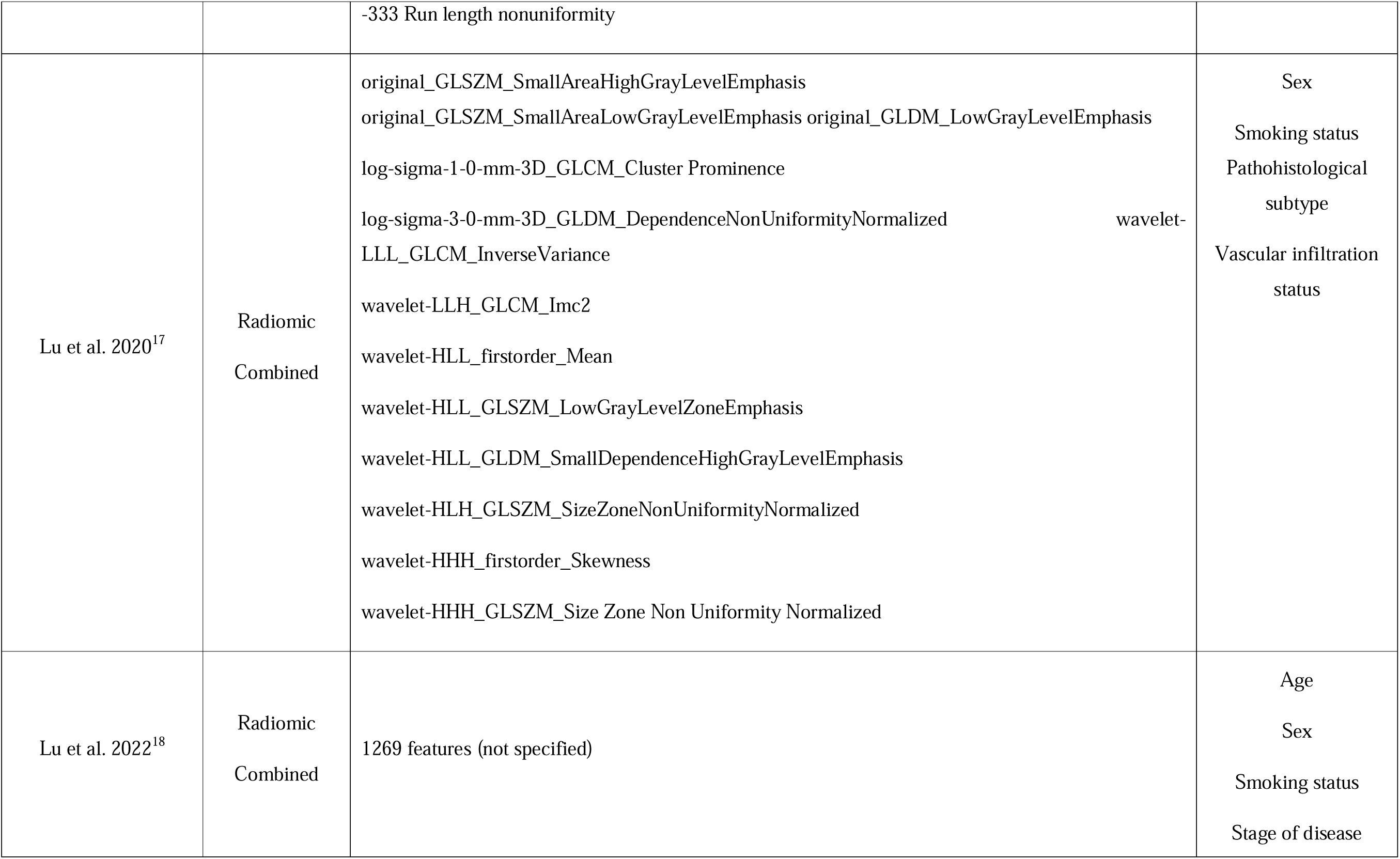
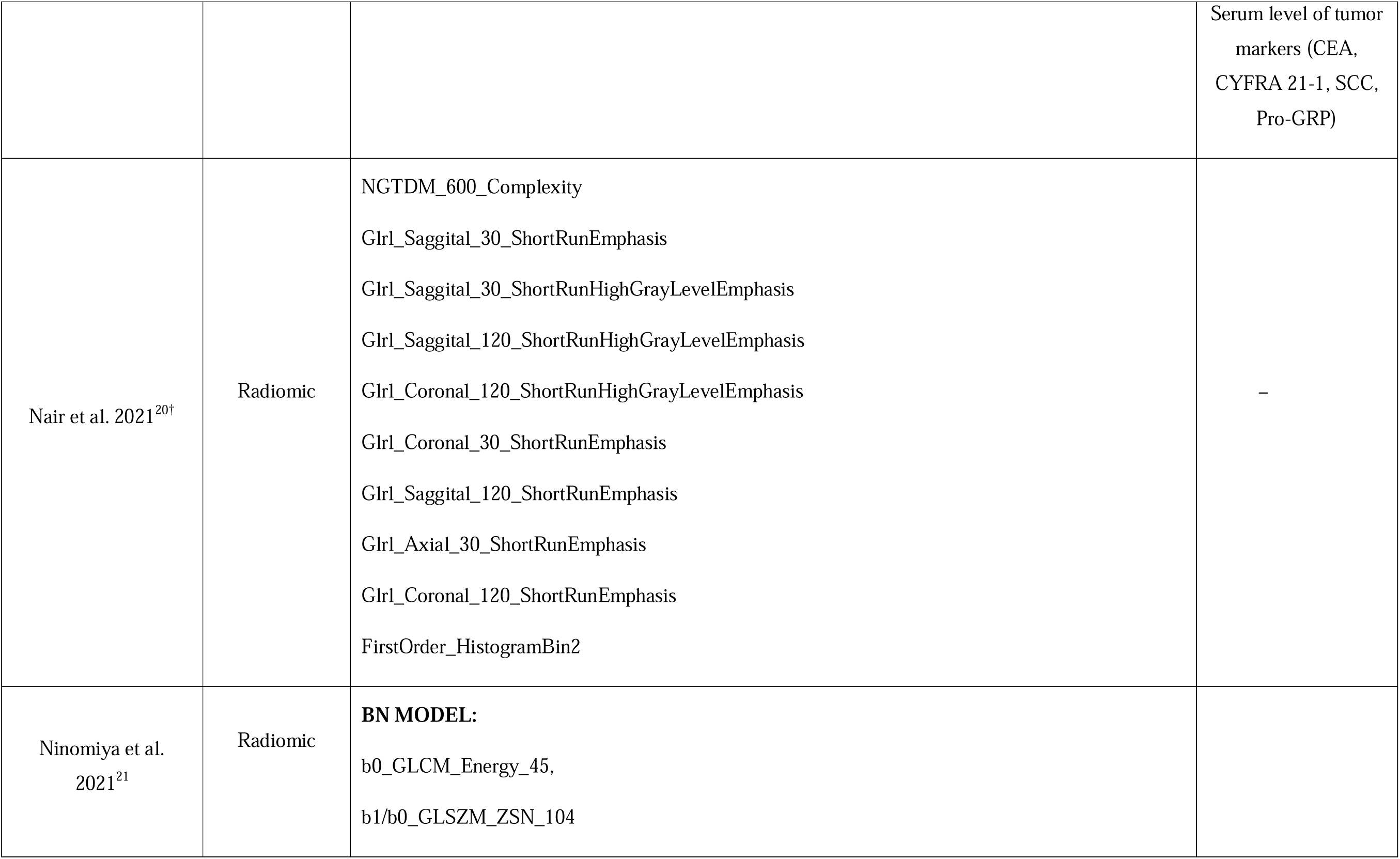
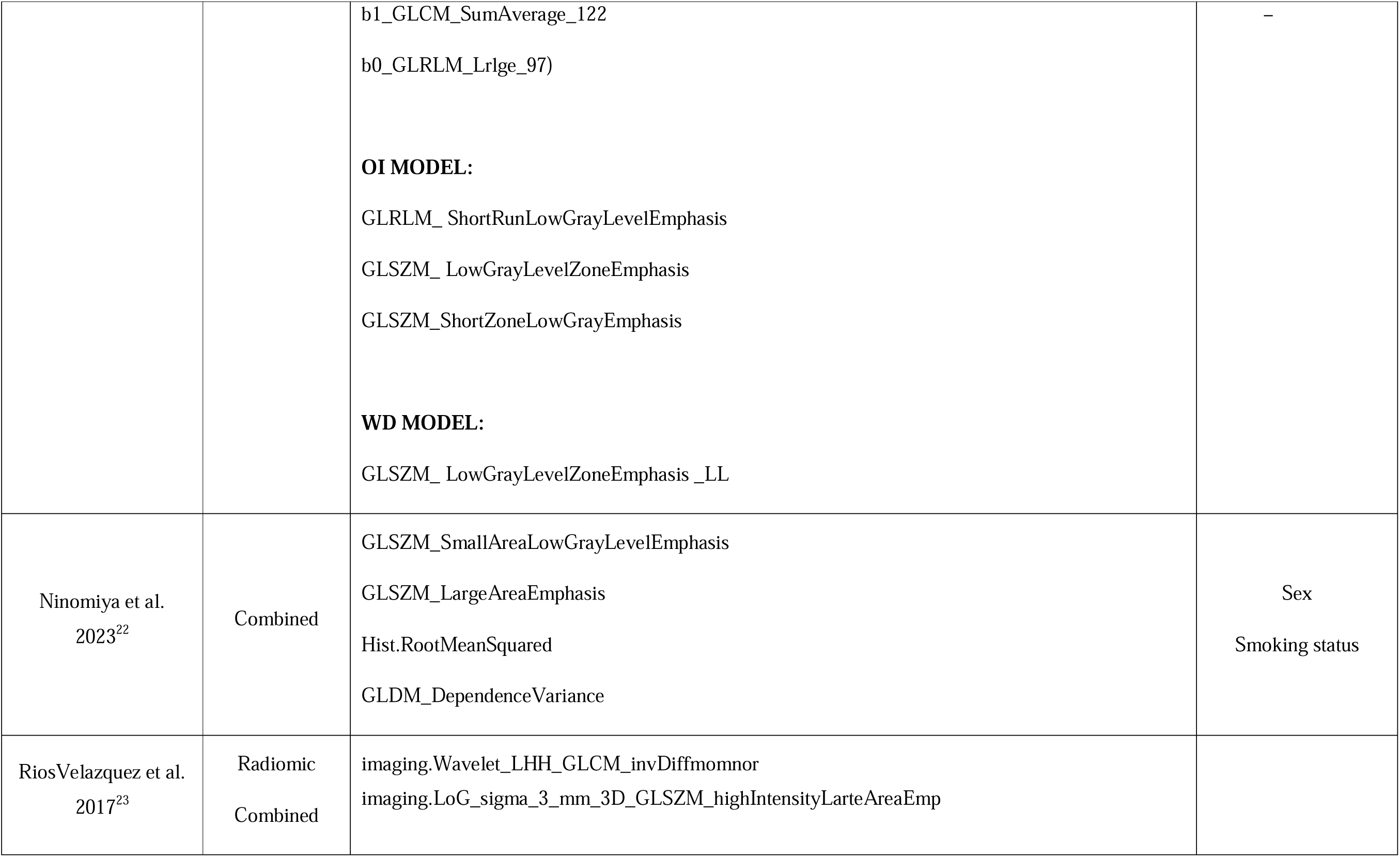
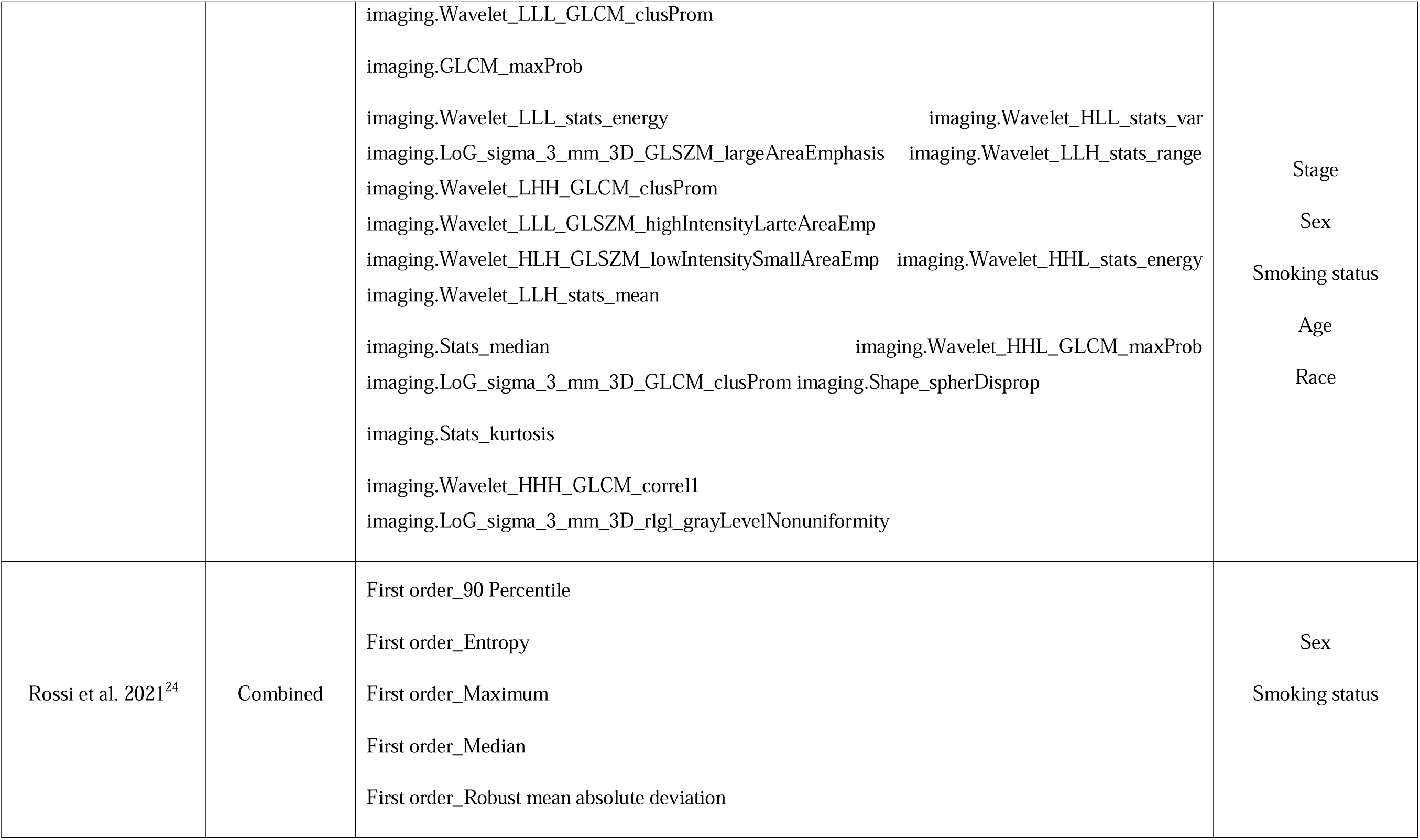
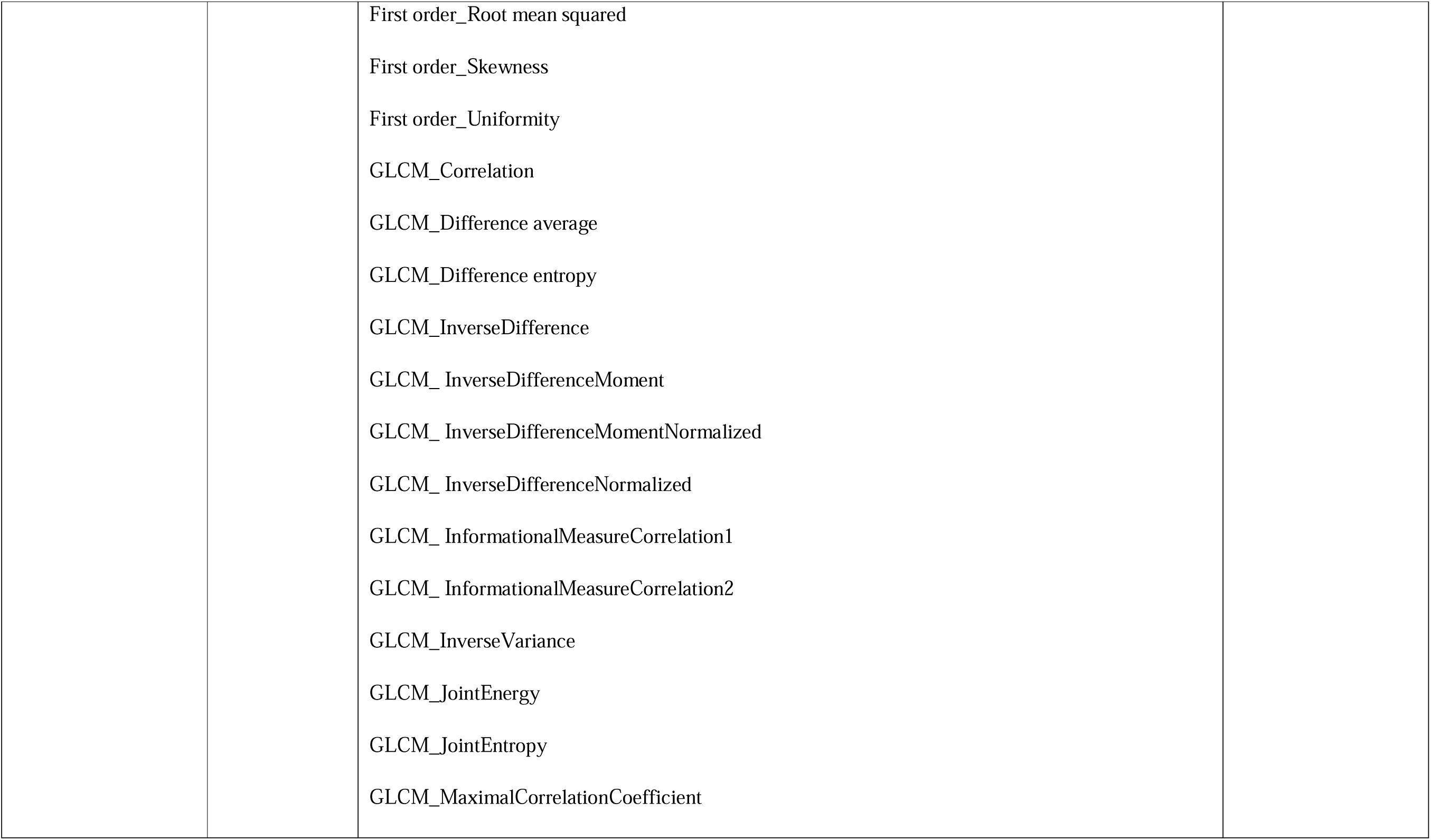
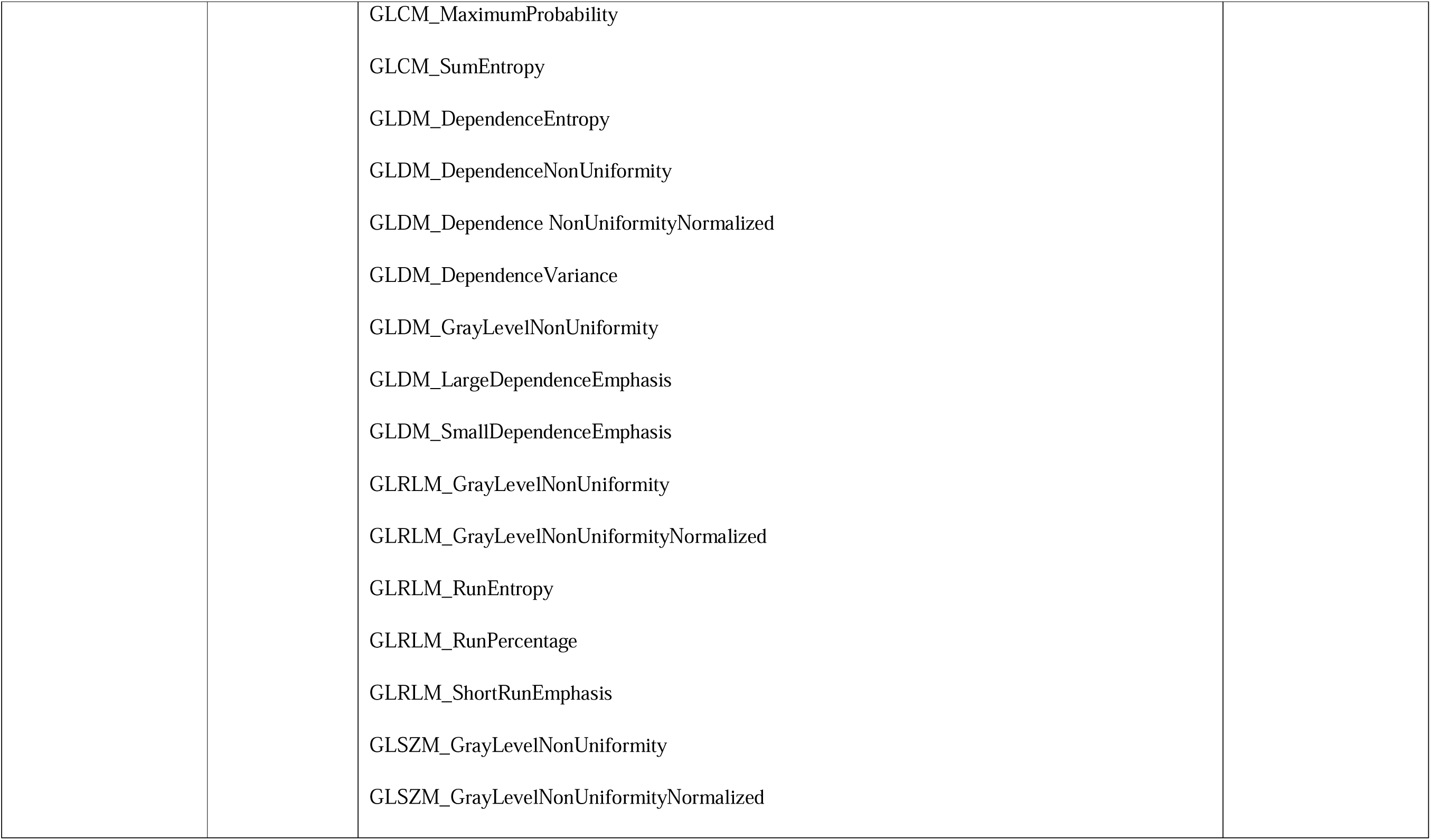
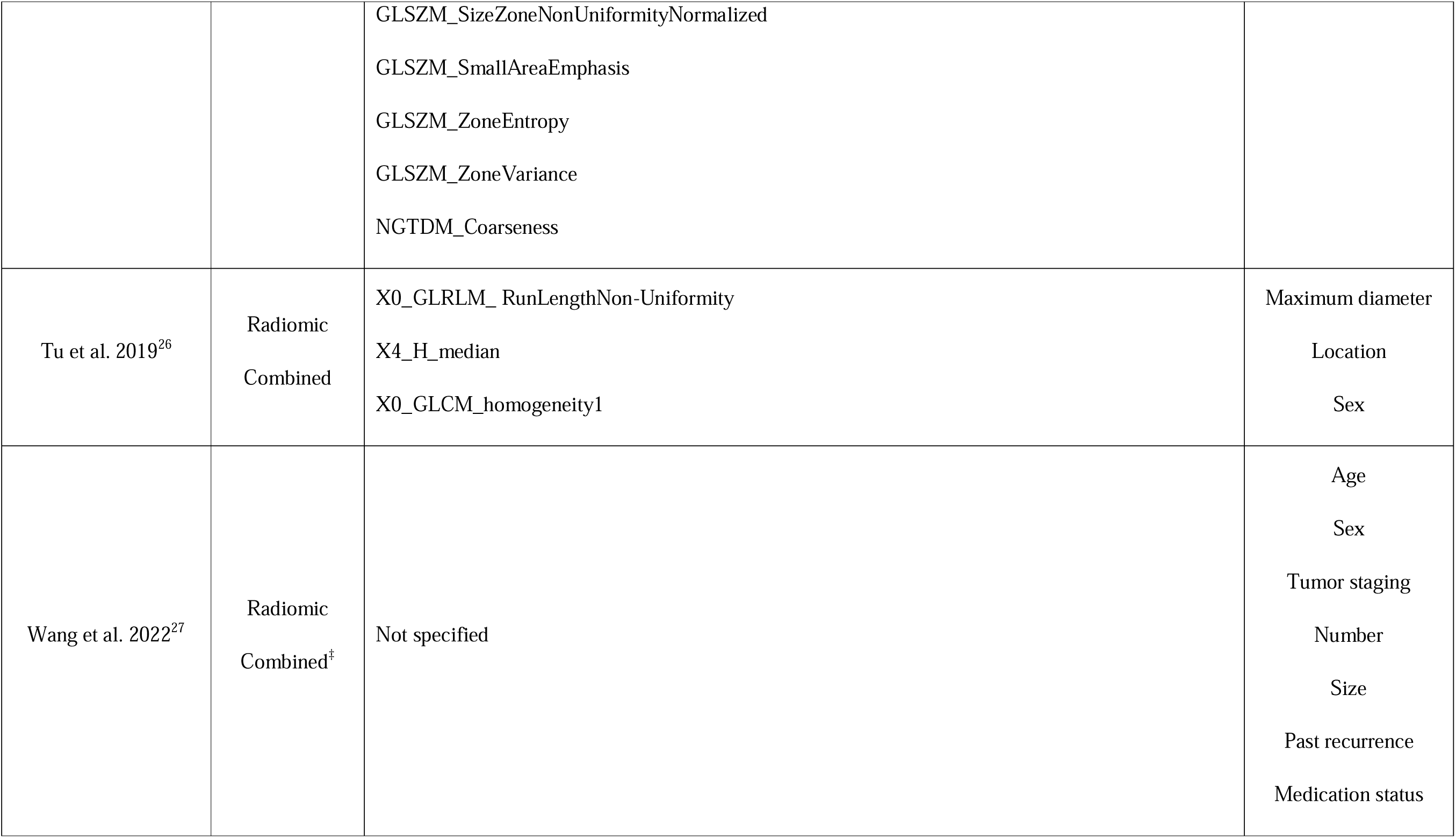
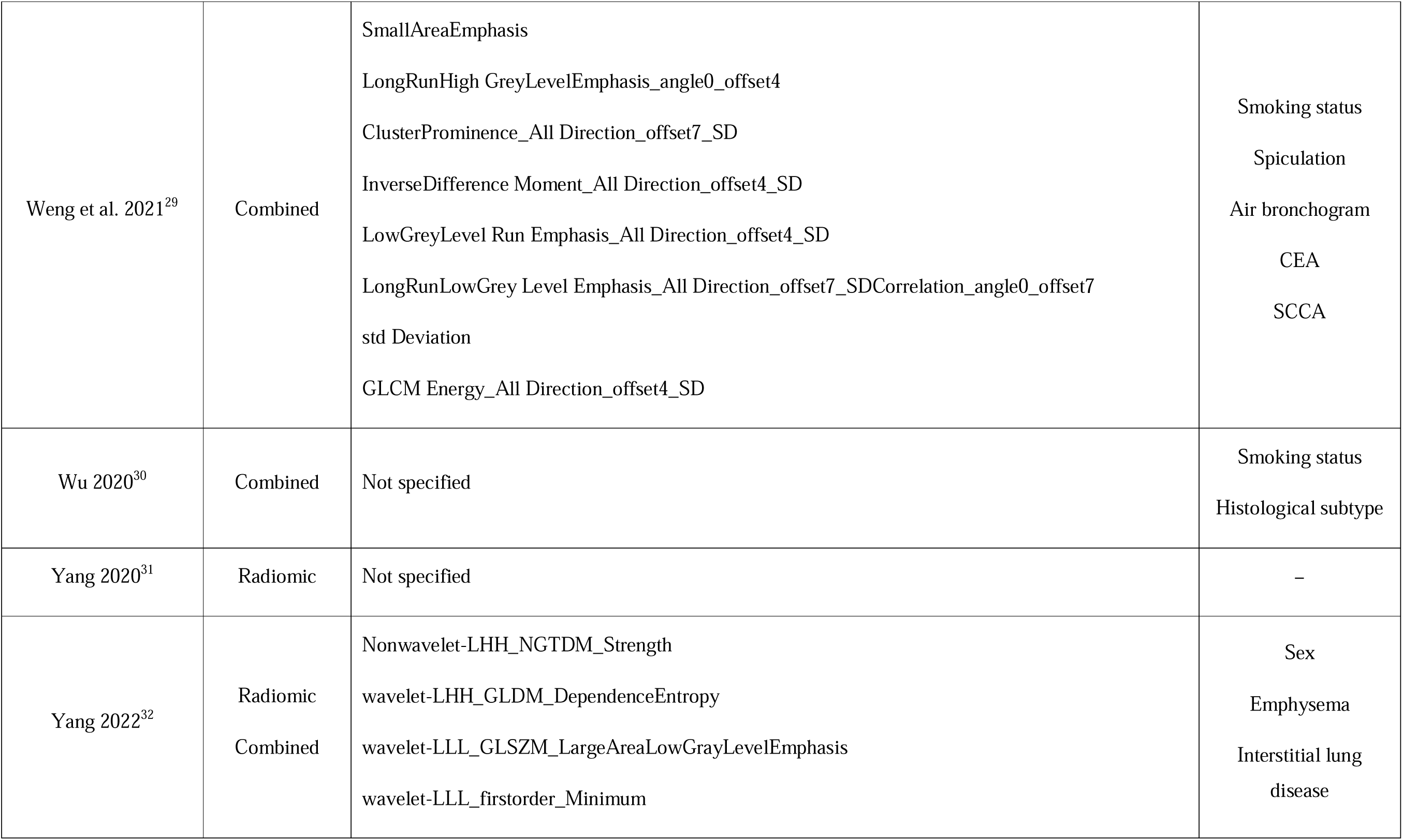
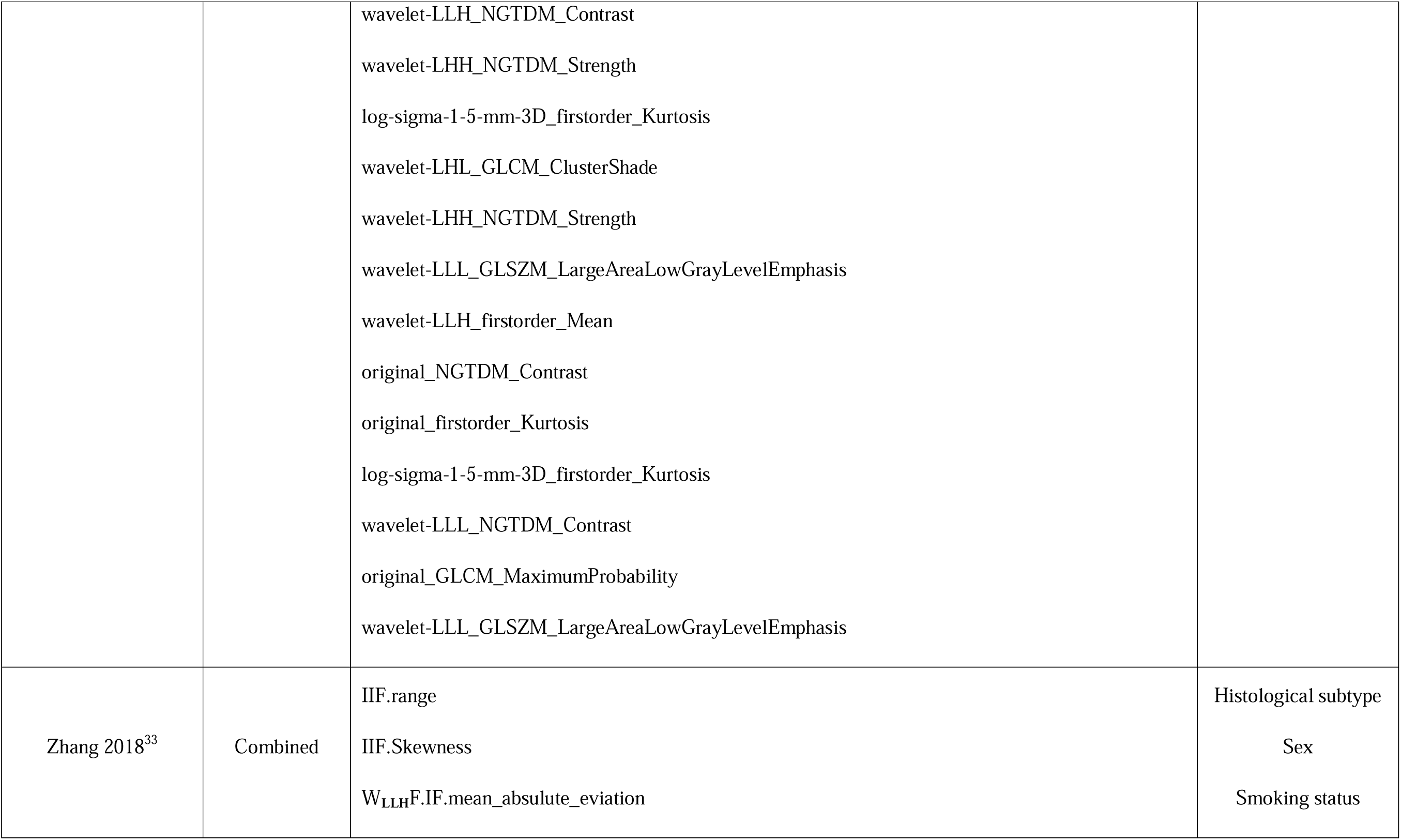
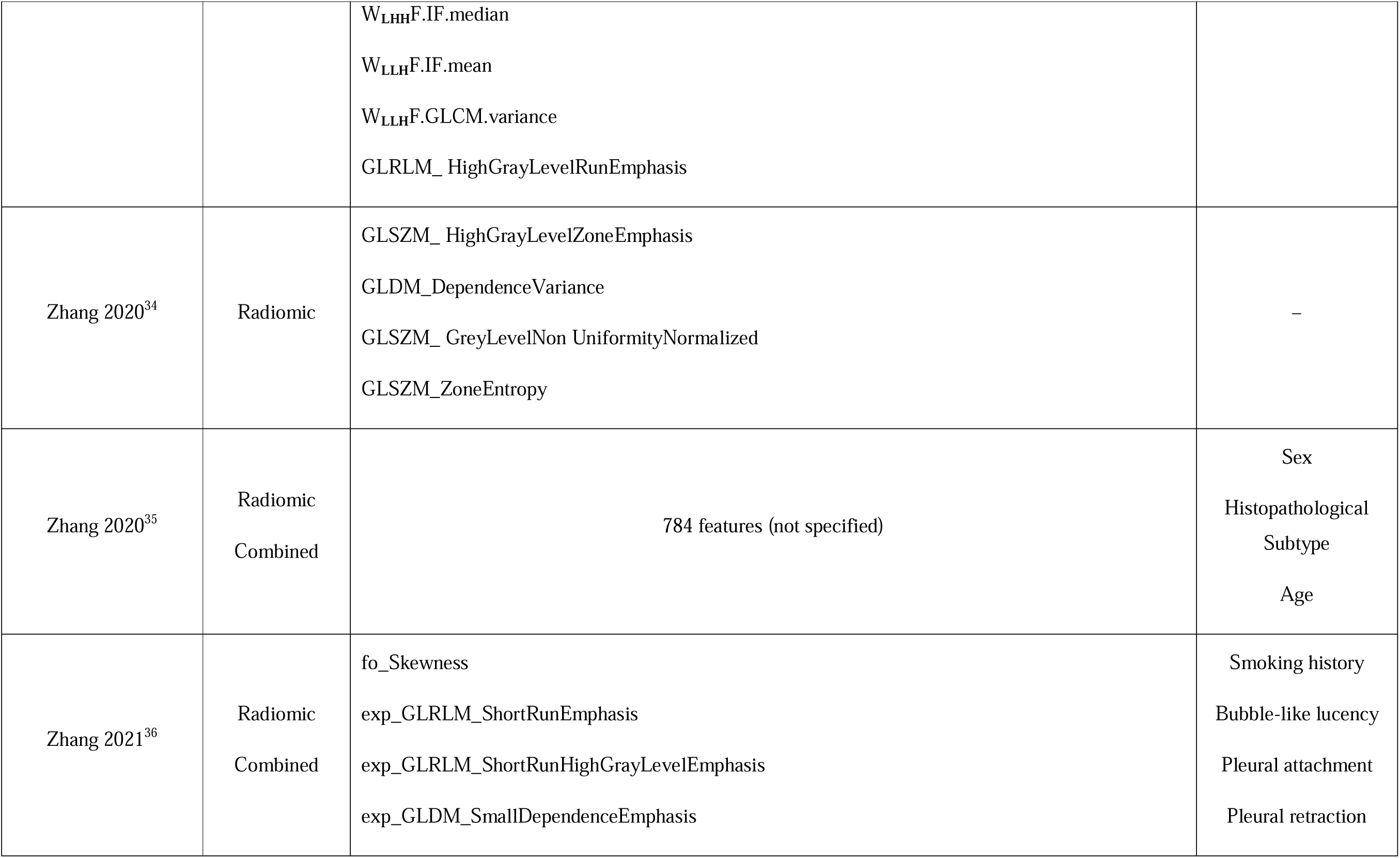
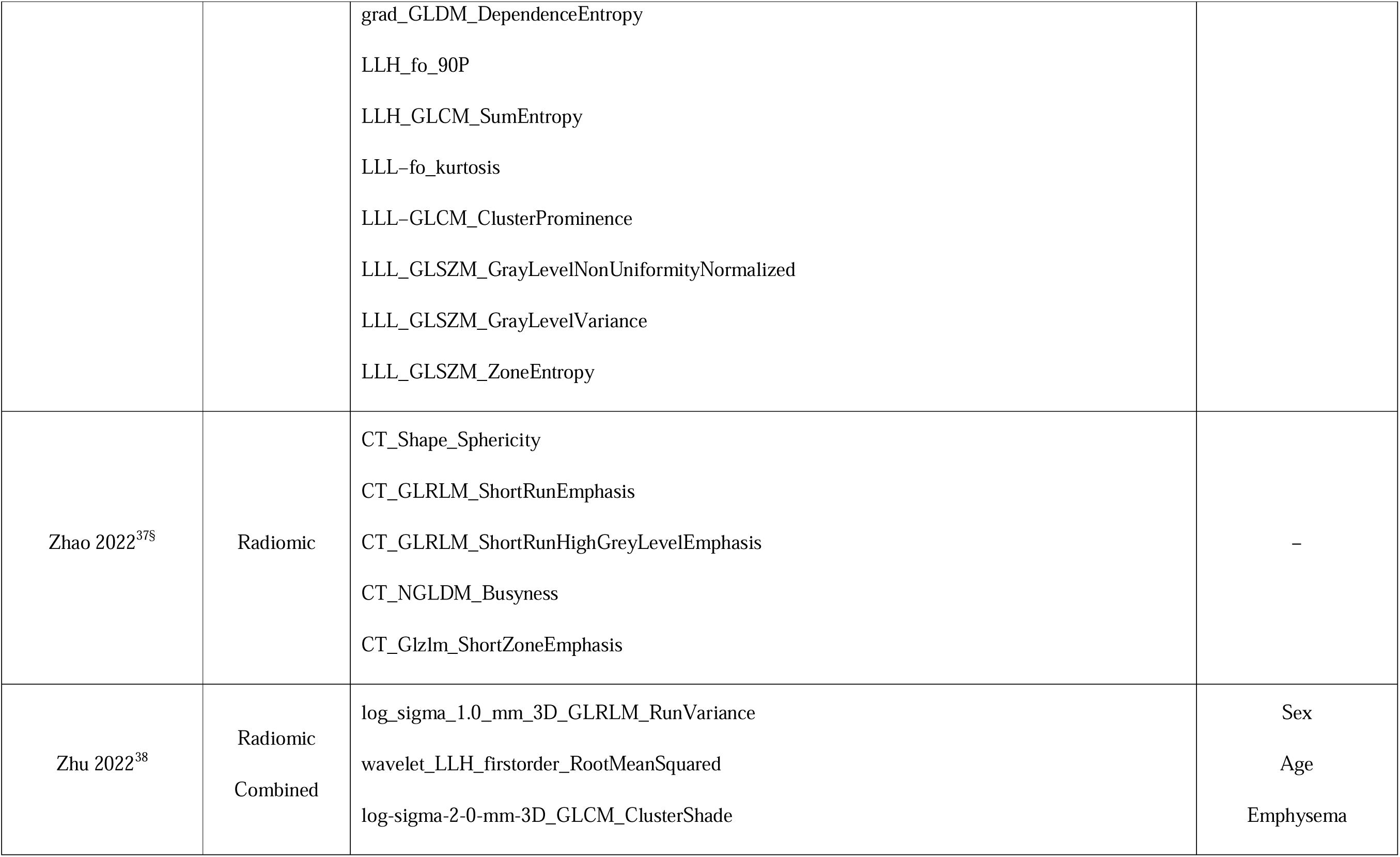
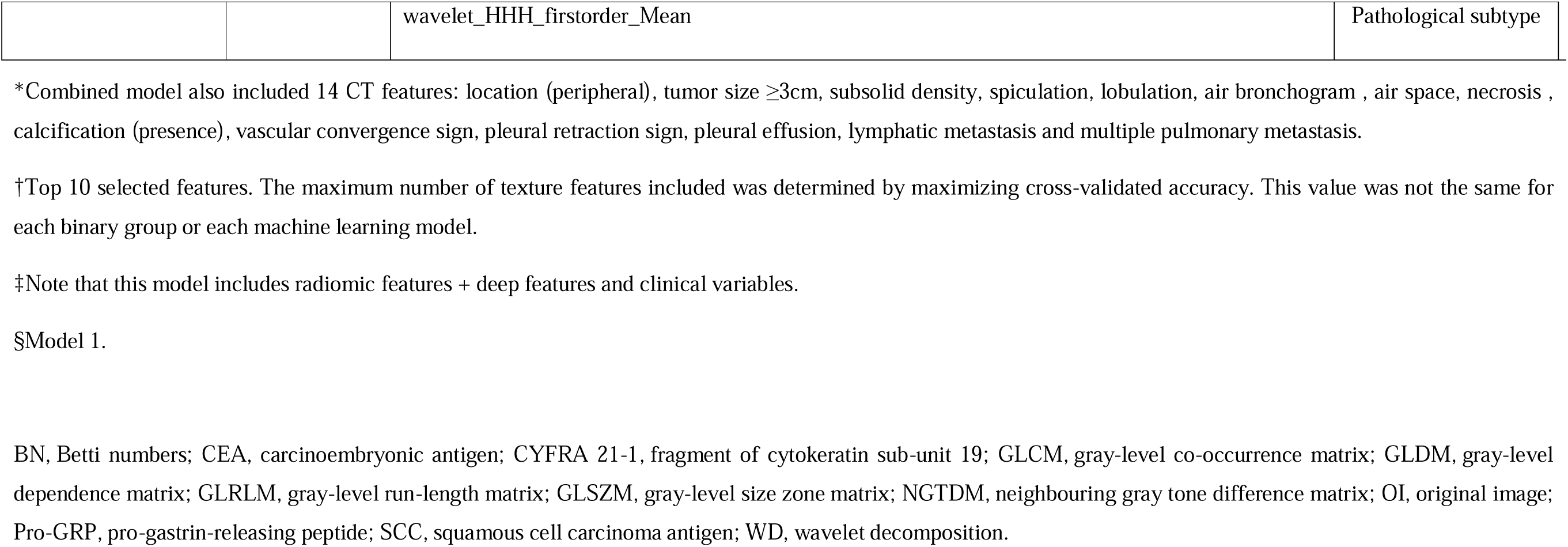
Type of models (radiomic model/deep learning or combined [radiomic features + clinical variables]) developed in the studies for EGFR prediction and the radiomics/clinical features included. EGFR, epidermal growth factor receptor.

**Supplementary Table S5.**
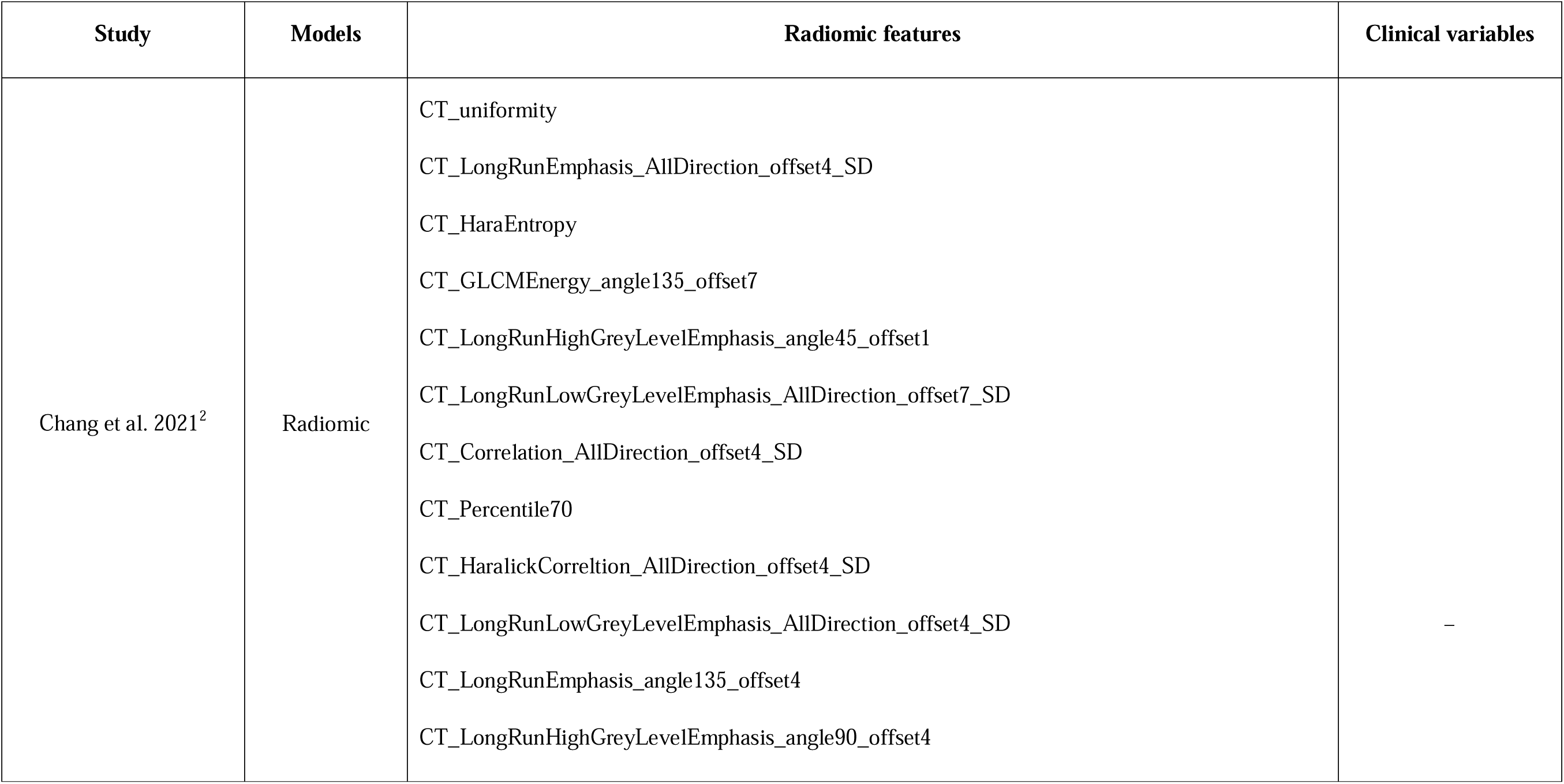
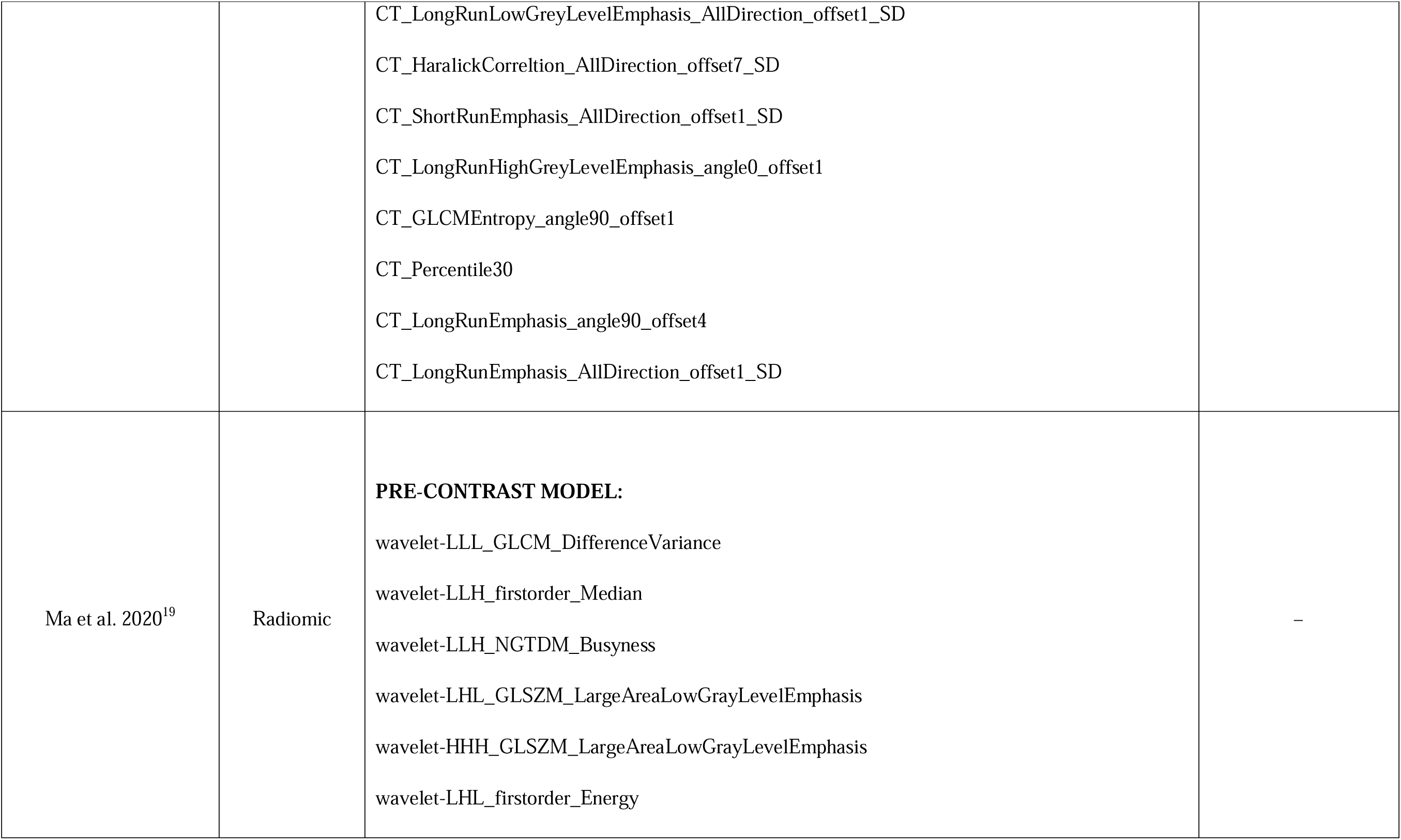
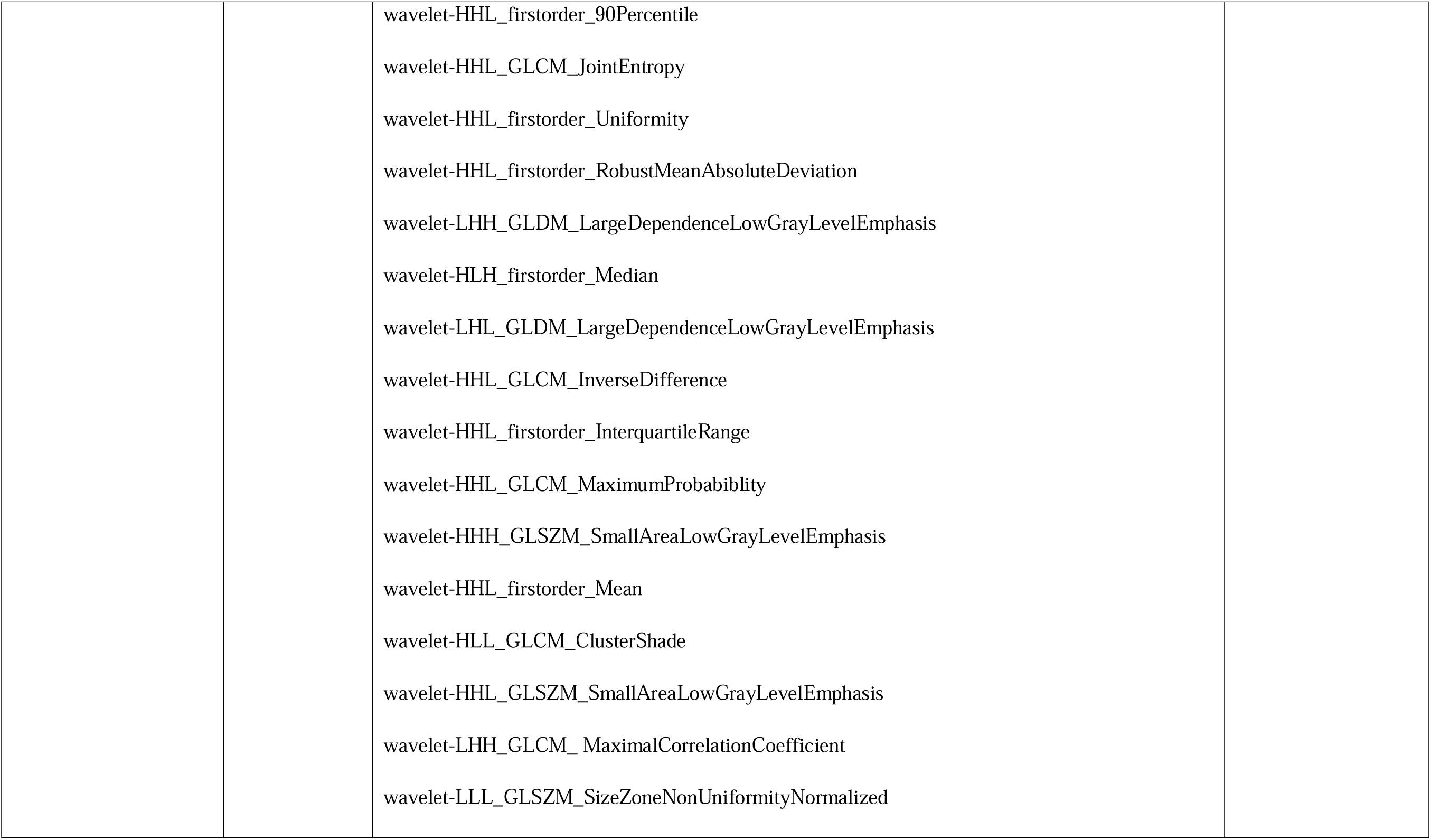
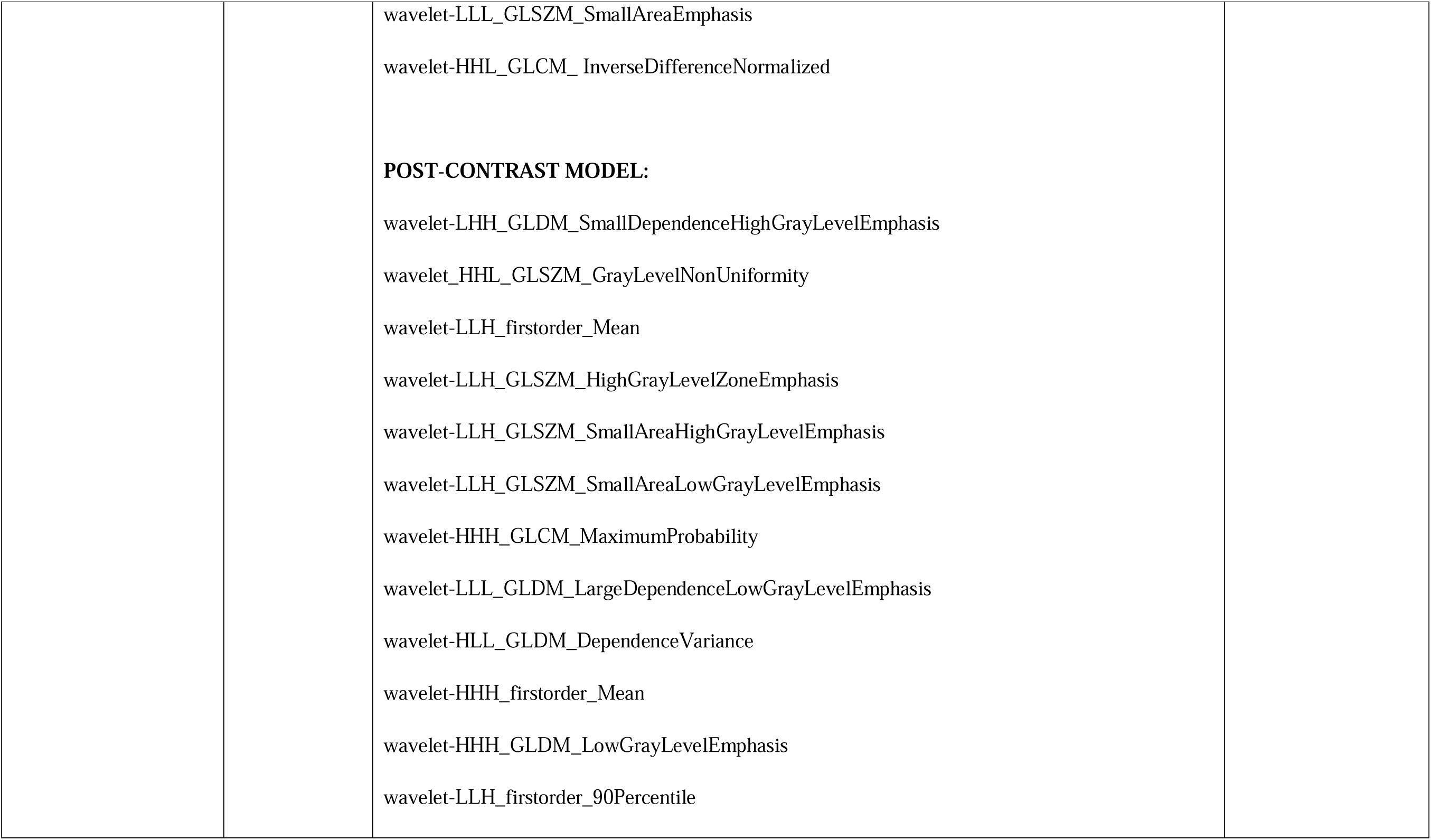
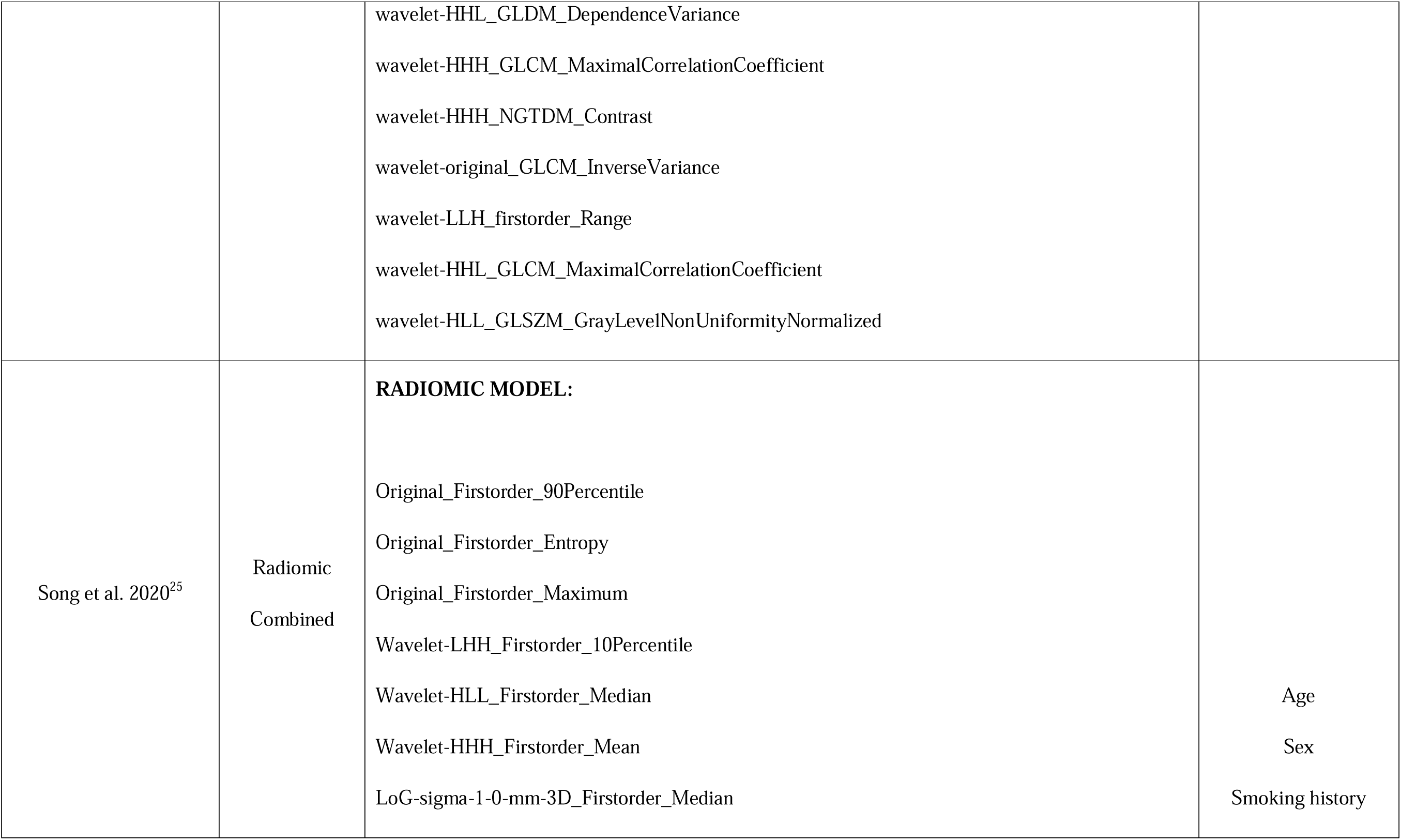
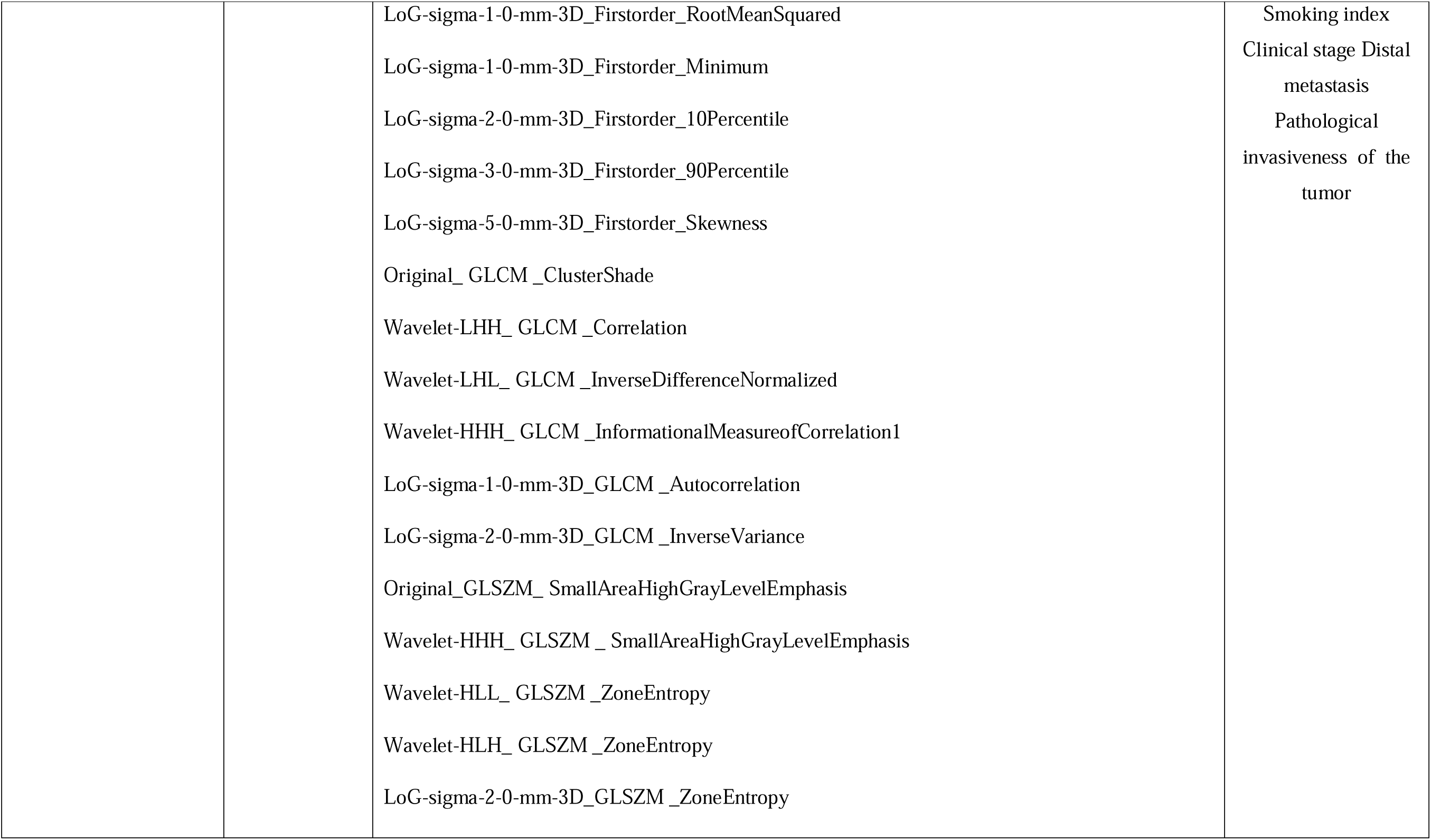
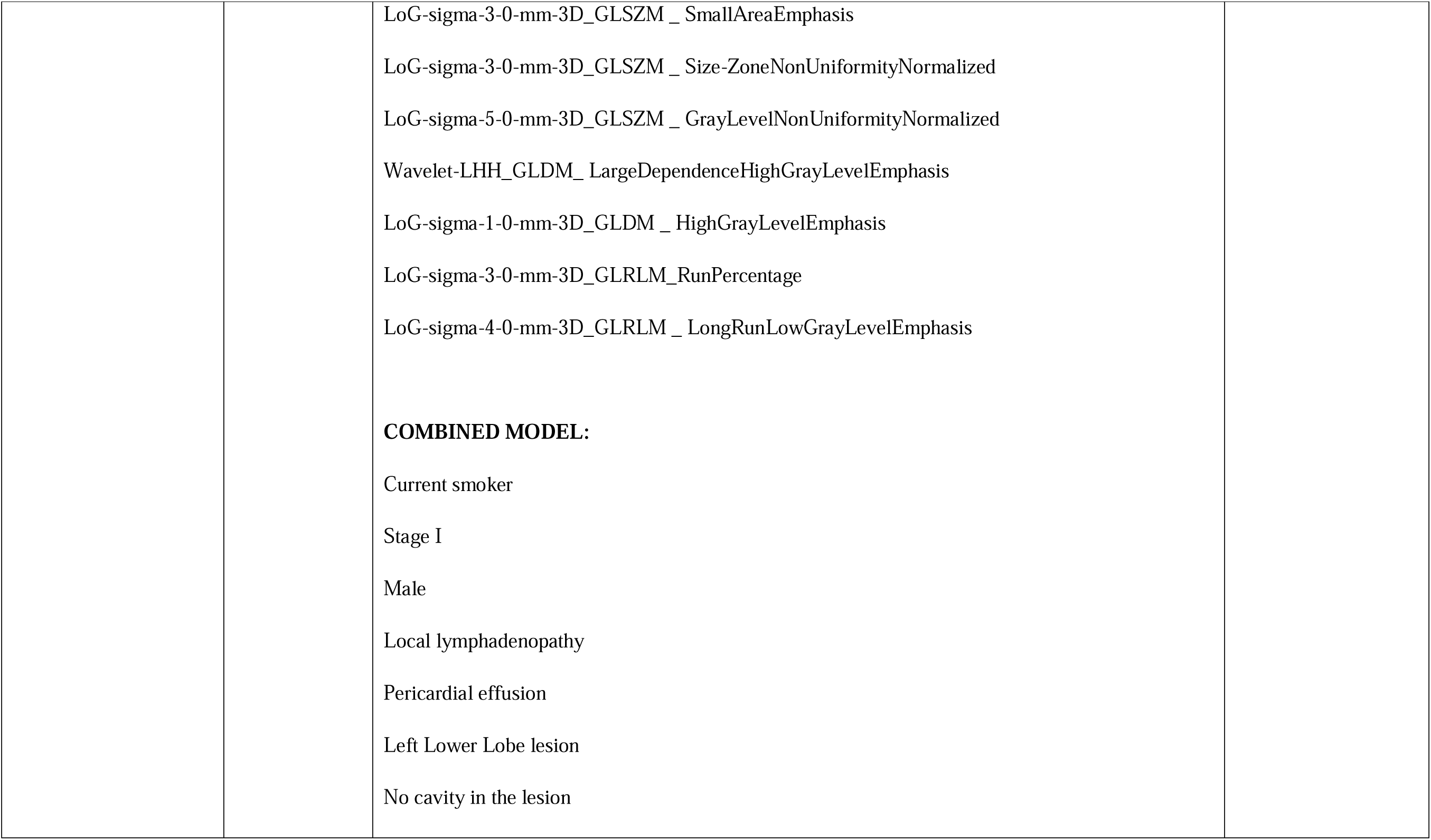
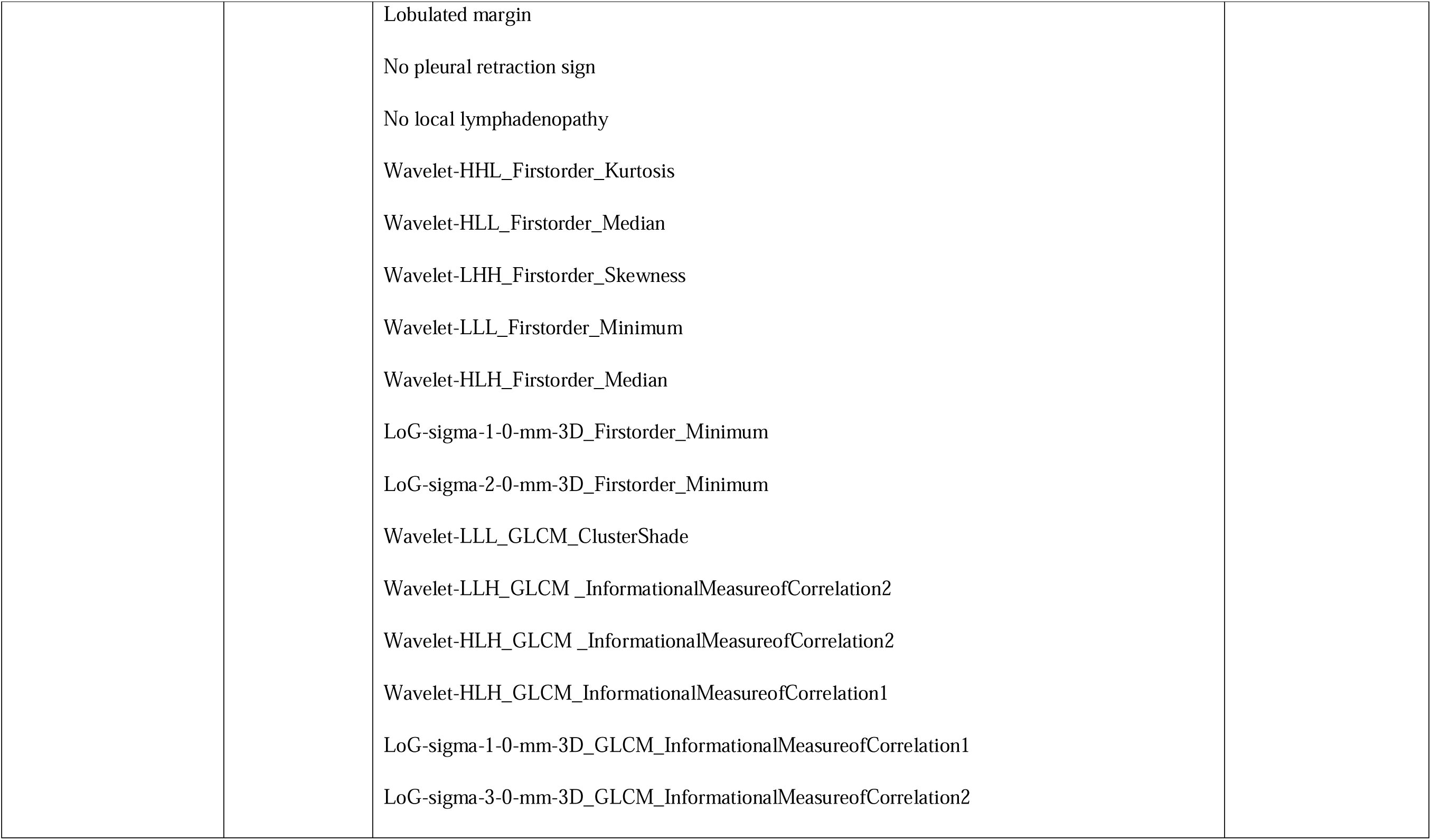
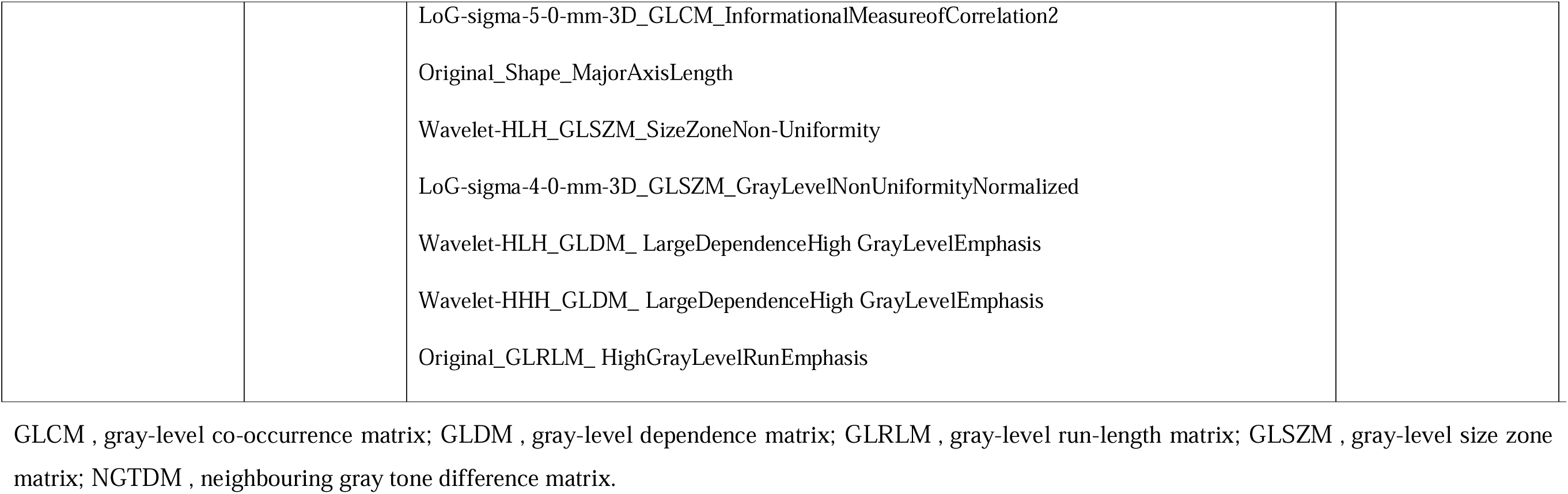
Type of models (radiomic model or combined [radiomic features + clinical variables]) developed in the studies for ALK prediction and the radiomics/clinical features included. ALK, anaplastic lymphoma kinase.

**Supplementary Table S6.** Type of models (radiomic model or combined [radiomic features + clinical variables]) developed in the studies for KRAS prediction and the radiomics/clinical features included. KRAS, Kirsten rat sarcoma viral oncogene homologue.

| Study | Models | Radiomic features | Clinical variables |
| --- | --- | --- | --- |
| Dong et al. 2021<sup>3</sup> | Radiomic | Not specified | — |
| Le et al. 2021<sup>10</sup> | Radiomic | wavelet-LLHGLSZMLargeAreaEmphasis<br>wavelet-LLLGLDMDependenceEntropy<br>wavelet-LHHGLDMLargeDependenceLowGrayLevelEmphasis<br>ori-firstorderkurtosis<br>wavelet-HLHGLCMInverseVariance<br>wavelet-HLLGLSZMSmallAreaHighGrayLevelEmphasis<br>wavelet-LHHGLCMId<br>wavelet-HHLGLCMDifferenceEntropy | — |
|  |  | wavelet-LLLGLSZMGrayLevelNonUniformityNormalized<br>wavelet-HHHGLCMDifferenceAverage<br>wavelet-HHHGLDMDependenceEntropy |  |
| RiosVelazquez et al.<br>2017<sup>23</sup> | Radiomic<br>Combined | imaging.LoG_sigma_3_mm_3D_GLSZM_highIntensityLarteAreaEmp<br>imaging.Wavelet_LHH_GLCM_clusProm imaging.Wavelet_LHH_GLCM_energy<br>imaging.Wavelet_LLL_stats_energy imaging.Wavelet_LLL_stats_median<br>imaging.LoG_sigma_3_mm_3D_GLSZM_largeAreaEmphasis<br>imaging.Wavelet_HHH_GLSZM_lowIntensitySmallAreaEmp<br>imaging.Wavelet_HHH_GLCM_correl1 imaging.LoG_sigma_3_mm_3D_GLCM_clusProm<br>imaging.Wavelet_LLL_GLSZM_highIntensityLarteAreaEmp imaging.Wavelet_HHL_stats_energy<br>imaging.Wavelet_HLL_stats_var imaging.Wavelet_HLH_GLSZM_lowIntensitySmallAreaEmp<br>imaging.Wavelet_LHH_rlg1_GrayLevelNonuniformity<br>imaging.LoG_sigma_3_mm_3D_GLSZM_lowIntensitySmallAreaEmp imaging.GLCM_clusShade<br>imaging.Wavelet_LHH_GLCM_invDiffmomnor imaging.Wavelet_HLL_stats_min<br>imaging.Wavelet_LLL_rlg1_longRunHighGrayLevEmpha imaging.Wavelet_LLH_stats_mean | Stage<br>Sex<br>Smoking status<br>Age<br>Race |
| Wang et al. 2022<sup>28</sup> | Radiomic | CT_square_GLSZM_SizeZoneNonUniformityNormalized<br>CT_wavelet-LHH_GLDM_DependenceNonUniformityNormalized | — |
|  |  | CT_wavelet-HHL_firstorder_Skewness |  |
|  |  | CT_wavelet-HHL_GLDM_DependenceNonUniformityNormalized |  |

GLCM , gray-level co-occurrence matrix; GLDM , gray-level dependence matrix; GLRLM , gray-level run-length matrix; GLSZM , gray-level size zone matrix.

**Supplementary Table S7.** Results of the meta-regression analyzing the effects of age, type of segmentation (manual/semi-automatic/automatic), type of model (radiomics/combined [radiomic features + clinical data) and artificial intelligence methodology (machine learning/deep learning).

| AGE |  |  |  |  |  |
| --- | --- | --- | --- | --- | --- |
| Fixed-effects coefficients |  |  |  |  |  |
|  | Estimate | SE | z | p-value | CI 95% |
| tsens.(Intercept) | 4.488 | 2.181 | 2.058 | 0.040 | [0.214, 8.762] |
| tsens.AGE | -0.052 | 0.035 | -1.483 | 0.138 | [-0.121, 0.017] |
| tfpr.(Intercept) | -0.156 | 2.288 | -0.068 | 0.946 | [-4.640, 4.328] |
| tfpr.AGE | -0.012 | 0.037 | -0.318 | 0.750 | [-0.084, 0.061] |
| Variance components: between-studies Std. Dev and correlation matrix |  |  |  |  |  |
|  | SD |  | tsens |  | tfpr |
| tsens | 0.395 |  | - |  | 0.819 |
| tfpr | 0.431 |  | 0.819 |  | - |
| TYPE OF SEGMENTATION |  |  |  |  |  |
| Fixed-effects coefficients |  |  |  |  |  |
|  | Estimate | SE | z | p-value | CI 95% |
| tsens.(Intercept) | 1.217 | 0.127 | 9.599 | 0.000 | [0.968, 1.465] |
| tsens.SegmentationSemiautomatic | -0.086 | 0.241 | -0.356 | 0.722 | [-0.558, 0.387] |
| tsens.SegmentationUnknown | 0.347 | 0.708 | 0.491 | 0.624 | [-1.040, 1.734] |
| tfpr.(Intercept) | -0.984 | 0.133 | -7.426 | 0.000 | [-1.244, -0.725] |
| tfpr.SegmentationSemiautomatic | 0.272 | 0.254 | 1.067 | 0.286 | [-0.227, 0.770] |
| tfpr.SegmentationUnknown | 0.397 | 0.697 | 0.569 | 0.569 | [-0.970, 1.763] |
| Variance components: between-studies Std. Dev and correlation matrix |  |  |  |  |  |
|  | SD | tsens |  | tfpr |  |
| tsens | 0.465 | - |  | 0.827 |  |
| tfpr | 0.511 | 0.827 |  | - |  |
| CONTRAST |  |  |  |  |  |
| Fixed-effects coefficients |  |  |  |  |  |
|  | Estimate | SE | z | p-value | CI 95% |
| tsens.(Intercept) | 1.244 | 0.608 | 2.045 | 0.041 | [0.052, 2.437] |
| tsens.Contrastcontrast-enhanced | 0.151 | 0.650 | 0.232 | 0.817 | [-1.124, 1.425] |
| tsens.Contrastnon-contrast CT | -0.096 | 0.620 | -0.156 | 0.876 | [-1.312, 1.119] |
| tfpr.(Intercept) | -0.619 | 0.638 | -0.970 | 0.332 | [-1.870, 0.632] |
| tfpr.Contrastcontrast-enhanced | -0.509 | 0.683 | -0.745 | 0.456 | [-1.848, 0.829] |
| tfpr.Contrastnon-contrast CT | -0.309 | 0.650 | -0.476 | 0.634 | [-1.584, 0.965] |
| Variance components: between-studies Std. Dev and correlation matrix |  |  |  |  |  |
|  | SD |  | tsens |  | tfpr |
| tsens | 0.468 |  | - |  | 0.889 |
| tfpr | 0.511 |  | 0.889 |  | - |
| TYPE OF MODEL |  |  |  |  |  |
| Fixed-effects coefficients |  |  |  |  |  |
|  | Estimate | SE | z | p-value | CI 95% |
| tsens.(Intercept) | 1.281 | 0.133 | 9.603 | 0.000 | [1.020, 1.543] |
| tsens.Modelrad | -0.193 | 0.197 | -0.980 | 0.327 | [-0.579, 0.193] |
| tfpr.(Intercept) | -0.939 | 0.146 | -6.413 | 0.000 | [-1.226, -0.652] |
| tfpr.Modelrad | 0.006 | 0.222 | 0.026 | 0.979 | [-0.429, 0.441] |
| Variance components: between-studies Std. Dev and correlation matrix |  |  |  |  |  |
|  | SD |  | tsens |  | tfpr |
| tsens | 0.434 |  | - |  | 0.828 |
| tfpr | 0.524 |  | 0.828 |  | - |
| AI METHODOLOGY |  |  |  |  |  |
| Fixed-effects coefficients |  |  |  |  |  |
|  | Estimate | SE | z | p-value | CI 95% |
| tsens.(Intercept) | 1.215 | 0.232 | 5.2240 | 0.000 | [0.761, 1.670] |
| tsens.TypeML | -0.022 | 0.257 | -0.087 | 0.930 | [-0.526, 0.481] |
| tfpr.(Intercept) | -0.898 | 0.257 | -3.489 | 0.000 | [-1.403, 0.394] |
| tfpr.TypeML | -0.052 | 0.285 | -0.184 | 0.854 | [-0.610, 0.505] |
| Variance components: between-studies Std. Dev and correlation matrix |  |  |  |  |  |
|  | **SD** | **tsens** | **tfpr** |  |  |
| tsens | 0.442 | - | 0.795 |  |  |
| tfpr | 0.520 | 0.795 | - |  |  |

AI , artificial intelligence; CI , confidence interval; ML , machine learning; rad, model including only radiomic features; SE, standard error; SD, Standard deviation; z, standard score in a gaussian distribution; tsens, logarithmic transformation of sensitivity; tfpr, logarithmic transformation of false positive rate.

